# Attention-mediated genetic influences on psychotic symptomatology in adolescence

**DOI:** 10.1101/2024.02.19.24303048

**Authors:** Sarah E. Chang, Dylan E. Hughes, Jinhan Zhu, Mahnoor Hyat, Sullivan D. Salone, Zachary T. Goodman, Joshua L. Roffman, Nicole R. Karcher, Leanna M. Hernandez, Jennifer K. Forsyth, Carrie E. Bearden

## Abstract

Attention problems are among the earliest precursors of schizophrenia (SCZ). Here we examine relationships between multi-trait polygenic scores (PGS), psychotic spectrum symptoms, and attention-related phenotypes in an adolescent cohort (ABCD; n=11,855, mean baseline age: 9.93). Across three biennial visits, greater attentional variability and altered functional connectivity were associated with severity of psychotic-like experiences (PLEs). In European ancestry youth, neuropsychiatric and cognitive PGS were associated with greater PLE severity and greater attentional variability; notably, the effect of multi-trait PGS on PLEs weakened over time. Attentional variability partially mediated relationships between multi-trait PGS and PLEs, explaining 4-16% of these associations. Lastly, multi-trait PGS parsed by developmental co-expression patterns were significantly associated with greater PLE severity, though effect sizes were larger for genome-wide PGS. Findings suggest that broad neurodevelopmental liability is implicated in pathophysiology of psychotic spectrum symptomatology in adolescence, and attentional variability may act as an intermediate between risk variants and symptom expression.

## INTRODUCTION

Against a backdrop of generalized cognitive deficit, attention is among the most significantly impaired domains in schizophrenia (SCZ)^1,2^. In individuals who later develop SCZ, attention problems often present long before the onset of psychotic symptoms^1,3-8^,. Moreover, attention deficits are considered to be some of the earliest signs for psychosis risk^2,9^ and are observed in first-degree relatives of SCZ patients^10,11^. Collectively, findings suggest that attentional impairment and accompanying alterations in neurodevelopment may reflect genetic liability to SCZ^12-18^. While hundreds of genome-wide variants associated with SCZ have been identified in adults^19^, less is known about the genetic etiology of psychotic spectrum symptoms during critical developmental periods. Notably, emerging work in typical adolescence indicates that polygenic scores (PGS) for attention deficit hyperactivity disorder (ADHD) and neurodevelopmental disorders, but not SCZ, are associated with subthreshold psychotic symptoms^14,20-22^. Conversely, SCZ PGS has been associated with broad childhood and adolescent psychopathology^23^. Here, we seek to expand knowledge about genetic liability associated with psychosis spectrum symptoms and related neurodevelopmental processes. Understanding of such mechanisms may aid early intervention for psychosis-spectrum conditions and improve functional outcomes.

SCZ risk variants impact both attention and functional connectivity (FC) of brain networks that support cognition^24-26^. Intra-individual variability (IIV) in reaction time, a key behavioral metric of attention lapses, is elevated in patients with SCZ, their first-degree relatives, and those at clinical high risk for psychosis^27-30^. Moreover, IIV shows genetic overlap with SCZ and ADHD^24,31,32^, and IIV impairments in SCZ are stable across five years^33,34^. On a neural level, reduced IIV, indicating stable cognitive performance, is associated with stronger negative correlation (‘anticorrelation’) between the default mode network (DMN) and task-positive networks (TPNs)^35,36^. DMN-TPN anticorrelation may be a fundamental property of functional network organization, and is thought to support attention allocation by focusing on the task at hand (greater TPN activity) and suppressing internally-directed thoughts (lower DMN activity^37^). Imbalance within and between these networks may play an important role in SCZ pathophysiology and the psychosis spectrum^38-40^ and is associated with cognitive deficits in SCZ^41,42^. Moreover, family studies show that DMN-TPN anticorrelation is heritable^40,43^; yet whether DMN-TPN anticorrelation is linked with genetic liability for SCZ is unknown. Increasing evidence also suggests that FC may serve as a mediator between SCZ genetic risk and cognition^44,45^. Yet to our knowledge, no study has interrogated FC as a mediator between genetic risk and psychosis spectrum symptoms in development. As attention and functional network organization dynamically change in adolescence^46,47^, studies that investigate genetic risk and its links to these phenotypes must consider underlying developmental processes.

Adolescence is a critical maturational period, marked by dynamic changes in cognition and functional brain network reorganization, and also represents the major risk period for development of psychosis^48,49^. In normative development, IIV decreases dramatically from childhood through adolescence, indicating a major developmental shift in sustained attention^46^. On a neural level, associative functional networks become more functionally segregated (i.e., stronger within-network FC, weaker between-network FC) with maturation, while networks recruited in executive function tasks become more integrated^50-54^. Specifically, the DMN and TPNs exhibit increasingly anticorrelated activity with age^55,56^. As pathophysiological processes in SCZ vary in onset and timing, SCZ risk variants may vary in their developmental expression^57-60^ or spatiotemporal gene expression properties^20^. As such, examination of longitudinal gene-brain-behavior relationships during adolescence may allow us to uncover proximal pathways wherein genetic risk is reflected in the emergence of psychosis spectrum symptoms.

The present study leverages the translational potential of multi-trait polygenic scores in the context of the Adolescent Brain and Cognitive Development (ABCD) study^61^ (**Supplementary Table 1**) to understand relationships among genetic risk, attentional variability, attention-related functional brain architecture, and subclinical psychotic-like experiences (PLEs) across early adolescent development. Within this framework, we tested whether: (1) IIV and/or attention-related FC are related to PLEs; (2) PGS related to attention and psychosis (i.e., PGS for ADHD, SCZ, cross-disorder neurodevelopment (NDV), IIV, and broad cognitive performance) are associated with attention-related FC, IIV, and/or PLEs; and (3) whether effects of PGS on PLEs are mediated by IIV or FC. We anticipated that: 1) elevated IIV and weaker DMN-TPN anticorrelation would be associated with PLE severity; 2) higher PGS for SCZ, ADHD and NDV and lower PGS for cognitive performance would be associated with more severe PLEs; and 3) attentional lapses (i.e., greater IIV) and/or within- and between-network FC will mediate the relationships between psychiatric and cognitive PGS on PLEs.

Given the neurodevelopmental nature of SCZ^62^, we conducted exploratory analyses to interrogate how polygenic risk for PLEs unfolds across development. First, we investigated whether associations between PGS and PLEs varied by study visit. Secondly, we tested whether partitioning PGS by previously identified modules of developmentally coexpressed genes^63^ from postmortem brain transcriptomic data^64^ drove any observed relationships between PLEs and PGS. We reasoned that partitioning risk variants by developmental co-expression may offer insight into the neurodevelopmental mechanisms that confer unique risk to PLEs in adolescents. By examining the developmental sensitivity of PGS-PLE relationships, we may identify optimal developmental periods for intervention.

## RESULTS

### 1. Greater reaction time variability (IIV) and altered functional connectivity are associated with more severe psychotic-like experiences in youth

Across three biennial visits across ages 9-15, greater IIV was associated with greater PLE severity (**Table 1**, model marginal R^2^ = 0.063); this was consistent across all three reaction-time tasks from the NIH Toolbox (**Supplementary Table 2**). In participants that passed the motion threshold per ABCD recommendations (https://wiki.abcdstudy.org/release-notes/imaging/quality-control.html**, Supplementary Methods, Figure 1, Supplementary** Figure 1 for N by timepoint), we found that weaker anticorrelation between the default mode network (DMN) and dorsal attention network (DAN), weaker anticorrelation between the DMN and cingulo-opercular network (CON) and weaker FC within the DAN, CON, and DMN were also significantly associated with more severe PLEs (**Table 1**, model marginal R^2^ range = 0.056-0.057). We repeated this analysis in the subset of subjects who passed more rigorous resting-state scan quality control^65^ (**Supplementary Methods**); the associations between weaker DMN-DAN anticorrelation and weaker within-network FC in DAN and PLE severity were maintained but there were no significant associations between DMN-CON anticorrelation or within-network FC of the CON with PLEs (**Supplementary Table 3**).

**Figure 1.**
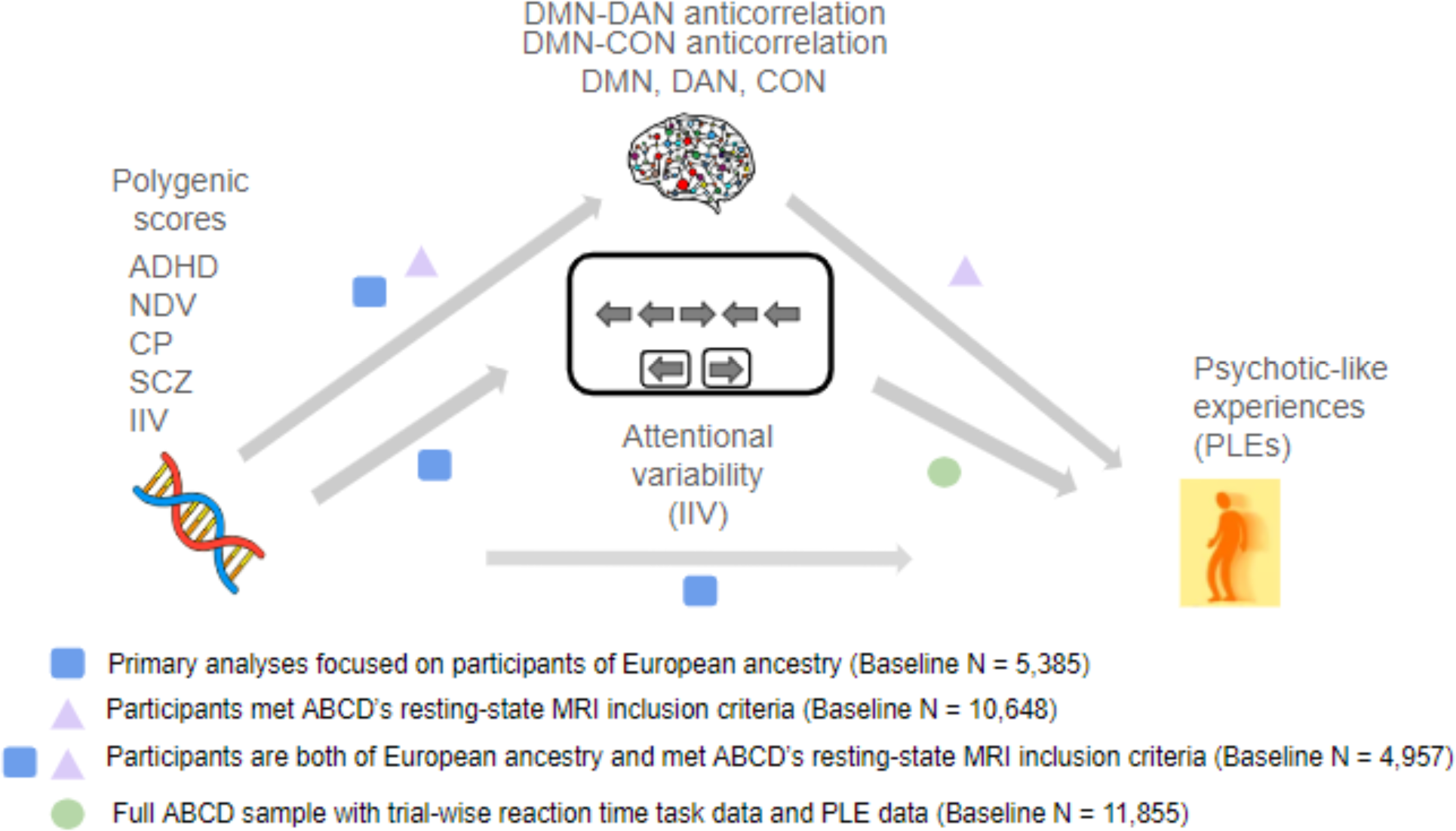
Study overview of associations tested. (1) Between IIV and PLEs; (2) Between FC and PLEs; (3) between PGS and FC; (4) PGS and IIV; (5) PGS and PLEs; N is based on the number of participants with complete PQ-BC information at the baseline study visit. Abbreviations: ADHD (attention-deficit/hyperactivity disorder); NDV (neurodevelopmental disorders); CP (cognitive performance); SCZ (schizophrenia); IIV (intra-individual variability); DMN (default mode network); DAN (dorsal attention network); CON (cingulo-opercular network); PLEs (psychotic-like experiences). See Supplemental Figure 1 for N’s across timepoints.

**Table 1.**
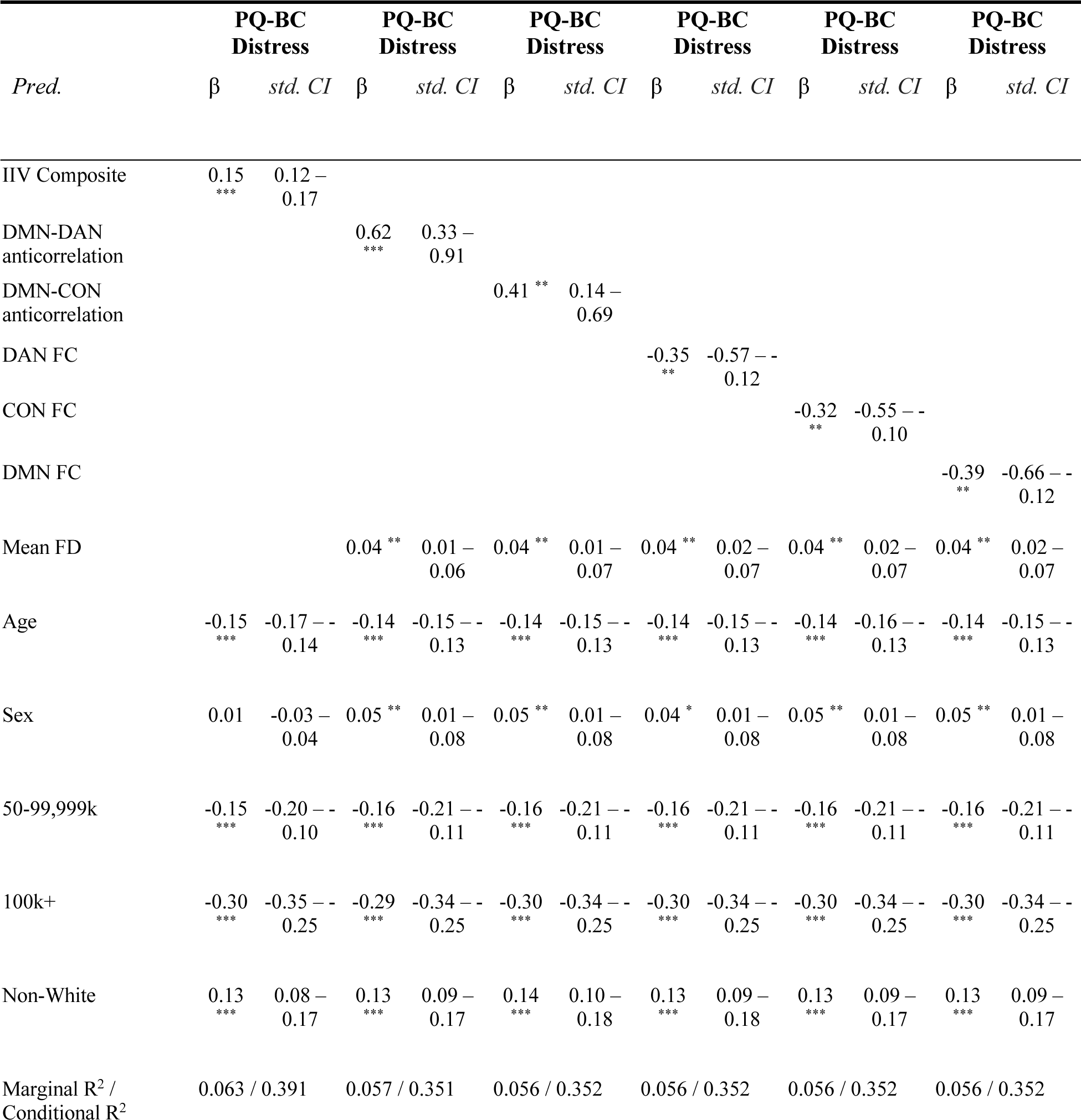
Linear Mixed Models: Associations between Attentional Phenotypes and PLE Severity (PQ-BC Distress Score). Each column (including β and standard CI) reflects a separate model, where PQ-BC Distress is the dependent variable across all models. Covariates include mean FD, age, sex, parental income, and race/ethnicity. Includes all data available across ages 9-15; please see Figure 2 for data availability for each measure. **Abbreviations:** Pred = Predictors, PQ-BC = Prodromal Questionnaire - Brief Child Version; IIV = intra-individual variability; DMN = default mode network; DAN = dorsal attention network; CON = cingulo-opercular network; FC = functional connectivity; FD = framewise displacement. **Reference categories:** Sex (male); Parental Income (<50k); Race/Ethnicity (White). **Significance:** * q<0.05 ** q<0.01 *** q<0.001. Marginal R^2^ is interpreted as the variance explained in the model by the fixed effects only; conditional R^2^ is the variance explained by the fixed and random effects

### 2. Polygenic scores for multiple psychiatric disorders and cognition are associated with greater attentional variability and more severe PLEs

In participants of European descent, aged 9-15 (n=5,385), higher PGS for ADHD, NDV, and IIV, and lower PGS for cognitive performance were significantly associated with greater IIV, but there was no association between SCZ PGS and IIV (**Table 2**, **Supplementary Table 4,** model marginal R^2^ range = 0.100-0.109). Moreover, higher PGS for SCZ, ADHD, NDV, and lower PGS for CP were all associated with more severe PLEs (**Table 2**, **Supplementary Table 5** marginal R^2^ range = 0.026-0.035), but there was no association between IIV PGS and PLEs. No PGS were associated with either DMN-DAN anticorrelation or within-network FC of the DAN (n=4,957 participants of European descent, **Supplementary Tables 6 and 7**).

**Table 2.**
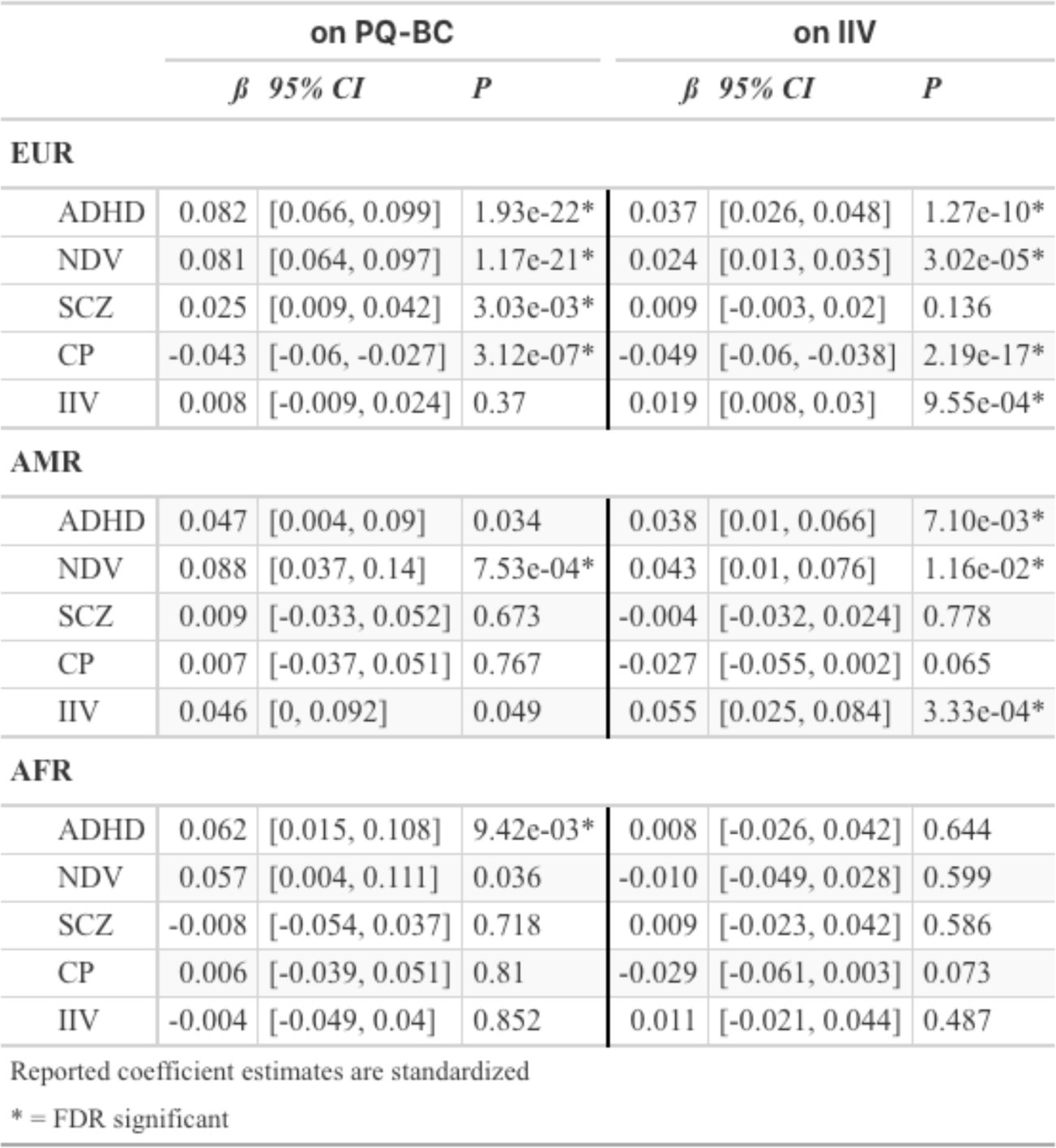
Results from models regressing PGS on PQ-BC and IIV across data collection waves. Effect sizes (ß), confidence intervals, and P-values reported from linear mixed effects models regressing PGS on PQ-BC (left) and composite IIV (right). Covariates include age, sex, and the top 5 genetic PCs as fixed effects with subject, nested within family, within site. EUR = European ancestry (Baseline N = 5,385), AMR = American/Latinx ancestry (Baseline N = 1,428), AFR = African ancestry (Baseline N = 1,644). Asterisks indicate FDR significance correcting for 30 comparisons (5 PGS x 3 populations x 2 dependent variables). Additional abbreviations: PQ-BC = Prodromal Questionnaire - Brief Child Version. NDV = neurodevelopmental disorders, SCZ = schizophrenia, CP = cognitive performance IIV = intra-individual variability.

### 3. Associations between PGS, IIV, and PLEs in non-European participants

For AMR participants, greater ADHD and NDV PGS were associated with more severe PLEs, and greater NDV and IIV PGS were associated with increased IIV. In AFR subjects, greater ADHD PGS was associated with more severe PLEs (**Table 2**). Additional associations between non-EUR PGS, IIV, and PLEs were directionally consistent with those in EUR, but did not survive correction for multiple comparisons (**Table 2**). Results from non-European participants should be interpreted with caution (see Discussion).

### 4. IIV mediates the relationship between neuropsychiatric and cognitive polygenic scores and PLEs

To investigate our hypotheses that attentional variability mediates the relationship between polygenic scores and PLEs in adolescence, in the European ancestry subset (N=5,385) we tested for statistical mediation, as elevated IIV was associated with both neuropsychiatric and cognitive PGS and more severe PLEs. We found indirect effects via IIV between greater PGS for ADHD, NDV, and CP and PLE severity, according to 10,000 simulations of quasi-Bayesian bootstrapping. The proportion of the associations between CP, ADHD, and NDV PGS and PLEs mediated by IIV was 16%, 7%, and 4% respectively **(Figure 2a, b, c**).

**Figure 2.**
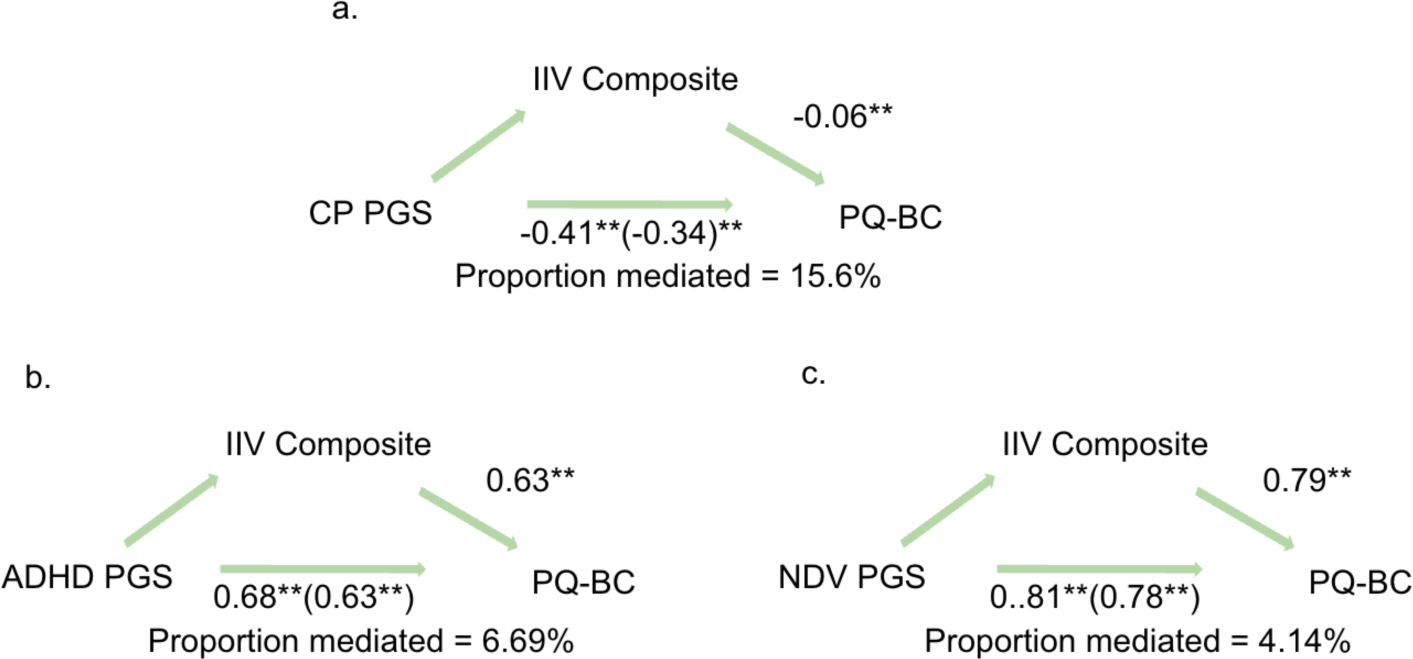
Mediation models. Standardized beta values in parentheses take into account the effect of IIV on PQ-BC. Abbreviations: CP (cognitive performance); ADHD (attention deficit/hyperactivity disorder); NDV (neurodevelopmental disorders); IIV (intra-individual variability). Significance: * q<0.05 ** q<0.01 *** q<0.001

### 5. Time-varying effects of PGS on PLE severity

Across 5 yearly visits, the associations between PGS for ADHD, NDV, and CP and PLEs weakened over time (**Figure 3, Supplementary Tables 8-11**). In contrast, there was no significant interaction between SCZ PGS and time.

**Figure 3.**
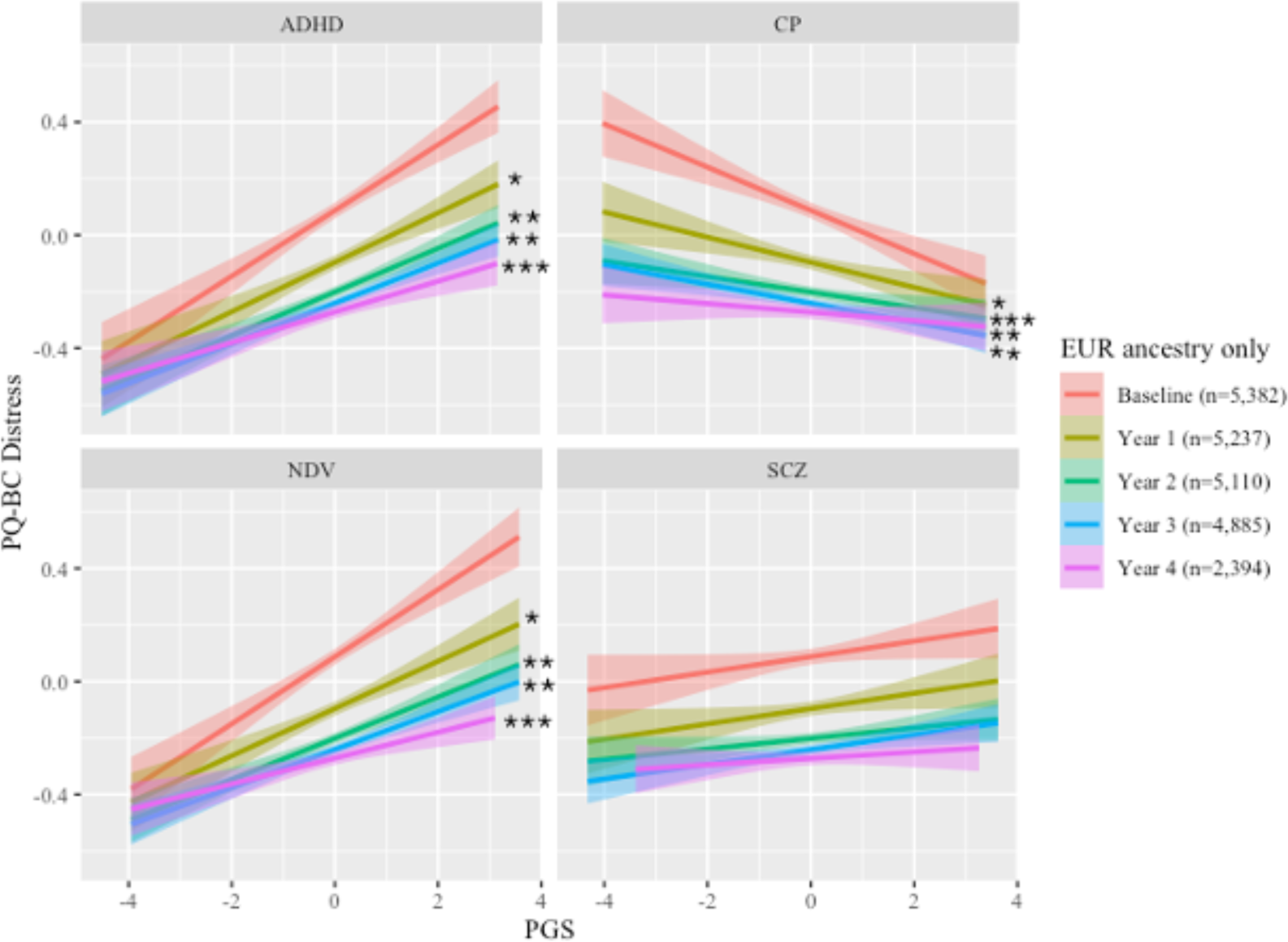
Time-varying effects of PGS on PQ-BC Distress. Relationships between PGS for ADHD, NDV (neurodevelopmental disorders) and cognitive performance (CP) with PQ-BC Distress weaken with increasing age, whereas SCZ (schizophrenia) PGS* time interaction was not significant. Standardized (z-transformed) PGS and PQ-BC values are represented on the x- and y-axis. Shading around the best-fit lines indicates standard error. Stars indicate significant difference in the interaction term for a given time point and PGS*Baseline. All analyses control for age, sex, and the top 5 genetic principal components as fixed effects, with subject nested within family within site. * q<0.05 ** q<0.01 *** q<0.001

### 6. Brainspan partitioned PGS (pPGS) and PLEs

In the EUR sample, several module pPGS exhibited significant associations with PLE scores in expected directions; however, effect sizes were notably smaller compared to genome-wide PGS (**Figure 4**). ADHD and NDV module pPGS showed similarly diffuse and largely overlapping signal across modules. One CP module pPGS, M1, showed significant negative associations with PLE severity after multiple comparison correction. No effects of SCZ module pPGS on PQ-BC remained significant after multiple comparison correction. There was a significant, moderate correlation between size of module (number of SNPs) and effect size, such that larger modules typically showed larger associations between pPGS and PQ-BC (Pearson’s *r* = 0.48, P = 1.85x10^-5^; **Supplementary** Figure 2). Indeed, results from permutation tests, controlling for module size, indicated that only one pPGS (ADHD M5) in EUR performed significantly better than pPGS derived from an equal number of randomly selected variants (**Figure 4, Supplementary Table 12**). The observed association of ADHD M5 pPGS with PLE severity fell within the top 5% of the distribution of permuted null effects (ß=0.029, 95% CI [0.012 - 0.045], P = 6.25x10^-4^, permutation P = 0.03).

**Figure 4.**
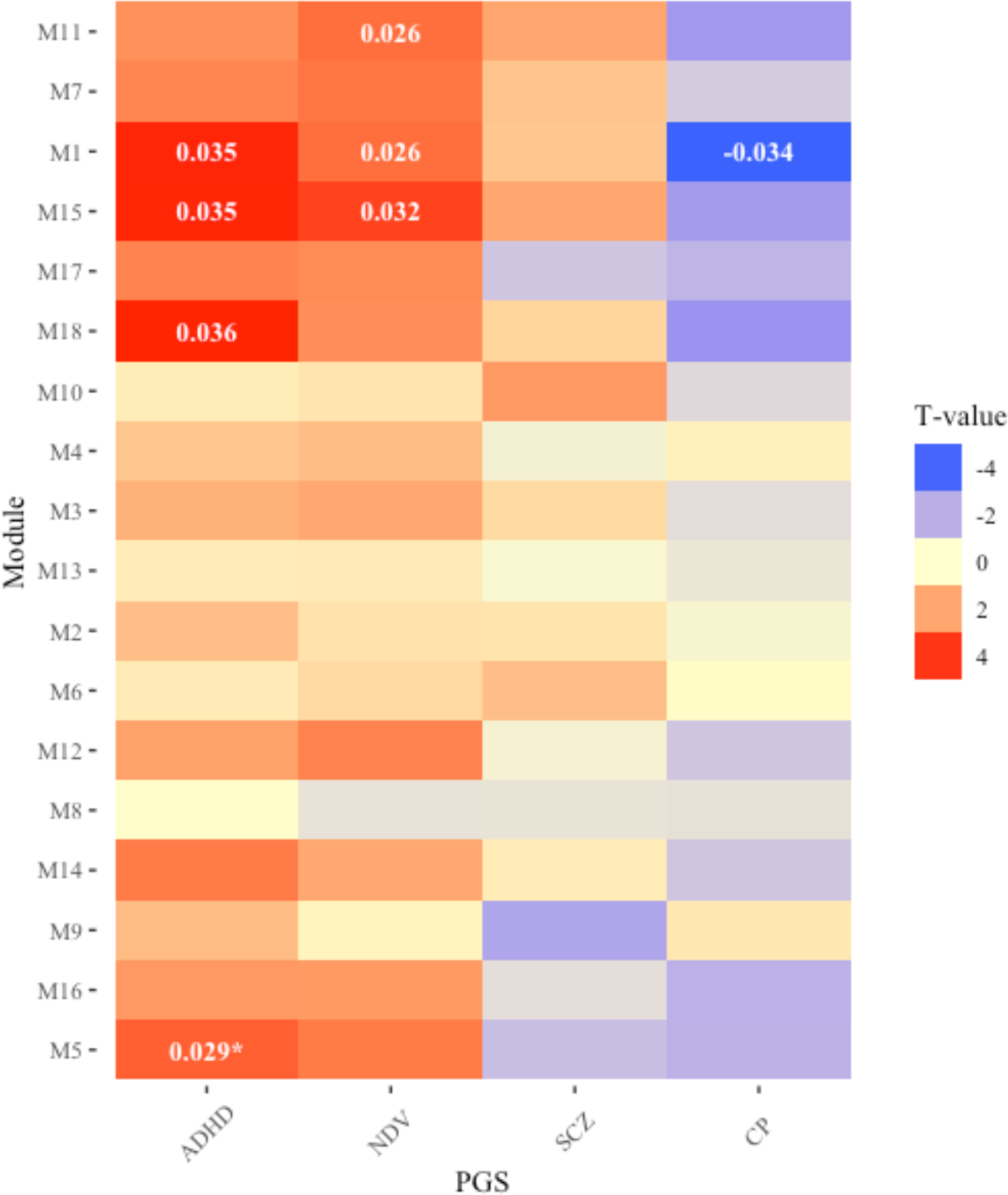
Effects of polygenic score, partitioned by modules of developmentally co-expressed genes (pPGS), on PQ-BC Distress in the European sub-sample. Modules are ordered from bottom to top, by smallest to largest module. Each column represents a different PGS. Standardized effect sizes of modules that survive FDR correction (n=216 including tests with nonEUR PGS) are indicated within its respective cell. Effects with an asterisk indicate effects that survived FDR correction and were also significant after controlling for module size via permutation test (ADHD M5 only).

Results from pPGS analyses of non-EUR samples were likely underpowered compared to EUR pPGS, with no single module surviving FDR correction (**Supplementary** Figure 3). Indeed, effect sizes of randomly generated EUR PGS were significantly larger than those of non-EUR PGS (average unstandardized absolute difference = 0.18, range = 0.014 - 0.366, P-values < 2.10^-16^). For non-EUR modular pPGS that were significantly associated with PQ-BC before FDR correction, several showed significance via permutation and competitive p-value tests (see **Supplementary Tables 12 and 13**).

## DISCUSSION

Our longitudinal investigation of the genetic architecture and neural substrates of attentional variability and subthreshold psychotic symptoms across early adolescence revealed several novel findings. First, greater attentional variability and altered connectivity within and between attention-related networks are associated with greater PLE severity. Secondly, genetic predisposition for several neurodevelopmental disorders (ADHD, cross-disorder neurodevelopment, and SCZ) and lower cognitive performance are associated with greater attentional variability and PLE severity, with PGS partitioned based on developmental co-expression patterns showing weaker associations with PLE severity than genome-wide PGS. The associations between cognitive, ADHD and broad neurodevelopmental PGS and PLE severity weaken with increasing age, whereas the association between SCZ PGS and PLEs does not. Lastly, attentional variability partially mediates the relationship between multi-trait PGS and PLEs. Together, these findings suggest that genetic liability related to broad neurodevelopmental disorders? may manifest as subthreshold psychotic symptoms in adolescence, partially through attention lapses. Understanding the pathophysiology of shared genetic vulnerability of established clinical phenotypes and emerging PLEs in early adolescence is critical for early intervention.

Our finding that SCZ PGS is associated with dimensionally measured PLEs in a population-based study of early adolescence align with those in the ALSPAC cohort^21^, adding to associations between SCZ PGS and disorganized symptoms found in adults with SCZ^22^ and PLEs in typically developing,older adolescents^67^ (aged 15-19). While two prior studies using ABCD data have found null associations between SCZ PGS and PLEs^20,68^, our analyses differ in several ways. Our analyses generate PGS from a 2022 GWAS of 53,386 EUR individuals with SCZ^19^, whereas previous work uses a smaller, 2018 GWAS with 40,675 cases^69^. Secondly, we leverage PLE symptomatology across ages 9-15, compared to age 9-10^20,68^. Lastly, using the latest recommendations in genomics, our methodological pipeline overcomes bias from including related individuals, resulting in a larger sample. As such, power from these analytic choices and inclusion of PLEs across a wider, older age range may have allowed us to detect subtle associations between SCZ PGS and PLEs in early adolescence.

We also find that PGS for other broad neurodevelopmental indicators (higher PGS for ADHD and NDV, lower PGS for cognition) were consistently associated with more severe PLEs in EUR youth, with greater strength than SCZ PGS. These results replicate and extend associations between ADHD and NDV PGS and PLEs previously found in typically developing adolescents^20^ and youth at clinical high risk for psychosis^14^. Yet while SCZ is only weakly genetically correlated with ADHD, relative to adult-onset diagnoses like bipolar disorder^70^, ADHD symptoms and subthreshold psychotic symptoms in youth are phenotypically correlated^14,70,71^. We also find that PGS for cognition is negatively associated with PLEs, aligning with work showing that PGS for nonverbal IQ and educational attainment are associated with lower likelihood of SCZ diagnosis^72.73^. Moreover, there is genetic overlap between SCZ and IQ, though this may vary by cognitive domain^74-76^. Findings in non-EUR samples are largely directionally consistent with those in EUR samples. In fact, of the associations that are significant in the non-EUR group, effect sizes are comparable to, if not stronger than, those observed in the EUR group. These results suggest that, despite PGS typically performing poorly in predicting outcomes in non-EUR groups, similar biological pathways may confer risk to PLEs across ancestries. Collectively, our convergent PGS findings suggest that subthreshold psychotic symptoms in young adolescents in the general population are influenced by genetic variants that are not specific to schizophrenia, the effects of which vary by time. That the effect of broad risk variants on PLEs weakens across early adolescence may reflect a differentiation of the psychosis phenotype that begins to occur over the same developmental period.

As predicted, higher polygenic scores for NDV, ADHD, and IIV, and lower polygenic scores for CP, are associated with greater attentional variability, extending previous findings in ADHD to typical development^77.78^. Cognitive abilities in healthy individuals and cognitive deficits in schizophrenia are highly heritable^79-81^. Furthermore, inverse relationships between schizophrenia PGS and cognition are observed in population-based samples^72,82-89^ and in those at high risk for psychosis^90^. Interestingly, some studies of patients with SCZ have found an association between SCZ PGS and cognition^66,91-95^, while others have not^96-99^. There is likely a non-uniform association between genetic risk variants and cognition^98^. As such, we split the PGS into sets of developmentally co-expressed genes in humans, to determine whether the observed polygenic signal may be accounted for by more specific neurodevelopmental pathways.

In contrast to previous work conducted in patients with SCZ^26,100^, healthy adults^101-103^, and a large sample of individuals across the psychosis spectrum from childhood to adulthood^104^, here we did not find associations between SCZ PGS and resting-state FC, nor any other PGS, in participants of EUR ancestry. There are several potential explanations for these null findings. First, given our *a priori* focus on attention, our analyses focus on connectivity within and between three functional networks, limiting our investigative scope. Second, genetically driven patterns of FC alteration may potentially emerge later in development. These null findings align with Passiatore and colleagues^104^, who find no association between PGS and FC across ages 8-15, but did so at older ages. Third, associations between SCZ PGS and alterations in functional connectivity detected in adults in prior studies are subtle (eg. β=-0.02, from 294,000+ subjects^101^), whereas our sample is much smaller and younger. Lastly, altered DMN-TPN connectivity in psychosis spectrum conditions may be more closely tied to environmental risk factors, warranting further investigation.

To our knowledge, we are the first to show that attentional variability mediates the relationship between PGS and subthreshold psychotic symptoms in early adolescence. This novel finding broadly aligns with previous work showing that PGS increases schizophrenia risk and symptoms through cognition-relevant pathways in SCZ and healthy adults^95,105^. However, different cognitive phenotypes may vary in their relationship with genetic etiology of SCZ^99^. The current findings harness attentional variability as cognition undergoes its steepest rate of development in adolescence^106^ (roughly ages 10-15). This developmental period is also when altered cognitive development emerges in those who later develop schizophrenia^1,8^. Future investigation of the molecular basis of cognition may further our understanding of how genes may converge on constrained pathways^107^ and potentially yield attention deficits that often precede psychosis.

In exploratory analyses, we partition the genome-wide signal for each PGS into 18 subsets of scores, each representing effects associated with gene sets that share brain expression patterns across development. This method represents a powerful, albeit nascent, extension of classical PGS methods, which are agnostic to gene characteristics. In this study, pPGS leverages developmental gene co-expression patterns to probe whether variants involved in specific neurodevelopmental processes might influence PLEs. In EUR subjects, a single module M5, weighted by ADHD polygenic risk, is associated with PLEs, after accounting for module size. Genes in this module are implicated in the organization of synapses, membranes, and vesicles across development, and are enriched for neuronal cell-type markers^63^. Thus, ADHD risk variants involved in synaptic and membrane organization may disproportionately contribute to PLEs, reinforcing the importance of synaptic integrity in biological mechanisms related to psychosis spectrum symptoms.

No other ADHD pPGS survived correction for module size, nor did module pPGS weighted by SNP effects for other psychiatric or neurocognitive traits. In non-EUR samples, results vary, do not converge on any individual module pPGS, and are underpowered (see **Supplementary** Figure 3). These mixed findings likely partially reflect the inadequacy of current methodologies to meaningfully apply PGS methods to non-EUR samples^108^. Permuted null distributions reveal that EUR PGS generated from randomly sampled SNPs systematically exhibit significantly larger effects on PQ-BC than did non-EUR PGS.

While the findings in EUR participants suggest that PLEs may be predominantly driven by SNP effects that are broadly distributed across the genome, testing other gene sets, alternative methods for pPGS, and/or examining other phenotypes may yield new insights. Nonetheless, methods to parse PGS are an active area of development; in certain circumstances pPGS can outperform genome-wide PGS, but interpretations warrant caution^109^ (see Supplement for more detail). Future work may leverage more sensitive, biologically grounded methods to stratify genome-wide risk, towards precision medicine.

## Limitations

Allele frequency, linkage disequilibrium structure, and effects sizes of common variants vary across populations^110-112^. As such, sensitivity of EUR PGS to psychiatric traits in non-European samples is poor. We include multi-ancestry results not to draw any firm conclusions but (1) to demonstrate this crucial equity issue stemming from inadequate power in multi-ancestry GWAS and (2) for transparency and to allow comparisons in future research^113^. Moreover, PGS is a measure of the raw genetic code; measurements like chromatin openness related to cell-types implicated in SCZ may more closely index alterations in specific mechanisms^114^. Future studies could leverage measurements of biological function or interrogate mechanisms in animal models. While these results point to broad neurodevelopmental etiology of PLEs in early adolescence, we underscore that effect sizes for PGS here and elsewhere are very small, limiting clinical utility. Lastly, the results are limited by the absence of a replication sample with comparable, longitudinal measures.

## CONCLUSION

Collectively, our results indicate that broad genetic liability for neurodevelopmental disorders and cognitive performance are associated with subthreshold psychotic symptoms in early adolescence; further, attentional variability may serve as a link between risk variants and symptom expression. By establishing relationships from genomic to behavioral levels of study during this critical stage of cognitive development, we offer a foundation for future mechanistic studies that may identify molecular targets for early intervention.

## Methods

### Section 1: Participants

Data from 11,855 youth were obtained from the ABCD study, a prospective, longitudinal study that tracks youth aged 9-11 at their baseline study visit (mean (sd) age: 9.93(0.63); 47.37% female, **Supplementary Table 1**), for the following 10 years. Participants were recruited from 22 study sites located in all census regions, where recruitment efforts actively enrolled participants from differing socioeconomic and racial/ethnic backgrounds to recapitulate the sociodemographic characteristics of the general US population^115^. All sites obtained informed consent from parents and assent from children, approved by each site’s Institutional Review Board, with centralized IRB approval at the University of California, San Diego. All analyses were conducted using de-identified data from ABCD Data Release 5.1, accessed through the National Institute of Mental Health Data Archive (Collection 2147); thus, this study was exempt from IRB approval. Data collected across 5 visits were used for analysis with each visit on average separated by 12.3 months and each subject having 4.3 timepoints of PLE data, on average, as assessed via the PQ-BC. Imaging and cognitive data were collected only at baseline and even visits (i.e., Baseline, Year 2, and Year 4); thus, analyses of these data included fewer timepoints, with each subject having on average 1.9 timepoints. See **Figure 1** and **Supplementary** Figure 1 for more detail on data availability for each construct and analysis.

### Section 2: Intra-individual Variability

Subjects completed 7 developmentally-appropriate tasks across episodic memory, executive function, attention, working memory, processing speed and language abilities from the National Institutes of Health (NIH) Toolbox^116^. The current study used the timed-reaction time tasks from NIH Toolbox, which includes the Flanker, Pattern Comparison Processing Speed, and Dimensional Change Card Sort tasks. Details of these tasks have been published previously^116,117^ (**Supplementary Methods, section 1**) Intra-individual variability for each task was examined in relation to PLEs and polygenic scores; however, the results for each task were nearly identical in both these analyses (Supplement) and in previous work^36^. Therefore, we generated a cross-task composite score for intra-individual variability (IIV) by calculating standard deviation in reaction time across all correct trials for each task^118,119^. Then, the data for each task were z-scored, and the average of the three z-scores was calculated to generate the IIV composite score for each participant. To remove extremely large outliers, the IIV composite score was winsorized at three standard deviations.

### Section 3: Psychotic-like experiences

Psychotic-like experiences (PLEs) were assessed using the Prodromal Questionnaire - Brief Child Version (PQ-BC), a 21-item, developmentally-appropriate questionnaire that was validated in the ABCD study sample^120^ (**Supplementary Methods, section 2**).

### Section 4: Brain Imaging

Data collection and processing protocols for the ABCD study have been previously described. All participants were scanned on 3T scanners (Siemens Prisma, General Electric, and Phillips) with a 32-channel head coil. Subjects completed T1-weighted and T2-weighted anatomical scans, and four 5-minute resting-state blood oxygen level-dependent (BOLD) scans, with their eyes open, fixed at a crosshair. Further details about the resting state imaging acquisition that varied by 3T scanner have also been detailed previously^61^.

The ABCD Data Analysis, Informatics and Resource Center conducts centralized processing of MRI data across ABCD sites, using the Multi-Modal Processing Stream^121^ (see Supplement). Summary metrics of within- and between-network FC were available in the tabulated data. After processing, within-network FC was calculated by computing pairwise correlations between each ROI in a given network, defined by the Gordon parcellation^122^. For between-network FC, pairwise correlations were calculated between each ROI within the first network and each ROI within the second network. These correlations were averaged and Fisher-Z transformed to generate summary metrics of within- and between-network connectivity strength.

Due to our *a priori* hypotheses, only attention-related functional networks were tested for associations, including DMN-TPN anticorrelation (TPNs included the dorsal attention network (DAN) and cingulo-opercular network (CON)), and within-network FC of the DMN, DAN, and CON. As noted above, weaker DMN-DAN and DMN-CON anticorrelation has been associated with both greater IIV and schizophrenia^36,40^. Further, altered FC within these networks has been related to worse cognition in psychosis spectrum disorders and in the general population^38,39,41,42^,. To reduce the number of statistical tests, associations between PGS and FC were only conducted for FC metrics that were significantly related to PLEs (Supplement). We used the ABCD’s inclusion recommendations for each participant, based on the quality of their resting-state imaging scans (**Supplementary Methods, section 3**). We also conducted secondary analyses using a smaller subset of subjects (see Figure 2 for N by visit) with more stringent motion quality control^65^ (**Supplementary Methods, section 4)**.

### Section 5: Genotyping, imputation, and Quality Control (QC)

Two sets of ABCD genotypes were downloaded from the NIMH Data Archive (NDA): the imputed and the minimally QC’d, pre-imputation (mQC) genotype data, the latter of which was used to assign ancestries to subjects and partition the former into ancestrally homogenous groups for subsequent PGS analyses. Pre-imputation quality control of the ABCD-imputed data can be found here: https://wiki.abcdstudy.org/release-notes/non-imaging/genetics.html. See **Supplementary** Figure 4 for overview of genotyping pipeline.

#### Ancestry assignment

To create ancestrally homogenous groups of subjects, principal components analysis (PCA) of high-quality SNPs from mQC ABCD data merged with 1000 Genomes Project (1KG) data was conducted, followed by a random forest (RF) algorithm to classify ABCD subjects as belonging to one of five superpopulations. To obtain high quality SNPs, quality control was conducted following recommendations outlined by Peterson et al.^123^ for the processing of ancestrally diverse populations. In brief, SNPs were retained if they had < 5% missingness and > 5% minor allele frequency; individuals were removed if they were missing more than 98% of their data and/or if they were derived from plate 461 (as recommended by ABCD). ABCD data were then merged with the 1KG data after removing 1st and 2nd degree relatives (n=14) from 1KG. Strand-flipped, multi-allelic, and strand-ambiguous SNPs were removed as well as SNPs violating Hardy Weinberg Equilibrium at a P-threshold of 0.001.

Using the GENESIS package in R^124^, PC-AiR was used to partition merged genotype data into an unrelated and related group, run PCA on the unrelated sample, and project the related sample onto that PCA space, thereby allowing the inclusion of sibling pairs and other related samples in the analysis sample while eliminating the bias induced by including sibling pairs in genetic PCA^125^ (see **Supplementary** Figure 5). An RF classifier was trained on the 1KG subjects principal components data, for which population labels are known. ABCD subjects were assigned a population label if their predicted probability of a given ancestry membership was greater than 0.9 per recommendations for diverse samples^123^. Due to the small number of ABCD subjects assigned to the South Asian (N=43) and East Asian (N=141) superpopulations, data were only included for European (EUR; N=5,386), Admixed American (AMR; N=1,429), and African (AFR; N=1,644) ancestry subjects. PCA was conducted again within ancestry groups to acquire PC scores for inclusion in main mixed effects models to control for population stratification within ancestry groups. Derived ABCD population labels were used to divide the ABCD-imputed data into 3 groups (EUR, AMR, and AFR).

#### Quality control of ABCD-imputed data

After dividing the ABCD-imputed data into 3 population groups, two subjects were removed per recommendations from ABCD and additional QC was performed within ancestry prior to calculation of PGS. SNPs were removed if they were not in rs format, had missingness greater than 5%, or had minor allele frequencies less than 1%. After filtering, 7,822,323 SNPs remained in the EUR sample; 8,520,435 in AMR; and 13,502,132 in AFR.

#### PGS calculation

Summary statistics from the latest GWAS of psychiatric phenotypes were downloaded from the Psychiatric Genomics Consortium (PGC) database. Psychiatric phenotypes included SCZ^19^, ADHD^126^, autism spectrum disorder^127^ (ASD), major depressive disorder^128^ (MDD), and Tourette’s syndrome^129^ (TS). Of note, only SCZ and ADHD PGS were tested individually, whereas summary statistics for ASD, MDD, and TS were used to calculate the neurodevelopmental (NDV) PGS (see below). NDV was chosen specifically because it has previously been shown to predict both diffuse psychopathology and PLEs over and above other disorder-specific PGS such as ADHD and SCZ^20,130^,. Cognitive performance (CP) represents a well-powered proxy measuring cognitive processes related to attention^82^. Given that broad cognitive deficits are present in SCZ^1^, we hypothesized behavioral attention (indexed by IIV) would account for some, but not all, of the variance in the association between CP PGS and PLEs. Summary statistics for CP were downloaded from Social Science Genetic Association Consortium^131^ (SSGAC); those for IIV were provided by Wootton and colleagues^132^. ADHD was included to index attentional problems and because ADHD PGS has previously shown strong association with PQ-BC^20^.

PRS-CS was used to calculate posterior effects of SNPs that were overlapping between ABCD and each set of summary statistics. We leveraged PRS-CS as previous work in the ABCD study shows that it is the PGS method that shows the strongest association with psychopathology across ancestries^133^. We used default parameters and set the global shrinkage parameter, ɸ, to 0.01 for summary statistics derived from sample sizes of less than 200,000 and allowed PRS-CS to estimate ɸ from those derived from samples of greater than 200,000. Resultant posterior SNP effect sizes were summed across ABCD genotypes using the –score function from PLINK v2. Linkage disequilibrium (LD) blocks for EUR, AFR, and AMR populations were estimated from 1KG data and downloaded from the PRS-CS GitHub repository. Only the SCZ summary statistics have separate statistics available for EUR, AFR, and AMR populations; for estimates of posterior effects from these statistics, the LD matrix was constructed from the corresponding population LD structure. For ADHD, CP, and NDV PGS, posterior effects derived from EUR GWAS data and accounting for EUR LD structure were summed across AFR and AMR individuals’ genotypes from ABCD. This choice was made to comprehensively assess polygenic pathways across a diverse sample given the richness of representation in ABCD; however, these results are to be interpreted cautiously due to weak portability of EUR GWAS to non-EUR samples^110^. As such, our primary results are reported for those of EUR ancestry.

NDV scores were derived as previously described^20^. In brief, using genomicSEM^134^, SNPs were regressed onto a latent factor representing shared genetic architecture among ADHD, ASD, TS, and MDD risk. PRS-CS was applied to the resultant SNP effects on the common factor in the same way as described above and used to generate PGS in ABCD representing individual-level polygenic load for the latent NDV factor. Of note, all GWAS summary statistics included in the calculation of NDV were derived from EUR samples and thus NDV SNP effects are treated as EUR-derived.

#### Partitioned (pathway) PGS

In contrast to classical PGS methods which evaluate SNP effects across the whole genome, newer approaches allow for interrogation of partitions of the genome that share characteristics. These shared features can be functional in nature (e.g., partitioning the genome by genes involved in a specific biological pathway^109^ or developmental (e.g., partitioning by genes that exhibit unique spatiotemporal expression patterns^20^. In the current study, the genome was partitioned into 18 functionally connected gene sets which were defined by their co-expression across development, as defined previously^63^. In brief, Forsyth et al. used weighted-gene coexpression network analysis (WGCNA) to identify spatiotemporally coexpressed genes using gene expression data from timepoints spanning 6 weeks post-conception to 30 years of age across brain regions including 11 cortical regions, cerebellar cortex, and multiple subcortical regions (BrainSpan atlas^64^). Thus genes that cluster within the same module tend to show similar expression patterns throughout the brain across development (see Supplement). For each set of posterior SNP effects generated from PRS-CS, SNPs were partitioned by their position relative to genes in each of the 18 modules defined in Forsyth et al. For a SNP to be considered within a gene range, it must have resided within a window of 35 kb upstream and 10 kb downstream from the gene^20,135^. SNP-to-gene annotations to convert between SNPs and genes were generated with MAGMA v1.08^136^. Thus, each set of posterior effects (1 per GWAS) were partitioned into one of 18 modules and summed across ABCD genotypes (see **Supplementary Methods, section 5** for more detail). Resultant PGS represented polygenic effects of SNPs that map onto clusters of genes that show unique co-expression patterns across brain development weighted by effects on a psychiatric phenotype (e.g., SCZ). These types of PGS are referred to as partitioned or pathway PGS (pPGS)^20,109^, and are referred to hereafter as module pPGS or M*n* pPGS where *n* equals module number. Because the gene modules contain varying numbers of genes (and thus SNPs), a series of permutation tests were conducted to determine whether observed signal from any module pPGS was due to intrinsic characteristics of the genes that make up the module or simply due to the size of the module. These tests were conducted only for pPGS that showed significant associations with the outcome variable (PLEs) before multiple comparison correction. For each tested module pPGS, 10,000 PGS were permuted, which each consisted of a random selection of SNPs with *n_SNP_* equal to the number of SNPs in a module. For example, ADHD EUR M2 pPGS consisted of 14,729 SNPs and thus each of 10k permuted pPGS represented the sum of 14,729 random SNP effects sampled without replacement from the list of whole-genome SNPs. PLE scores were then regressed onto the permuted pPGS, the beta coefficient estimates from which collectively generated a null distribution to compare to the observed effect. One-sided p-values could then be calculated as the proportion of permuted effects that are greater than the observed effect (beta coefficient). Additionally, competitive p-value tests were conducted which calculate a p-value that represents the proportion of permuted p-values that fall below the observed p-value. Primary interpretations relied on one-sided permuted p-values (see **Supplementary Methods, section 5b** for details). To compare PRS-CS-based partitioned results to the analogous clumping and thresholding method, PRSet^109^ was used with default parameters to generate 18 modular pPGS weighted by risk for ADHD (see **Supplementary Methods, section 5c**). We chose to compare methods using ADHD PGS because these pPGS showed the most robust associations with PQ-BC compared to other pPGS.

### Section 6: Statistical analyses

Associations were conducted via linear mixed effects models (LMEs), using R version 4.3.1 *lme4* package (R Foundation for Statistical Computing, Vienna, Austria), with research site, family unit (nested within site, to account for 1336 twins and 23 triplets), and subject ID (nested within-site and family unit) as random effects. In PGS analyses, age, sex, and the top 5 within-ancestry PCs were included as fixed effects covariates; analyses including functional connectivity (without PGS) omitted ancestral PCs and included mean framewise displacement (FD) as an additional covariate. Associations between PGS and FC were tested if the given FC metric was robustly associated with PLEs in analyses involving rsMRI scans recommended by ABCD and Chen et al.^65^. Longitudinal analyses investigating the dynamic relationship between PGS and PLEs and IIV over time included the interaction of PGS with time (PGS x Time). These interaction models were tested only for PGS that showed significant associations with PLE in main models (i.e., ADHD, CP, NDV, and SCZ PGS in EUR participants). False Discovery Rate (FDR) correction was used to correct significance values for multiple comparisons and reduce the probability of Type I error. Main PGS analyses (**Table 2**) corrected for 30 tests (5 PGS x 3 ancestry groups x 2 dependent variables). Module pPGS-PLE analyses (**Figure 4, Supplementary** Figure 3) corrected for 216 tests (18 modules x 4 PGS x 3 ancestry groups). Marginal and conditional R^2^’s are reported for main models and can be interpreted respectively as the proportion of variance in the dependent variable that is explained by the fixed effects only and the fixed and random effects together. A large proportion of the variance is explained by the covariates in most models reported. These R^2^’s are to be distinguished from delta R^2^’s, which represent the additional variance explained by including a term of interest, typically the main independent variable. Although not reported directly, delta R^2^’s for the predictors of interest are directly proportional to, i.e., the square of the reported standardized coefficients. Of note, the PQ-BC data are zero-inflated and positively skewed; however, main analyses conducted with zero-inflated gamma distribution general linear models (hurdle models; glmmTB package) yielded results that were largely consistent with LMEs^137^ (**Supplementary Tables 14-19**). To facilitate intuitive interpretations, and per similar studies^38^, results reported reflect estimates from LMEs.

Mediation models were conducted for relationships for which all paths were significant on their own (X→Y, X→M, M→Y). Using the mediation package in R, we evaluated whether either IIV (not FC metrics as they did not exhibit X→Y) mediated associations between PGS and psychotic-like symptoms, on average across ages 9-15. Confidence intervals were estimated by a quasi-Bayesian bootstrap approach with 10,000 random samples. Because no available packages in R can conduct two-or three-level mediation models, we modeled site, family ID, age, sex, and the top 5 within-ancestry PCs as fixed-effect covariates and modeled subject ID as a random effect.

## Acknowledgements

The ABCD Study is supported by the National Institutes of Health and additional federal partners via the following awards: U01DA041048, U01DA050989, U01DA051016, U01DA041022, U01DA051018, U01DA051037, U01DA050987, U01DA041174, U01DA041106, U01DA041117, U01DA041028, U01DA041134, U01DA050988, U01DA051039, U01DA041156, U01DA041025, U01DA041120, U01DA051038, U01DA041148, U01DA041093, U01DA041089, U24DA041123 and U24DA041147. A full list of federal supporters is available at https://abcdstudy.org/federal-partners.html. Participating study sites and site principal investigators can be found at https://abcdstudy.org/consortium_members/. We thank the investigators and staff at the ABCD sites and coordinating centers, as well as study participants and their families, for their essential contributions to this work. This manuscript reflects the views of the authors and may not reflect the opinions or views of the NIH or ABCD consortium investigators. SEC is supported by the Achievement Rewards for College Scientists. This work was supported by the National Institute of Mental Health (K23 MH121792-01 to NRK, K01 1K01MH135289-01 to LMH, R01MH124694 to JLR, K08MH118577 to JKF, R01MH129858 to CEB). We thank Olivia Wootton for providing the summary statistics from the genome-wide association study on intra-individual variability in reaction time, which was conducted with data from the UK Biobank. We also thank the members of the Bearden Lab and Marcelo Francia for their feedback and support.

## Data Availability

The ABCD measures used in this article can be found at https://doi.org/10.15154/j8kt-v104. The GWAS summary statistics for major psychiatric disorders are available through the Psychiatric Genomics Consortium site: https://pgc.unc.edu/for-researchers/download-results/. Those for cognitive performance are available through the Social Science Genetic Association Consortium site: https://www.thessgac.org/.

## Code Availability

All relevant code is available at https://github.com/hughesdy/ABCD-Attention-PLE

## Author Contributions

Conceptualization was done by SEC, DEH, and CEB. Data processing was performed by SEC and DEH. Formal analysis was carried out by SEC and DEH. Code contributions were made by SEC, DEH, and JZ. Manuscript preparation was performed by SEC and DEH. Review and editing was carried out by all co-authors. Supervision was provided by CEB.

## References

1. Reichenberg, A. (2010). The assessment of neuropsychological functioning in schizophrenia. Dialogues Clin. Neurosci., 12(3), 383–392.

2. Fett, A.-K. J., Reichenberg, A., & Velthorst, E. (2022). Lifespan evolution of neurocognitive impairment in schizophrenia—A narrative review. Schizophr Res Cogn, 28, 100237.

3. Seidman, L. J., Shapiro, D. I., Stone, W. S., Woodberry, K. A., Ronzio, A., Cornblatt, B. A., Addington, J., Bearden, C. E., Cadenhead, K. S., Cannon, T. D., Mathalon, D. H., McGlashan, T. H., Perkins, D. O., Tsuang, M. T., Walker, E. F., & Woods, S. W. (2016). Association of Neurocognition With Transition to Psychosis: Baseline Functioning in the Second Phase of the North American Prodrome Longitudinal Study. JAMA Psychiatry, 73(12), 1239–1248.

4. Cornblatt, B. A., & Erlenmeyer-Kimling, L. (1985). Global attentional deviance as a marker of risk for schizophrenia: Specificity and predictive validity. J. Abnorm. Psychol., 94(4), 470–486.

5. Cornblatt, B., Obuchowski, M., Roberts, S., Pollack, S., & Erlenmeyer-Kimling, L. (1999). Cognitive and behavioral precursors of schizophrenia. Dev. Psychopathol., 11(3), 487–508.

6. Wallace, S., & Linscott, R. J. (2018). Intra-individual variability and psychotic-like experiences in adolescents: Findings from the ALSPAC cohort. Schizophr. Res., 195, 154–159.

7. Sheffield, J. M., Karcher, N. R., & Barch, D. M. (2018). Cognitive Deficits in Psychotic Disorders: A Lifespan Perspective. Neuropsychol. Rev., 28(4), 509–533.

8. Mollon, J., David, A. S., Zammit, S., Lewis, G., & Reichenberg, A. (2018). Course of Cognitive Development From Infancy to Early Adulthood in the Psychosis Spectrum. JAMA Psychiatry, 75(3), 270–279.

9. Uhlhaas, P. J., & Singer, W. (2015). Oscillations and neuronal dynamics in schizophrenia: The search for basic symptoms and translational opportunities. Biol. Psychiatry, 77(12), 1001–1009.

10. Egan, M. F., Goldberg, T. E., Gscheidle, T., Weirich, M., Bigelow, L. B., & Weinberger, D. R. (2000). Relative risk of attention deficits in siblings of patients with schizophrenia. Am. J. Psychiatry, 157(8), 1309–1316.

11. Erlenmeyer-Kimling, L., Rock, D., Roberts, S. A., Janal, M., Kestenbaum, C., Cornblatt, B., Adamo, U. H., & Gottesman, I. I. (2000). Attention, memory, and motor skills as childhood predictors of schizophrenia-related psychoses: The New York High-Risk Project. Am. J. Psychiatry, 157(9), 1416–1422.

12. Cosgrove, D., Mothersill, O., Kendall, K., Konte, B., Harold, D., Giegling, I., Hartmann, A., Richards, A., Mantripragada, K., Owen, M. J., O’Donovan, M. C., Gill, M., Rujescu, D., Walters, J., Corvin, A., Morris, D. W., & Donohoe, G. (2017). Cognitive Characterization of Schizophrenia Risk Variants Involved in Synaptic Transmission: Evidence of CACNA1C’s Role in Working Memory. Neuropsychopharmacology, 42(13), 2612–2622.

13. Nicodemus, K.K., Hargreaves, A., Morris, D., Anney, R., Gill, M., Corvin, A., Donohoe, G., Schizophrenia Psychiatric Genome-wide Association Study Consortium and The Wellcome Trust Case Control Consortium 2. (2014). Variability in working memory performance explained by epistasis vs polygenic scores in the ZNF804A pathway. JAMA Psychiatry.

14. Olde Loohuis, L. M., Mennigen, E., Ori, A. P. S., Perkins, D., Robinson, E., Addington, J., Cadenhead, K. S., Cornblatt, B. A., Mathalon, D. H., McGlashan, T. H., Seidman, L. J., Keshavan, M. S., Stone, W. S., Tsuang, M. T., Walker, E. F., Woods, S. W., Cannon, T. D., Gur, R. C., Gur, R. E., … Ophoff, R. A. (2021). Genetic and clinical analyses of psychosis spectrum symptoms in a large multiethnic youth cohort reveal significant link with ADHD. Transl. Psychiatry, 11(1), 80.

15. Cornblatt, B. A., & Malhotra, A. K. (2001). Impaired attention as an endophenotype for molecular genetic studies of schizophrenia. Am. J. Med. Genet., 105(1), 11–15.

16. Weinberger, D. R. (1987). Implications of normal brain development for the pathogenesis of schizophrenia. Arch. Gen. Psychiatry, 44(7), 660–669.

17. Murray, R. M., & Lewis, S. W. (1987). Is schizophrenia a neurodevelopmental disorder? Br. Med. J., 295(6600), 681–682.

18. Cannon, T. D., Kaprio, J., Lönnqvist, J., Huttunen, M., & Koskenvuo, M. (1998). The genetic epidemiology of schizophrenia in a Finnish twin cohort. A population-based modeling study. Arch. Gen. Psychiatry, 55(1), 67–74.

19. Trubetskoy, V., Pardiñas, A. F., Qi, T., Panagiotaropoulou, G., Awasthi, S., Bigdeli, T. B., Bryois, J., Chen, C.-Y., Dennison, C. A., Hall, L. S., Lam, M., Watanabe, K., Frei, O., Ge, T., Harwood, J. C., Koopmans, F., Magnusson, S., Richards, A. L., Sidorenko, J., … Schizophrenia Working Group of the Psychiatric Genomics Consortium. (2022). Mapping genomic loci implicates genes and synaptic biology in schizophrenia. Nature, 604(7906), 502–508.

20. Hughes, D. E., Kunitoki, K., Elyounssi, S., Luo, M., Bazer, O. M., Hopkinson, C. E., Dowling, K. F., Doyle, A. E., Dunn, E. C., Eryilmaz, H., Gilman, J. M., Holt, D. J., Valera, E. M., Smoller, J. W., Cecil, C. A. M., Tiemeier, H., Lee, P. H., & Roffman, J. L. (2023). Genetic patterning for child psychopathology is distinct from that for adults and implicates fetal cerebellar development. Nat. Neurosci., 26(6), 959–969.

21. Jones, H. J., Stergiakouli, E., Tansey, K. E., Hubbard, L., Heron, J., Cannon, M., Holmans, P., Lewis, G., Linden, D. E. J., Jones, P. B., Davey Smith, G., O’Donovan, M. C., Owen, M. J., Walters, J. T., & Zammit, S. (2016). Phenotypic Manifestation of Genetic Risk for Schizophrenia During Adolescence in the General Population. JAMA Psychiatry, 73(3), 221–228.

22. Zammit, S., Hamshere, M., Dwyer, S., Georgiva, L., Timpson, N., Moskvina, V., Richards, A., Evans, D. M., Lewis, G., Jones, P., Owen, M. J., & O’Donovan, M. C. (2014). A population-based study of genetic variation and psychotic experiences in adolescents. Schizophr. Bull., 40(6), 1254–1262.

23. Nivard, M. G., Gage, S. H., Hottenga, J. J., Van Beijsterveldt, C. E. M., Abdellaoui, A., Bartels, M., Baselmans, B. M. L., Ligthart, L., Pourcain, B. S., Boomsma, D. I., & Others. (2017). Genetic overlap between schizophrenia and developmental psychopathology: Longitudinal and multivariate polygenic risk prediction of common psychiatric traits during development. Schizophr. Bull., 43(6), 1197–1207.

24. Hatzimanolis, A., Bhatnagar, P., Moes, A., Wang, R., Roussos, P., Bitsios, P., Stefanis, C. N., Pulver, A. E., Arking, D. E., Smyrnis, N., Stefanis, N. C., & Avramopoulos, D. (2015). Common genetic variation and schizophrenia polygenic risk influence neurocognitive performance in young adulthood. Am. J. Med. Genet. B Neuropsychiatr. Genet., 168B(5), 392–401.

25. Moreau, C. A., Harvey, A., Kumar, K., Huguet, G., Urchs, S. G. W., Douard, E. A., Schultz, L. M., Sharmarke, H., Jizi, K., Martin, C.-O., Younis, N., Tamer, P., Rolland, T., Martineau, J.-L., Orban, P., Silva, A. I., Hall, J., van den Bree, M. B. M., Owen, M. J., … Jacquemont, S. (2023). Genetic Heterogeneity Shapes Brain Connectivity in Psychiatry. Biol. Psychiatry, 93(1), 45–58.

26. Cao, H., Zhou, H., & Cannon, T. D. (2021). Functional connectome-wide associations of schizophrenia polygenic risk. Mol. Psychiatry, 26(6), 2553–2561.

27. Chidharom, M., Krieg, J., & Bonnefond, A. (2021). Impaired Frontal Midline Theta During Periods of High Reaction Time Variability in Schizophrenia. Biol Psychiatry Cogn Neurosci Neuroimaging, 6(4), 429–438.

28. Panagiotaropoulou, G., Thrapsanioti, E., Pappa, E., Grigoras, C., Mylonas, D., Karavasilis, E., Velonakis, G., Kelekis, N., & Smyrnis, N. (2019). Hypo-activity of the dorsolateral prefrontal cortex relates to increased reaction time variability in patients with schizophrenia. Neuroimage Clin, 23, 101853.

29. Shin, Y. S., Kim, S. N., Shin, N. Y., Jung, W. H., Hur, J.-W., Byun, M. S., Jang, J. H., An, S. K., & Kwon, J. S. (2013). Increased intra-individual variability of cognitive processing in subjects at risk mental state and schizophrenia patients. PLoS One, 8(11), e78354.

30. Hilti, C. C., Hilti, L. M., Heinemann, D., Robbins, T., Seifritz, E., & Cattapan-Ludewig, K. (2010). Impaired performance on the Rapid Visual Information Processing task (RVIP) could be an endophenotype of schizophrenia. Psychiatry Res., 177(1–2), 60–64.

31. Crosbie, J., Arnold, P., Paterson, A., Swanson, J., Dupuis, A., Li, X., Shan, J., Goodale, T., Tam, C., Strug, L. J., & Schachar, R. J. (2013). Response inhibition and ADHD traits: Correlates and heritability in a community sample. J. Abnorm. Child Psychol., 41(3), 497–507.

32. Kuntsi, J., Frazier-Wood, A. C., Banaschewski, T., Gill, M., Miranda, A., Oades, R. D., Roeyers, H., Rothenberger, A., Steinhausen, H.-C., van der Meere, J. J., Faraone, S. V., Asherson, P., & Rijsdijk, F. (2013). Genetic analysis of reaction time variability: Room for improvement? Psychol. Med., 43(6), 1323–1333.

33. Roalf, D. R., Gur, R. C., Almasy, L., Richard, J., Gallagher, R. S., Prasad, K., Wood, J., Pogue-Geile, M. F., Nimgaonkar, V. L., & Gur, R. E. (2013). Neurocognitive performance stability in a multiplex multigenerational study of schizophrenia. Schizophr. Bull., 39(5), 1008–1017.

34. Wootton, O., Dalvie, S., Susser, E., Gur, R. C., & Stein, D. J. (2023). Within-individual variability in cognitive performance in schizophrenia: A narrative review of the key literature and proposed research agenda. Schizophr. Res., 252, 329–334.

35. Kelly, A. M. C., Uddin, L. Q., Biswal, B. B., Castellanos, F. X., & Milham, M. P. (2008). Competition between functional brain networks mediates behavioral variability. Neuroimage, 39(1), 527–537.

36. Chang, S. E., Lenartowicz, A., Hellemann, G. S., Uddin, L. Q., & Bearden, C. E. (2023). Variability in Cognitive Task Performance in Early Adolescence Is Associated With Stronger Between-Network Anticorrelation and Future Attention Problems. Biol Psychiatry Glob Open Sci, 3(4), 948–957.

37. Fox, M. D., Snyder, A. Z., Vincent, J. L., Corbetta, M., Van Essen, D. C., & Raichle, M. E. (2005). The human brain is intrinsically organized into dynamic, anticorrelated functional networks. Proc. Natl. Acad. Sci. U. S. A., 102(27), 9673–9678.

38. Karcher, N. R., O’Brien, K. J., Kandala, S., & Barch, D. M. (2019). Resting-State Functional Connectivity and Psychotic-like Experiences in Childhood: Results From the Adolescent Brain Cognitive Development Study. Biol. Psychiatry, 86(1), 7–15.

39. Chai, X. J., Whitfield-Gabrieli, S., Shinn, A. K., Gabrieli, J. D. E., Nieto Castañón, A., McCarthy, J. M., Cohen, B. M., & Ongür, D. (2011). Abnormal medial prefrontal cortex resting-state connectivity in bipolar disorder and schizophrenia.

40. Whitfield-Gabrieli, S., Thermenos, H. W., Milanovic, S., Tsuang, M. T., Faraone, S. V., McCarley, R. W., Shenton, M. E., Green, A. I., Nieto-Castanon, A., LaViolette, P., Wojcik, J., Gabrieli, J. D. E., & Seidman, L. J. (2009). Hyperactivity and hyperconnectivity of the default network in schizophrenia and in first-degree relatives of persons with schizophrenia. Proc. Natl. Acad. Sci. U. S. A., 106(4), 1279–1284.

41. Adhikari, B. M., Hong, L. E., Sampath, H., Chiappelli, J., Jahanshad, N., Thompson, P. M., Rowland, L. M., Calhoun, V. D., Du, X., Chen, S., & Kochunov, P. (2019). Functional network connectivity impairments and core cognitive deficits in schizophrenia. Hum. Brain Mapp., 40(16), 4593–4605.

42. Sheffield, J. M., Karcher, N. R., & Barch, D. M. (2018). Cognitive Deficits in Psychotic Disorders: A Lifespan Perspective. Neuropsychol. Rev., 28(4), 509–533.

43. Teeuw, J., Brouwer, R. M., Guimarães, J. P. O. F. T., Brandner, P., Koenis, M. M. G., Swagerman, S. C., Verwoert, M., Boomsma, D. I., & Hulshoff Pol, H. E. (2019). Genetic and environmental influences on functional connectivity within and between canonical cortical resting-state networks throughout adolescent development in boys and girls. Neuroimage, 202, 116073.

44. Cao, H., Dixson, L., Meyer-Lindenberg, A., & Tost, H. (2016). Functional connectivity measures as schizophrenia intermediate phenotypes: Advances, limitations, and future directions. Curr. Opin. Neurobiol., 36, 7–14.

45. Meyer-Lindenberg, A. (2009). Neural connectivity as an intermediate phenotype: Brain networks under genetic control. Hum. Brain Mapp., 30(7), 1938–1946.

46. MacDonald, S. W. S., Nyberg, L., & Bäckman, L. (2006). Intra-individual variability in behavior: Links to brain structure, neurotransmission and neuronal activity. Trends Neurosci., 29(8), 474– 480.

47. Stevens, M. C. (2016). The contributions of resting state and task-based functional connectivity studies to our understanding of adolescent brain network maturation. Neurosci. Biobehav. Rev., 70, 13–32.

48. Crone, E. A., & Dahl, R. E. (2012). Understanding adolescence as a period of social–affective engagement and goal flexibility. Nat. Rev. Neurosci., 13(9), 636–650.

49. Insel, T. R. (2010). Rethinking schizophrenia. Nature, 468(7321), 187–193.

50. Keller, A. S., Sydnor, V. J., Pines, A., Fair, D. A., Bassett, D. S., & Satterthwaite, T. D. (2023). Hierarchical functional system development supports executive function. Trends Cogn. Sci., 27(2), 160–174.

51. Sydnor, V. J., Larsen, B., Bassett, D. S., Alexander-Bloch, A., Fair, D. A., Liston, C., Mackey, A. P., Milham, M. P., Pines, A., Roalf, D. R., Seidlitz, J., Xu, T., Raznahan, A., & Satterthwaite, T. D. (2021). Neurodevelopment of the association cortices: Patterns, mechanisms, and implications for psychopathology. Neuron, 109(18), 2820–2846.

52. Pines, A. R., Larsen, B., Cui, Z., Sydnor, V. J., Bertolero, M. A., Adebimpe, A., Alexander-Bloch, A. F., Davatzikos, C., Fair, D. A., Gur, R. C., Gur, R. E., Li, H., Milham, M. P., Moore, T. M., Murtha, K., Parkes, L., Thompson-Schill, S. L., Shanmugan, S., Shinohara, R. T., … Satterthwaite, T. D. (2022). Dissociable multi-scale patterns of development in personalized brain networks. Nat. Commun., 13(1), 2647.

53. Fair, D. A., Dosenbach, N. U. F., Church, J. A., Cohen, A. L., Brahmbhatt, S., Miezin, F. M., Barch, D. M., Raichle, M. E., Petersen, S. E., & Schlaggar, B. L. (2007). Development of distinct control networks through segregation and integration. Proc. Natl. Acad. Sci. U. S. A., 104(33), 13507–13512.

54. Wang, C., Hu, Y., Weng, J., Chen, F., & Liu, H. (2020). Modular segregation of task-dependent brain networks contributes to the development of executive function in children. Neuroimage, 206, 116334.

55. Chai, X. J., Ofen, N., Gabrieli, J. D. E., & others. (2014). Selective development of anticorrelated networks in the intrinsic functional organization of the human brain. Journal of Cognitive.

56. Whitfield-Gabrieli, S., Wendelken, C., Nieto-Castañón, A., Bailey, S. K., Anteraper, S. A., Lee, Y. J., Chai, X.-Q., Hirshfeld-Becker, D. R., Biederman, J., Cutting, L. E., & Bunge, S. A. (2020). Association of Intrinsic Brain Architecture With Changes in Attentional and Mood Symptoms During Development. JAMA Psychiatry, 77(4), 378–386.

57. Price, A. J., Jaffe, A. E., & Weinberger, D. R. (2021). Cortical cellular diversity and development in schizophrenia. Mol. Psychiatry, 26(1), 203–217.

58. Forsyth, J. K., & Lewis, D. A. (2017). Mapping the Consequences of Impaired Synaptic Plasticity in Schizophrenia through Development: An Integrative Model for Diverse Clinical Features. Trends Cogn. Sci., 21(10), 760–778.

59. Gulsuner, S., Walsh, T., Watts, A. C., Lee, M. K., Thornton, A. M., Casadei, S., Rippey, C., Shahin, H., Consortium on the Genetics of Schizophrenia (COGS), PAARTNERS Study Group, Nimgaonkar, V. L., Go, R. C. P., Savage, R. M., Swerdlow, N. R., Gur, R. E., Braff, D. L., King, M.-C., & McClellan, J. M. (2013). Spatial and temporal mapping of de novo mutations in schizophrenia to a fetal prefrontal cortical network. Cell, 154(3), 518–529.

60. Weinberger, D. R., & Harrison, P. J. (Eds.). (2011). Schizophrenia: Weinberger/Schizophrenia (3rd ed.). Wiley-Blackwell.

61. Casey, B. J., Cannonier, T., Conley, M. I., Cohen, A. O., Barch, D. M., Heitzeg, M. M., Soules, M. E., Teslovich, T., Dellarco, D. V., Garavan, H., Orr, C. A., Wager, T. D., Banich, M. T., Speer, N. K., Sutherland, M. T., Riedel, M. C., Dick, A. S., Bjork, J. M., Thomas, K. M., … ABCD Imaging Acquisition Workgroup. (2018). The Adolescent Brain Cognitive Development (ABCD) study: Imaging acquisition across 21 sites. Dev. Cogn. Neurosci., 32, 43–54.

62. Lewis, D. A., & Levitt, P. (2002). Schizophrenia as a disorder of neurodevelopment. Annu. Rev. Neurosci., 25, 409–432.

63. Forsyth, J. K., Nachun, D., Gandal, M. J., Geschwind, D. H., Anderson, A. E., Coppola, G., & Bearden, C. E. (2020). Synaptic and Gene Regulatory Mechanisms in Schizophrenia, Autism, and 22q11.2 Copy Number Variant–Mediated Risk for Neuropsychiatric Disorders. Biol. Psychiatry, 87(2), 150–163.

64. Kang, H. J., Kawasawa, Y. I., Cheng, F., Zhu, Y., Xu, X., Li, M., Sousa, A. M. M., Pletikos, M., Meyer, K. A., Sedmak, G., Guennel, T., Shin, Y., Johnson, M. B., Krsnik, Z., Mayer, S., Fertuzinhos, S., Umlauf, S., Lisgo, S. N., Vortmeyer, A., … Sestan, N. (2011).

65. Chen, J., Tam, A., Kebets, V., Orban, C., Ooi, L. Q. R., Asplund, C. L., Marek, S., Dosenbach, N. U. F., Eickhoff, S. B., Bzdok, D., Holmes, A. J., & Yeo, B. T. T. (2022). Shared and unique brain network features predict cognitive, personality, and mental health scores in the ABCD study. Nat. Commun., 13(1), 2217.

66. Legge, S. E., Cardno, A. G., Allardyce, J., Dennison, C., Hubbard, L., Pardiñas, A. F., Richards, A., Rees, E., Di Florio, A., Escott-Price, V., Zammit, S., Holmans, P., Owen, M. J., O’Donovan, M. C., & Walters, J. T. R. (2021). Associations Between Schizophrenia Polygenic Liability, Symptom Dimensions, and Cognitive Ability in Schizophrenia. JAMA Psychiatry, 78(10), 1143– 1151.

67. Pain, O., Dudbridge, F., Cardno, A. G., Freeman, D., Lu, Y., Lundstrom, S., Lichtenstein, P., & Ronald, A. (2018). Genome-wide analysis of adolescent psychotic-like experiences shows genetic overlap with psychiatric disorders. Am. J. Med. Genet. B Neuropsychiatr. Genet., 177(4), 416– 425.

68. Karcher, N. R., Paul, S. E., Johnson, E. C., Hatoum, A. S., Baranger, D. A. A., Agrawal, A., Thompson, W. K., Barch, D. M., & Bogdan, R. (2022). Psychotic-like Experiences and Polygenic Liability in the Adolescent Brain Cognitive Development Study. Biol Psychiatry Cogn Neurosci Neuroimaging, 7(1), 45–55.

69. Pardiñas, A. F., Holmans, P., Pocklington, A. J., Escott-Price, V., Ripke, S., Carrera, N., Legge, S. E., Bishop, S., Cameron, D., Hamshere, M. L., Han, J., Hubbard, L., Lynham, A., Mantripragada, K., Rees, E., MacCabe, J. H., McCarroll, S. A., Baune, B. T., Breen, G., … Walters, J. T. R. (2018). Common schizophrenia alleles are enriched in mutation-intolerant genes and in regions under strong background selection. Nat. Genet., 50(3), 381–389.

70. Bulik-Sullivan, B., Finucane, H. K., Anttila, V., Gusev, A., Day, F. R., Loh, P.-R., ReproGen Consortium, Psychiatric Genomics Consortium, Genetic Consortium for Anorexia Nervosa of the Wellcome Trust Case Control Consortium 3, Duncan, L., Perry, J. R. B., Patterson, N., Robinson, E. B., Daly, M. J., Price, A. L., & Neale, B. M. (2015). An atlas of genetic correlations across human diseases and traits. Nat. Genet., 47(11), 1236–1241.

71. Addington, J., Piskulic, D., Liu, L., Lockwood, J., Cadenhead, K. S., Cannon, T. D., Cornblatt, B. A., McGlashan, T. H., Perkins, D. O., Seidman, L. J., Tsuang, M. T., Walker, E. F., Bearden, C. E., Mathalon, D. H., & Woods, S. W. (2017). Comorbid diagnoses for youth at clinical high risk of psychosis. Schizophr. Res., 190, 90–95.

72. Hubbard, L., Tansey, K. E., Rai, D., Jones, P., Ripke, S., Chambert, K. D., Moran, J. L., McCarroll, S. A., Linden, D. E. J., Owen, M. J., O’Donovan, M. C., Walters, J. T. R., & Zammit, S. (2016). Evidence of Common Genetic Overlap Between Schizophrenia and Cognition. Schizophr. Bull., 42(3), 832–842.

73. Escott-Price, V., Bracher-Smith, M., Menzies, G., Walters, J., Kirov, G., Owen, M. J., & O’Donovan, M. C. (2020). Genetic liability to schizophrenia is negatively associated with educational attainment in UK Biobank. Mol. Psychiatry, 25(4), 703–705.

74. Toulopoulou, T., Picchioni, M., Rijsdijk, F., Hua-Hall, M., Ettinger, U., Sham, P., & Murray, R. (2007). Substantial genetic overlap between neurocognition and schizophrenia: Genetic modeling in twin samples. Arch. Gen. Psychiatry, 64(12), 1348–1355.

75. Smeland, O. B., Bahrami, S., Frei, O., Shadrin, A., O’Connell, K., Savage, J., Watanabe, K., Krull, F., Bettella, F., Steen, N. E., Ueland, T., Posthuma, D., Djurovic, S., Dale, A. M., & Andreassen, O. A. (2020). Genome-wide analysis reveals extensive genetic overlap between schizophrenia, bipolar disorder, and intelligence. Mol. Psychiatry, 25(4), 844–853.

76. Sniekers, S., Stringer, S., Watanabe, K., Jansen, P. R., Coleman, J. R. I., Krapohl, E., Taskesen, E., Hammerschlag, A. R., Okbay, A., Zabaneh, D., Amin, N., Breen, G., Cesarini, D., Chabris, C. F., Iacono, W. G., Ikram, M. A., Johannesson, M., Koellinger, P., Lee, J. J., … Posthuma, D. (2017). Genome-wide association meta-analysis of 78,308 individuals identifies new loci and genes influencing human intelligence. Nat. Genet., 49(7), 1107–1112.

77. A. Moses, M., Tiego, J., Demontis, D., Bragi Walters, G., Stefansson, H., Stefansson, K., Børglum, D., Arnatkeviciute, A., & Bellgrove, M. A. (2022). Working memory and reaction time variability mediate the relationship between polygenic risk and ADHD traits in a general population sample. Mol. Psychiatry, 27(12), 5028–5037.

77. Vainieri, I., Martin, J., Rommel, A.-S., Asherson, P., Banaschewski, T., Buitelaar, J., Cormand, B., Crosbie, J., Faraone, S. V., Franke, B., Loo, S. K., Miranda, A., Manor, I., Oades, R. D., Purves, K. L., Ramos-Quiroga, J. A., Ribasés, M., Roeyers, H., Rothenberger, A., … Kuntsi, J. (2022). Polygenic association between attention-deficit/hyperactivity disorder liability and cognitive impairments. Psychol. Med., 52(14), 3150–3158.

78. Plomin, R. (1999). Genetics and general cognitive ability. Nature, 402(6761 Suppl), C25-9.

79. Bora, E., Lin, A., Wood, S. J., Yung, A. R., McGorry, P. D., & Pantelis, C. (2014). Cognitive deficits in youth with familial and clinical high risk to psychosis: A systematic review and meta-analysis. Acta Psychiatr. Scand., 130(1), 1–15.

80. Blokland, G. A. M., Mesholam-Gately, R. I., Toulopoulou, T., Del Re, E. C., Lam, M., DeLisi, L. E., Donohoe, G., Walters, J. T. R., GENUS Consortium, Seidman, L. J., & Petryshen, T. L. (2017). Heritability of Neuropsychological Measures in Schizophrenia and Nonpsychiatric Populations: A Systematic Review and Meta-analysis. Schizophr. Bull., 43(4), 788–800.

81. Davies, G., Lam, M., Harris, S. E., Trampush, J. W., Luciano, M., Hill, W. D., Hagenaars, S. P., Ritchie, S. J., Marioni, R. E., Fawns-Ritchie, C., Liewald, D. C. M., Okely, J. A., Ahola-Olli, A. V., Barnes, C. L. K., Bertram, L., Bis, J. C., Burdick, K. E., Christoforou, A., DeRosse, P., … Deary, I. J. (2018). Study of 300,486 individuals identifies 148 independent genetic loci influencing general cognitive function. Nat. Commun., 10(1), 2068.

82. Lencz, T., Knowles, E., Davies, G., Guha, S., Liewald, D. C., Starr, J. M., Djurovic, S., Melle, I., Sundet, K., Christoforou, A., Reinvang, I., Mukherjee, S., DeRosse, P., Lundervold, A., Steen, V. M., John, M., Espeseth, T., Räikkönen, K., Widen, E., … Malhotra, A. K. (2014). Molecular genetic evidence for overlap between general cognitive ability and risk for schizophrenia: A report from the Cognitive Genomics consorTium (COGENT). Mol. Psychiatry, 19(2), 168–174.

83. Hagenaars, S. P., Harris, S. E., Davies, G., Hill, W. D., Liewald, D. C. M., Ritchie, S. J., Marioni, R. E., Fawns-Ritchie, C., Cullen, B., Malik, R., Worrall, B. B., Sudlow, C. L. M., Wardlaw, J. M., Gallacher, J., Pell, J., McIntosh, A. M., Smith, D. J., Gale, C. R., & Deary, I. J. (2016). Shared genetic aetiology between cognitive functions and physical and mental health in UK Biobank (N=112 151) and 24 GWAS consortia. Mol. Psychiatry, 21(11), 1624–1632.

84. Germine, L., Robinson, E. B., Smoller, J. W., Calkins, M. E., Moore, T. M., Hakonarson, H., Daly, M. J., Lee, P. H., Holmes, A. J., Buckner, R. L., Gur, R. C., & Gur, R. E. (2016). Association between polygenic risk for schizophrenia, neurocognition and social cognition across development. Transl. Psychiatry, 6(10), e924.

85. Liebers, D. T., Pirooznia, M., Seiffudin, F., Musliner, K. L., Zandi, P. P., & Goes, F. S. (2016). Polygenic Risk of Schizophrenia and Cognition in a Population-Based Survey of Older Adults. Schizophr. Bull., 42(4), 984–991.

86. Trampush, J. W., Yang, M. L. Z., Yu, J., Knowles, E., Davies, G., Liewald, D. C., Starr, J. M., Djurovic, S., Melle, I., Sundet, K., Christoforou, A., Reinvang, I., DeRosse, P., Lundervold, A. J., Steen, V. M., Espeseth, T., Räikkönen, K., Widen, E., Palotie, A., … Lencz, T. (2017). GWAS meta-analysis reveals novel loci and genetic correlates for general cognitive function: A report from the COGENT consortium. Mol. Psychiatry, 22(11), 1651–1652.

87. Riglin, L., Collishaw, S., Richards, A., Thapar, A. K., Maughan, B., O’Donovan, M. C., & Thapar, A. (2017). Schizophrenia risk alleles and neurodevelopmental outcomes in childhood: A population-based cohort study. Lancet Psychiatry, 4(1), 57–62.

88. Kendler, K. S., Ohlsson, H., Sundquist, J., & Sundquist, K. (2015). IQ and Schizophrenia in a Swedish National Sample: Their Causal Relationship and the Interaction of IQ With Genetic Risk. AJP, 172(3), 259–265.

89. He, Q., Jantac Mam-Lam-Fook, C., Chaignaud, J., Danset-Alexandre, C., Iftimovici, A., Gradels Hauguel, J., Houle, G., Liao, C., ICAAR study group, Dion, P. A., Rouleau, G. A., Kebir, O., Krebs, M.-O., & Chaumette, B. (2021). Influence of polygenic risk scores for schizophrenia and resilience on the cognition of individuals at-risk for psychosis. Transl. Psychiatry, 11(1), 518.

90. Jonas, K. G., Lencz, T., Li, K., Malhotra, A. K., Perlman, G., Fochtmann, L. J., Bromet, E. J., & Kotov, R. (2019). Schizophrenia polygenic risk score and 20-year course of illness in psychotic disorders. Transl. Psychiatry, 9(1), 300.

91. Dickinson, D., Zaidman, S. R., Giangrande, E. J., Eisenberg, D. P., Gregory, M. D., & Berman, K. F. (2020). Distinct Polygenic Score Profiles in Schizophrenia Subgroups With Different Trajectories of Cognitive Development. Am. J. Psychiatry, 177(4), 298–307.

93. Habtewold, T. D., Liemburg, E. J., Islam, M. A., de Zwarte, S. M. C., Boezen, H. M., GROUP Investigators, Bruggeman, R., & Alizadeh, B. Z. (2020). Association of schizophrenia polygenic risk score with data-driven cognitive subtypes: A six-year longitudinal study in patients, siblings and controls. Schizophr. Res., 223, 135–147.

93. Nakahara, S., Medland, S., Turner, J. A., Calhoun, V. D., Lim, K. O., Mueller, B. A., Bustillo, J. R., O’Leary, D. S., Vaidya, J. G., McEwen, S., Voyvodic, J., Belger, A., Mathalon, D. H., Ford, J. M., Guffanti, G., Macciardi, F., Potkin, S. G., & van Erp, T. G. M. (2018). Polygenic risk score, genome-wide association, and gene set analyses of cognitive domain deficits in schizophrenia. Schizophr. Res., 201, 393–399.

94. Toulopoulou, T., Zhang, X., Cherny, S., Dickinson, D., Berman, K. F., Straub, R. E., Sham, P., & Weinberger, D. R. (2019). Polygenic risk score increases schizophrenia liability through cognition-relevant pathways. Brain, 142(2), 471–485.

95. Richards, A. L., Pardiñas, A. F., Frizzati, A., Tansey, K. E., Lynham, A. J., Holmans, P., Legge, S. E., Savage, J. E., Agartz, I., Andreassen, O. A., Blokland, G. A. M., Corvin, A., Cosgrove, D., Degenhardt, F., Djurovic, S., Espeseth, T., Ferraro, L., Gayer-Anderson, C., Giegling, I., … Walters, J. T. R. (2020). The Relationship Between Polygenic Risk Scores and Cognition in Schizophrenia. Schizophr. Bull., 46(2), 336–344.

97. Mallet, J., Le Strat, Y., Dubertret, C., & Gorwood, P. (2020). Polygenic Risk Scores Shed Light on the Relationship between Schizophrenia and Cognitive Functioning: Review and Meta-Analysis. J. Clin. Med. Res., 9(2).

97. Engen, M. J., Lyngstad, S. H., Ueland, T., Simonsen, C. E., Vaskinn, A., Smeland, O., Bettella, F., Lagerberg, T. V., Djurovic, S., Andreassen, O. A., & Others. (2020). Polygenic scores for schizophrenia and general cognitive ability: Associations with six cognitive domains, premorbid intelligence, and cognitive composite score in individuals with a psychotic disorder and in healthy controls. Transl. Psychiatry, 10(1), 416.

98. Shafee, R., Nanda, P., Padmanabhan, J. L., Tandon, N., Alliey-Rodriguez, N., Kalapurakkel, S., Weiner, D. J., Gur, R. E., Keefe, R. S. E., Hill, S. K., Bishop, J. R., Clementz, B. A., Tamminga, C. A., Gershon, E. S., Pearlson, G. D., Keshavan, M. S., Sweeney, J. A., McCarroll, S. A., & Robinson, E. B. (2018). Polygenic risk for schizophrenia and measured domains of cognition in individuals with psychosis and controls. Transl. Psychiatry, 8(1), 78.

99. Iraji, A., Chen, J., Lewis, N., Faghiri, A., Fu, Z., Agcaoglu, O., Kochunov, P., Adhikari, B. M., Mathalon, D. H., Pearlson, G. D., Macciardi, F., Preda, A., van Erp, T. G. M., Bustillo, J. R., Díaz-Caneja, C. M., Andrés-Camazón, P., Dhamala, M., Adali, T., & Calhoun, V. D. (2023). Spatial Dynamic Subspaces Encode Sex-Specific Schizophrenia Disruptions in Transient Network Overlap and its Links to Genetic Risk. Biol. Psychiatry.

100. Moreau, C. A., Harvey, A., Kumar, K., Huguet, G., Urchs, S. G. W., Douard, E. A., Schultz, L. M., Sharmarke, H., Jizi, K., Martin, C.-O., Younis, N., Tamer, P., Rolland, T., Martineau, J.-L., Orban, P., Silva, A. I., Hall, J., van den Bree, M. B. M., Owen, M. J., … Jacquemont, S. (2023). Genetic Heterogeneity Shapes Brain Connectivity in Psychiatry. Biol. Psychiatry, 93(1), 45–58.

101. Qi, S., Sui, J., Pearlson, G., Bustillo, J., Perrone-Bizzozero, N. I., Kochunov, P., Turner, J. A., Fu, Z., Shao, W., Jiang, R., Yang, X., Liu, J., Du, Y., Chen, J., Zhang, D., & Calhoun, V. D. (2022). Derivation and utility of schizophrenia polygenic risk associated multimodal MRI frontotemporal network. Nat. Commun., 13(1), 4929.

102. Wang, T., Zhang, X., Li, A., Zhu, M., Liu, S., Qin, W., Li, J., Yu, C., Jiang, T., & Liu, B. (2017). Polygenic risk for five psychiatric disorders and cross-disorder and disorder-specific neural connectivity in two independent populations. Neuroimage Clin, 14, 441–449.

103. Passiatore, R., Antonucci, L. A., DeRamus, T. P., Fazio, L., Stolfa, G., Sportelli, L., Kikidis, G. C., Blasi, G., Chen, Q., Dukart, J., Goldman, A. L., Mattay, V. S., Popolizio, T., Rampino, A., Sambataro, F., Selvaggi, P., Ulrich, W., Apulian Network on Risk for Psychosis, Weinberger, D. R., … Pergola, G. (2023). Changes in patterns of age-related network connectivity are associated with risk for schizophrenia. Proc. Natl. Acad. Sci. U. S. A., 120(32), e2221533120.

104. Tiego, J., Thompson, K., Arnatkeviciute, A., Hawi, Z., Finlay, A., Sabaroedin, K., Johnson, B., Bellgrove, M. A., & Fornito, A. (2023). Dissecting Schizotypy and Its Association With Cognition and Polygenic Risk for Schizophrenia in a Nonclinical Sample. Schizophr. Bull., 49(5), 1217–1228.

105. Tervo-Clemmens, B., Calabro, F. J., Parr, A. C., Fedor, J., Foran, W., & Luna, B. (2023). A canonical trajectory of executive function maturation from adolescence to adulthood. Nat. Commun., 14(1), 6922.

106. Geschwind, D. H., & Flint, J. (2015). Genetics and genomics of psychiatric disease. Science, 349(6255), 1489–1494.

107. Ding, Y., Hou, K., Xu, Z., Pimplaskar, A., Petter, E., Boulier, K., Privé, F., Vilhjálmsson, B. J., Olde Loohuis, L. M., & Pasaniuc, B. (2023). Polygenic scoring accuracy varies across the genetic ancestry continuum. Nature, 618(7966), 774–781.

108. Choi, S. W., García-González, J., Ruan, Y., Wu, H. M., Porras, C., Johnson, J., Bipolar Disorder Working group of the Psychiatric Genomics Consortium, Hoggart, C. J., & O’Reilly, P. F. (2023). PRSet: Pathway-based polygenic risk score analyses and software. PLoS Genet., 19(2), e1010624.

109. Martin, A. R., Kanai, M., Kamatani, Y., Okada, Y., Neale, B. M., & Daly, M. J. (2019). Clinical use of current polygenic risk scores may exacerbate health disparities. Nat. Genet., 51(4), 584–591.

110. Kachuri, L., Chatterjee, N., Hirbo, J., Schaid, D. J., Martin, I., Kullo, I. J., Kenny, E. E., Pasaniuc, B., Polygenic Risk Methods in Diverse Populations (PRIMED) Consortium Methods Working Group, Witte, J. S., & Ge, T. (2024). Principles and methods for transferring polygenic risk scores across global populations. Nat. Rev. Genet., 25(1), 8–25.

111. Wang, Y., Kanai, M., Tan, T., Kamariza, M., Tsuo, K., Yuan, K., Zhou, W., Okada, Y., BioBank Japan Project, Huang, H., Turley, P., Atkinson, E. G., & Martin, A. R. (2023). Polygenic prediction across populations is influenced by ancestry, genetic architecture, and methodology. Cell Genom, 3(10), 100408.

112. Wray, N. R., Lin, T., Austin, J., McGrath, J. J., Hickie, I. B., Murray, G. K., & Visscher, P. M. (2021). From Basic Science to Clinical Application of Polygenic Risk Scores: A Primer. JAMA Psychiatry, 78(1), 101–109.

113. Hauberg, M. E., Creus-Muncunill, J., Bendl, J., Kozlenkov, A., Zeng, B., Corwin, C., Chowdhury, S., Kranz, H., Hurd, Y. L., Wegner, M., Børglum, A. D., Dracheva, S., Ehrlich, M. E., Fullard, J. F., & Roussos, P. (2020). Common schizophrenia risk variants are enriched in open chromatin regions of human glutamatergic neurons. Nat. Commun., 11(1), 5581.

114. Garavan, H., Bartsch, H., Conway, K., Decastro, A., Goldstein, R. Z., Heeringa, S., Jernigan, T., Potter, A., Thompson, W., & Zahs, D. (2018). Recruiting the ABCD sample: Design considerations and procedures. Dev. Cogn. Neurosci., 32, 16–22.

115. Luciana, M., Bjork, J. M., Nagel, B. J., Barch, D. M., Gonzalez, R., Nixon, S. J., & Banich, M. T. (2018). Adolescent neurocognitive development and impacts of substance use: Overview of the adolescent brain cognitive development (ABCD) baseline neurocognition battery. Dev. Cogn. Neurosci., 32, 67–79.

116. Anokhin, A. P., Luciana, M., Banich, M., Barch, D., Bjork, J. M., Gonzalez, M. R., Gonzalez, R., Haist, F., Jacobus, J., Lisdahl, K., McGlade, E., McCandliss, B., Nagel, B., Nixon, S. J., Tapert, S., Kennedy, J. T., & Thompson, W. (2022). Age-related changes and longitudinal stability of individual differences in ABCD Neurocognition measures. Dev. Cogn. Neurosci., 54, 101078.

117. Leth-Steensen, C., Elbaz, Z. K., & Douglas, V. I. (2000). Mean response times, variability, and skew in the responding of ADHD children: A response time distributional approach. Acta Psychol., 104(2), 167–190.

118. Williams, B. R., Hultsch, D. F., Strauss, E. H., Hunter, M. A., & Tannock, R. (2005). Inconsistency in reaction time across the life span. Neuropsychology, 19(1), 88–96

119. Karcher, N. R., Barch, D. M., Avenevoli, S., Savill, M., Huber, R. S., Simon, T. J., Leckliter, I. N., Sher, K. J., & Loewy, R. L. (2018). Assessment of the Prodromal Questionnaire– Brief Child Version for Measurement of Self-reported Psychoticlike Experiences in Childhood. JAMA Psychiatry, 75(8), 853–861.

120. Hagler, D. J., Jr, Hatton, S., Cornejo, M. D., Makowski, C., Fair, D. A., Dick, A. S., Sutherland, M. T., Casey, B. J., Barch, D. M., Harms, M. P., Watts, R., Bjork, J. M., Garavan, H. P., Hilmer, L., Pung, C. J., Sicat, C. S., Kuperman, J., Bartsch, H., Xue, F., … Dale, A. M. (2019). Image processing and analysis methods for the Adolescent Brain Cognitive Development Study. Neuroimage, 202, 116091.

121. Gordon, E. M., Laumann, T. O., Adeyemo, B., Huckins, J. F., Kelley, W. M., & Petersen, S. E. (2016). Generation and Evaluation of a Cortical Area Parcellation from Resting-State Correlations. Cereb. Cortex, 26(1), 288–303.

122. Peterson, R. E., Kuchenbaecker, K., Walters, R. K., Chen, C.-Y., Popejoy, A. B., Periyasamy, S., Lam, M., Iyegbe, C., Strawbridge, R. J., Brick, L., Carey, C. E., Martin, A. R., Meyers, J. L., Su, J., Chen, J., Edwards, A. C., Kalungi, A., Koen, N., Majara, L., … Duncan, L. E. (2019). Genome-wide Association Studies in Ancestrally Diverse Populations: Opportunities, Methods, Pitfalls, and Recommendations. Cell, 179(3), 589–603.

123. Gogarten, S. M., Sofer, T., Chen, H., Yu, C., Brody, J. A., Thornton, T. A., Rice, K. M., & Conomos, M. P. (2019). Genetic association testing using the GENESIS R/Bioconductor package. Bioinformatics, 35(24), 5346–5348.

124. Conomos, M. P., Miller, M. B., & Thornton, T. A. (2015). Robust inference of population structure for ancestry prediction and correction of stratification in the presence of relatedness. Genet. Epidemiol., 39(4), 276–293.

125. Demontis, D., Walters, G. B., Athanasiadis, G., Walters, R., Therrien, K., Nielsen, T. T., Farajzadeh, L., Voloudakis, G., Bendl, J., Zeng, B., Zhang, W., Grove, J., Als, T. D., Duan, J., Satterstrom, F. K., Bybjerg-Grauholm, J., Bækved-Hansen, M., Gudmundsson, O. O., Magnusson, S. H., … Børglum, A. D. (2023). Genome-wide analyses of ADHD identify 27 risk loci, refine the genetic architecture and implicate several cognitive domains. Nat. Genet., 55(2), 198–208.

126. Grove, J., Ripke, S., Als, T. D., Mattheisen, M., Walters, R. K., Won, H., Pallesen, J., Agerbo, E., Andreassen, O. A., Anney, R., & Others. (2019). Identification of common genetic risk variants for autism spectrum disorder. Nat. Genet., 51(3), 431–444.

127. Wray, N. R., Ripke, S., Mattheisen, M., Trzaskowski, M., Byrne, E. M., Abdellaoui, A., Adams, M. J., Agerbo, E., Air, T. M., Andlauer, T. M. F., Bacanu, S.-A., Bækvad-Hansen, M., Beekman, A. F. T., Bigdeli, T. B., Binder, E. B., Blackwood, D. R. H., Bryois, J., Buttenschøn, H. N., Bybjerg-Grauholm, J., … Major Depressive Disorder Working Group of the Psychiatric Genomics Consortium. (2018). Genome-wide association analyses identify 44 risk variants and refine the genetic architecture of major depression. Nat. Genet., 50(5), 668–681.

128. Yu, D., Sul, J. H., Tsetsos, F., Nawaz, M. S., Huang, A. Y., Zelaya, I., Illmann, C., Osiecki, L., Darrow, S. M., Hirschtritt, M. E., Greenberg, E., Muller-Vahl, K. R., Stuhrmann, M., Dion, Y., Rouleau, G., Aschauer, H., Stamenkovic, M., Schlögelhofer, M., Sandor, P., … Tourette Association of America International Consortium for Genetics, the Gilles de la Tourette GWAS Replication Initiative, the Tourette International Collaborative Genetics Study, and the Psychiatric Genomics Consortium Tourette Syndrome Working Group. (2019). Interrogating the genetic determinants of Tourette’s syndrome and other tic disorders through genome-wide association studies. Am. J. Psychiatry, 176(3), 217–227.

129. Paul, S.E., Colbert, S.M.C., Gorelik, A.J., Hansen, I.S., Nagella, I., Blaydon, L., Hornstein, A., Johnson, E.C., Hatoum, A.S., Baranger, D.A.A., Elsayed, N.M., Barch, D.M., Bogdan, R., Karcher, N.R (2023). Phenome-wide Investigation of Behavioral, Environmental and Neural Associations with Cross-Disorder Genetic Liability in Youth of European Ancestry. medRxiv.

131. A. Lee, J. J., Wedow, R., Okbay, A., Kong, E., Maghzian, O., Zacher, M., Nguyen-Viet, T., Bowers, P., Sidorenko, J., Karlsson Linnér, R., Fontana, M. A., Kundu, T., Lee, C., Li, H., Li, R., Royer, R., Timshel, P. N., Walters, R. K., Willoughby, E. A., … Cesarini, D. (2018). Gene discovery and polygenic prediction from a genome-wide association study of educational attainment in 1.1 million individuals. Nat. Genet., 50(8), 1112–1121.

130. Wootton, O., Shadrin, A. A., Mohn, C., Susser, E., Ramesar, R., Gur, R. C., Andreassen, O. A., Stein, D. J., & Dalvie, S. (2023). Genome-wide association study in 404,302 individuals identifies 7 significant loci for reaction time variability. Mol. Psychiatry, 28(9), 4011–4019.

131. Ahern, J., Thompson, W., Fan, C. C., & Loughnan, R. (2023). Comparing pruning and thresholding with continuous shrinkage polygenic score methods in a large sample of ancestrally diverse adolescents from the ABCD study®. Behav. Genet., 53(3), 292–309.

134. Grotzinger, A. D., Rhemtulla, M., de Vlaming, R., Ritchie, S. J., Mallard, T. T., Hill, W. D., Ip, H. F., Marioni, R. E., McIntosh, A. M., Deary, I. J., Koellinger, P. D., Harden, K. P., Nivard, M. G., & Tucker-Drob, E. M. (2019). Genomic structural equation modelling provides insights into the multivariate genetic architecture of complex traits. Nat Hum Behav, 3(5), 513– 525.

133. Baker, E., Sims, R., Leonenko, G., Frizzati, A., Harwood, J. C., Grozeva, D., GERAD/PERADES, CHARGE, ADGC, EADI, IGAP consortia, Morgan, K., Passmore, P., Holmes, C., Powell, J., Brayne, C., Gill, M., Mead, S., Bossù, P., … Escott-Price, V. (2019). Gene-based analysis in HRC imputed genome wide association data identifies three novel genes for Alzheimer’s disease. PLoS One, 14(7), e0218111.

136. de Leeuw, C. A., Mooij, J. M., Heskes, T., & Posthuma, D. (2015). MAGMA: generalized gene-set analysis of GWAS data. PLoS Comput. Biol., 11(4), e1004219.

137. Brooks, M. E., Kristensen, K., Van Benthem, K. J., Magnusson, A., Berg, C. W., Nielsen, A., Skaug, H. J., Machler, M., & Bolker, B. M. (2017). glmmTMB balances speed and flexibility among packages for zero-inflated generalized linear mixed modeling. R J., 9(2), 378–400.

