## Supplementary Material for "Attention-mediated genetic influences on psychotic symptomatology in adolescence"

Table of Contents

**Supplementary Methods and Results**

1. Reliability and validity of the intra-individual variability (IIV) tasks
2. Details on the Prodromal Questionnaire - Brief Child Version (PQ-BC)
3. Recommendations for resting state scan inclusion (ABCD Study)
4. Recommendations for resting state scan inclusion (Chen et al. 2023)
5. Methods for partitioned polygenic scores
    a. PRS-CS
    b. Permutations
    c. PRS-CS vs PRSet
6. Information about WGCNA (Brainspan) modules
7. Additional schizophrenia PGS analysis
8. Sensitivity Analyses Considering Non-Random Missingness

**Supplementary Tables**

Supplementary Table 1: Baseline Demographics, Entire Sample
Supplementary Table 2: Phenotypic associations between individual task IIV and PQ-BC Distress (all ancestries included)
Supplementary Table 3: Phenotypic associations between Attention-related Functional Connectivity Network Metrics and PQ-BC Distress (all ancestries)
Supplementary Table 4: Linear Mixed Models: Associations between Polygenic Scores (PGS) and Intra-Individual Variability (IIV) Composite (participants of European ancestry only, n=5162)
Supplementary Table 5: Linear Mixed Models: Associations between Multi-Trait Polygenic Scores (PGS) and PQ-BC Distress (participants of European ancestry only, n=5162)
Supplementary Table 6: Null Associations between PGS and DMN-DAN Anticorrelation (per ABCD's inclusion recommendations)
Supplementary Table 7: Null Associations between PGS and DAN Functional Connectivity (per ABCD's inclusion recommendations)
Supplementary Table 8: ADHD PGS*time analysis (European descent)
Supplementary Table 9: Cognitive Performance PGS*time analysis (European descent)
Supplementary Table 10: Neurodevelopmental Disorders PGS*time analysis (European descent)
Supplementary Table 11: Schizophrenia PGS*time analysis (European descent)
Supplementary Table 12: One-sided permuted p-values across ancestries and pPGS.
Supplementary Table 13: Competitive p values across ancestries and pPGS.
Supplementary Table 14: Hurdle Models: Associations between IIV Composite and PQ-BC
Supplementary Table 15: Hurdle Models: Associations between Attention-related FC and PQ-BC
Supplementary Table 16: Hurdle Models: Associations between ADHD PGS and PQ-BC (European ancestry only)
Supplementary Table 17: Hurdle Models: Associations between CP PGS and PQ-BC (European ancestry only)
Supplementary Table 18: Hurdle Models: Associations between NDV PGS and PQ-BC (European ancestry only)
Supplementary Table 19: Hurdle Models: Associations between SCZ PGS and PQ-BC (European ancestry only)
Supplementary Table 20: Effect size estimates of ADHD pPGS on PQ-BC stratified by method of partitioning.
Supplementary Table 21: Model estimates of SCZ PGS derived from multi-ancestry GWAS
Supplementary Table 22: Comparing mean baseline scores to number of complete timepoints
Supplementary Table 23: Sensitivity analyses replicating PGS by time interactions on PLEs while accounting for missingness.

**Supplementary Figures**

Supplementary Figure 1: Schematic representation of data used across visits, by domain
Supplementary Figure 2: Relationships between module size and effect size.
Supplementary Figure 3: Heatmap of pPGS effects on PQ-BC in European, American/Latinx, and African samples.
Supplementary Figure 4: Overall polygenic scores calculation workflow
Supplementary Figure 5: Visualization of principal components for ancestral assignment
Supplementary Figure 6: Graphical comparison of methods to partition ADHD PGS.
Supplementary Figure 7: Graphical depiction of pPGS permutation significance criteria

Supplementary References

### Supplementary Methods and Results

**1. Reliability and validity of the intra-individual variability (IIV) tasks**

In a pediatric validation sample (N=208, ages=3-15), the Flanker and Dimensional Change Card Sort task have excellent test-retest reliability (ICC=0.92 for both measures; Zelazo et al. 2013) and the Processing Speed task has good test-retest reliability (ICC=0.84) in children and adolescents over a two-week interval (Carlozzi et al. 2013).

**2. Details on the Prodromal Questionnaire - Brief Child Version (PQ-BC)**

Subjects indicated whether they experienced each item (yes/no). If participants endorsed a given PLE, then they indicated whether the experience bothered them (yes/no), and if so, then subjects indicated the level of distress from the experience on a 5-point Likert scale. Consistent with previous research (Loewy et al. 2011, Cicero et al. 2019, Karcher et al. 2018), a summary score of PQ-BC Distress was calculated by the adding the total number of endorsed items weighted by distress (for each item, 0 = did not experience PLE, 1 = experienced PLE with no distress, 2-6 = distressing PLE [+1 to the distress score for the item]). PQ-BC Distress was used as the measure of psychotic-like experiences (PLEs) in all analyses.

**3. Quality control for resting-state scans recommended by the ABCD study**

For our primary analyses of network functional connectivity (FC) we used the resting-state data recommended for inclusion by the ABCD study for further analyses. These scans have passed the following quality control metrics, as per the ABCD study Wiki page (https://wiki.abcdstudy.org/release-notes/imaging/quality-control.html): Resting state MRI (rsMRI) passed rawQC, T1 passed raw QC, greater than 375 frames remaining after censoring, fMRI B0 unwarp available, passed FreeSurfer QC, passed fMRI manual post-processing QC, passed fMRI registration to T1, passed dorsal and ventral field of view cutoff score, and existing derived results. Further details about the neuroimaging processing can be found in Hagler et al. 2019. Baseline rsMRI scans included: N=9,619; Year 2 rsMRI scans included:N=6,851; *Year 4 rsMRI scans included: N=2,816, *Year 4 rsMRI data is only partially released as of ABCD Release 5.1; **Supplementary Figure 1**).

**4. Quality control for resting-state scans recommended by Chen et al. 2022**

We conducted secondary analyses using the more stringent motion correction parameters recommended by Chen et al. 2022. Specifically, to address potential motion and physiological artifacts in FC analyses, we excluded individuals with resting state scans with framewise displacement (FD) larger than 5mm and/or scans with had more than half of their volumes censored in Chen et al. 2022 (due to volumes with FD (framewise displacement) > 0.3 mm or derivative of root mean square variance over voxels > 50). The total N at the baseline study visit for this subset is 1854, and within this subsample, 860 were of European descent (used for EUR PGS analyses).

**5. Methods for partitioned polygenic scores**

a. PRS-CS

There is currently no clear consensus on the best method for accounting for linkage disequilibrium when generating pPGS. In typical clumping + thresholding methods (e.g., PRSice), a lead SNP is selected from each LD block with all other SNPs in a block being removed from analysis. In practice, this process may omit a portion of SNPs that map onto a particular gene set of interest. Thus, LD clumping is performed on a partitioned set of markers so that LD is accounted for while considering only SNPs in a partition/pathway of interest. With PRS-CS (used in this study), LD is not accounted for in the same way. Per LD block, effect sizes are shrunk via phi (general shrinkage parameter) and psi (marker specific shrinkage parameter, which is a function of a SNPs contribution to GWAS signal (Ge et al. 2019). Shrinkage is not linear, such that effects of important SNPs in an LD block tend to be preserved while small effects tend to be shrunk more. By partitioning summary statistics before running this shrinkage algorithm, the main (i.e., the most important) SNPs of LD blocks may be removed; alternatively, if the main SNP falls within a pathway of interest, it may be preserved while less important SNPs are removed. Such practice might inflate the effects of pathway-specific markers. Thus, we chose to provide PRS-CS with as much information as possible (i.e., whole-genome summary statistics) so that a SNP's contribution would not be biased by incomplete data for nearby SNPs. PRS-CS output (posterior effect sizes per marker) was then partitioned based on gene set. This choice additionally made permutation tests more computationally tractable.

To compare main results, PGS were calculated using the alternative approach to account for LD in sets of variants with PRS-CS. That is, first, summary statistics were partitioned by gene sets; then, shrinkage was performed on the partitioned summary statistics. Our approach was perhaps more conservative, as effect sizes tended to be smaller with our choice of pPGS method, though main findings and interpretations do not differ by method **(Supplementary Table 20, Supplementary Figure 6)**. Additionally, effect sizes did not differ significantly by method (mean difference = 0.003, $p > 0.05$).

b. pPGS Permutations

In addition to the reasons noted above, we chose to partition posterior SNP effects, as opposed to summary statistics, because permutation tests were made substantially more computationally tractable. Partitioning posterior effects ensured that the PRS-CS algorithm had to be implemented only once per module per ancestry; otherwise, PRS-CS would have to have been called 10,000 times per module per ancestry.

Of note, Choi et al. 2023 recommend testing significance of pPGS via competitive P value test whereby observed P-values are compared to permuted P values, which in effect is analogous to a two-sided test. Our pipeline diverged partially from this recommendation in that it placed emphasis too on one-sided P-values, which were derived by comparing observed to permuted effect sizes. Thus, our analyses tested whether the observed significance value was smaller than expected by chance *and* if the observed effect was greater (stronger) than the effects generated from a randomly sampled subset of SNPs of equal size. Comparing Supplementary Table 12 (one-sided P values) to Supplementary Table 13 (two-sided P values) reveals that the latter method results in a greater number of significant findings. Given the novelty of these methods, our interpretations thus rely on rather conservative criteria to make inference about implicated biology. For a module to be interpreted, its observed P-value must have passed FDR correction and both its competitive and one-sided P-value must have been less than 0.05. See **Supplementary Figure 7** for a depiction of these stringent criteria.

c. Comparison of PRS-CS to PRSet

To compare methods of generating partitioned PGS, additional pPGS were derived using PRSet with default parameters. Given that ADHD pPGS exhibited the most robust effects on PQ-BC, and thus were at greater risk of Type I error, comparisons of partitioning methods were made using these scores. ADHD GWAS summary statistics were partitioned by sets of SNPs identical to those used for PRS-CS-derived pPGS. Resultant scores were regressed onto PQ-BC scores and effect sizes were compared to those reported in the main text. There was a significant difference in effect sizes between PRS-CS and PRSet-derived modular ADHD pPGS, such that the average effect of PRS-CS-derived pPGS on PQ-BC was greater than those derived from PRSet (mean difference = 0.006, $P < 0.05$). Qualitatively, PRS-CS pPGS appeared to be better powered to detect differences at smaller module sizes (see **Supplementary Figure 6**); however, there was no relationship between module size and difference in beta coefficients ($ß = -2.52 \times 10^{-7}$, $P = 0.58$).

**6. Information about WGCNA (Brainspan) modules**

In Forsyth et al. 2020, modules M7, M13, and M15 were found to be enriched for both SCZ- and ASD-related common variants and copy number variants (CNVs). These modules were enriched for genes related broadly to synapse function and gene expression regulation. Genes in M7 show a peak in expression early in fetal development that declines towards birth; those in M13 steadily increase expression until birth at which point expression plateaus; and those in M15 increase expression through adolescence. ASD-related common variants and CNVs were additionally overrepresented in modules M1 and M4, both of which are enriched for genes involved in neuronal differentiation; M6, involved in translation and protein catabolism; and M12, involved in regulating membrane potential. De novo mutations found in ASD were also nominally enriched for M5, which was a neuronal module involved in membrane and synapse organization. In addition, genes in M5 exhibited a relatively volatile developmental expression pattern, with decreases in prenatal expression prior to birth and in postnatal expression around early childhood through early adolescence. Contrary to most other modules, a relatively large proportion of variance in M5 expression was explained by between-subject variance (Forsyth et al. 2020).

**7. Additional Schizophrenia PGS Analysis**

Given our finding that SCZ PGS was significantly associated with PQ-BC scores in EUR participants in spite of previously observed null associations in the same sample (ABCD, Hughes 2023, Karcher 2022, Hernandez 2023), we tested whether our selection of GWAS summary statistics influenced this discrepancy. The SCZ PGS were derived from the latest GWAS of SCZ (Trubetskoy et al. 2022), which made available several sets of summary statistics with each differing by the ancestral makeup of its discovery sample. Summary statistics used for main analyses and interpretations were homogeneously of European ancestry. We calculated another set of SCZ PGS scores using multi-ancestry GWAS summary statistics derived from EUR, AMR, AFR, and EAS samples and tested their effect on PQ-BC scores. This analysis too suggested a significant association; model estimates are reported in **Supplementary Table 21**.

**8. Sensitivity Analyses Considering Non-Random Missingness**

To test whether unmeasured characteristics associated with attrition drove the observed interaction effects of multi-trait PGS by time on PLEs, missingness was assessed for nonrandomness. These models specifically were chosen to test because, although all analyses used data from multiple timepoints, only these models directly tested an effect of time, which is more likely to inadvertently capture subject-level variance associated with attrition. Subjects were assigned to one of five groups (missingness groups) where the first group comprised of individuals with only 1 timepoint of PQ-BC data, the second with 2 timepoints of PQ-BC data, etc. Next, one-way ANOVA tested the null hypothesis that baseline PQ-BC and IIV scores did not differ by missingness group. The omnibus P-values (p's $\leq$ 0.001) suggested that the null be rejected, indicating that missingness was nonrandom with respect to baseline PQ-BC and IIV scores (**Supplementary Table 22**). Given this, main PGS by time interaction models with PQ-BC as the dependent variable were rerun with the inclusion of the missingness group variable. Results did not differ and are reported in **Supplementary Table 23**.

### Supplementary Tables

**Supplementary Table 1. Baseline Demographics, Entire Sample**

| | |
|---|---|
| Total Participants, N | 11,855 |
| Sex, N (%) | |
| Female | 5672(47.84) |
| Male | 6180(52.13) |
| Intersex | 3 (0.02) |
| Age, mean (SD) | 9.91(0.62) |
| Race/Ethnicity | |
| White | 6168 (52.02) |
| Black | 1782 (15.03) |
| Hispanic/Latinx | 2406 (20.3) |
| Asian | 251 (2.12) |
| Other/Mixed | 1246 (10.51) |
| Income Category, N (%) | |
| < 50k | 3218(27.08) |
| 50-99k | 3068(25.78) |
| 100k+ | 4561(38.33) |
| Refuse to report/don't know income | 1048(8.81) |

*N is based on the number of participants with complete PQ-BC information at the baseline study visit. Primary analyses of polygenic scores in the main text include participants of European ancestry only. See Figure 1 and Supplementary Figure 1 for overview of these analyses. Race/ethnicity in this table is based on self-report.*

**Supplementary Table 2. Linear Mixed Models: Phenotypic associations between individual task IIV and PQ-BC Distress Score** (all ancestries included)

| | PQ-BC Distress | | PQ-BC Distress | | PQ-BC Distress | | PQ-BC Distress | |
|---|---|---|---|---|---|---|---|---|
| *Predictors* | β | *std. CI* | β | *std. CI* | β | *std. CI* | β | *std. CI* |
| IIV Composite | 0.15 *** | 0.12 – 0.17 | | | | | | |
| Flanker IIV | | | 0.06 *** | 0.04 – 0.08 | | | | |
| DCCS IIV | | | | | 0.12 *** | 0.09 – 0.15 | | |
| Processing Speed IIV | | | | | | | 0.09 *** | 0.06 – 0.11 |
| Age | -0.15 *** | -0.17 – -0.14 | -0.16 *** | -0.18 – -0.15 | -0.11 *** | -0.16 – -0.06 | -0.16 *** | -0.17 – -0.14 |
| Sex | 0.01 | -0.03 – 0.04 | -0.00 | -0.04 – 0.03 | -0.05 * | -0.10 – -0.00 | 0.00 | -0.03 – 0.04 |
| 50-99,999k | -0.15 *** | -0.20 – -0.10 | -0.16 *** | -0.21 – -0.10 | -0.12 *** | -0.19 – -0.06 | -0.16 *** | -0.21 – -0.11 |
| 100k+ | -0.30 *** | -0.35 – -0.25 | -0.31 *** | -0.36 – -0.26 | -0.33 *** | -0.40 – -0.27 | -0.32 *** | -0.37 – -0.27 |
| Non-White | 0.13 *** | 0.08 – 0.17 | 0.14 *** | 0.09 – 0.18 | 0.13 *** | 0.08 – 0.19 | 0.13 *** | 0.09 – 0.18 |
| Marginal $R^2$ / Conditional $R^2$ | 0.063 / 0.391 | | 0.060 / 0.392 | | 0.040 / 0.949 | | 0.060 / 0.391 | |

*Abbreviations: PQ-BC = Prodromal Questionnaire - Brief Child Version, IIV = intra-individual variability, DCCS = Dimensional Change Card Sort*

*Reference categories: Sex (Male), Parental Income (<50k), Race/Ethnicity (White)*

* $p<0.05$ ** $p<0.01$ *** $p<0.001$

**Supplementary Table 3. Linear Mixed Models: Phenotypic associations between Attention-related Functional Connectivity Network Metrics and PQ-BC Distress** (including all ancestries, recommended by Chen et al. 2022)

| | PQ-BC Distress | | PQ-BC Distress | | PQ-BC Distress | | PQ-BC Distress | | PQ-BC Distress | |
|---|---|---|---|---|---|---|---|---|---|---|
| *Pred.* | β | *std. CI* | β | *std. CI* | β | *std. CI* | β | *std. CI* | β | *std. CI* |
| DMN-DAN anticorrelation | 1.01 *** | 0.45 – 1.58 | | | | | | | | |
| DMN-CON anticorrelation | | | 0.19 | -0.36 – 0.74 | | | | | | |
| DAN FC | | | | | -0.45 * | -0.89 – -0.01 | | | | |
| CON FC | | | | | | | -0.23 | -0.67 – 0.22 | | |
| DMN FC | | | | | | | | | -0.34 | -0.86 – 0.18 |
| Mean FD | 0.05 | -0.06 – 0.16 | 0.08 | -0.03 – 0.20 | 0.08 | -0.03 – 0.19 | 0.09 | -0.03 – 0.20 | 0.08 | -0.03 – 0.19 |
| Age | -0.13 *** | -0.16 – -0.11 | -0.13 *** | -0.16 – -0.11 | -0.13 *** | -0.16 – -0.11 | -0.13 *** | -0.16 – -0.11 | -0.13 *** | -0.16 – -0.11 |
| Sex | 0.06 | -0.01 – 0.12 | 0.05 | -0.01 – 0.12 | 0.05 | -0.02 – 0.11 | 0.05 | -0.01 – 0.12 | 0.06 | -0.01 – 0.12 |
| 50-99,999k | -0.14 ** | -0.24 – -0.04 | -0.15 ** | -0.24 – -0.05 | -0.14 ** | -0.24 – -0.04 | -0.14 ** | -0.24 – -0.05 | -0.14 ** | -0.24 – -0.05 |
| 100k+ | -0.27 *** | -0.36 – -0.17 | -0.28 *** | -0.37 – -0.18 | -0.27 *** | -0.37 – -0.18 | -0.28 *** | -0.37 – -0.18 | -0.28 *** | -0.37 – -0.18 |
| Non-White | 0.16 *** | 0.08 – 0.24 | 0.16 *** | 0.08 – 0.24 | 0.16 *** | 0.08 – 0.24 | 0.16 *** | 0.08 – 0.24 | 0.16 *** | 0.08 – 0.24 |
| Marginal $R^2$ / Conditional $R^2$ | 0.093 / NA | | 0.063 / 0.337 | | 0.089 / NA | | 0.064 / 0.337 | | 0.064 / 0.337 | |

*Abbreviations: PQ-BC (Prodromal Questionnaire - Brief Child Version), DMN = default mode network, DAN = dorsal attention network, CON = cingulo-opercular network, FD = framewise displacement*
*Reference groups: Sex (Male), Parental Income (<50k), Race/Ethnicity (White)*
** p<0.05 ** p<0.01 *** p<0.001*
*Recommended participants pass quality control criteria from Chen et al. 2022, details included in Supplementary Methods, section 4, N across visits in Supplementary Figure 1.*

**Supplementary Table 4.** Linear Mixed Models: Associations between Polygenic Scores (PGS) and Intra-Individual Variability (IIV) Composite (European ancestry only)

| | **IIV Composite** | | **IIV Composite** | | **IIV Composite** | | **IIV Composite** | | **IIV Composite** | |
|---|---|---|---|---|---|---|---|---|---|---|
| *Pred.* | β | *std. CI* | β | *std. CI* | β | *std. CI* | β | *std. CI* | β | *std. CI* |
| ADHD PGS | 0.04 *** | 0.03 – 0.05 | | | | | | | | |
| NDV PGS | | | 0.02 *** | 0.01 – 0.04 | | | | | | |
| SCZ PGS | | | | | 0.01 | -0.00 – 0.02 | | | | |
| CP PGS | | | | | | | -0.05 *** | -0.06 – -0.04 | | |
| IIV PGS | | | | | | | | | 0.02 *** | 0.01 – 0.03 |
| pc1 | -0.02 * | -0.04 – -0.00 | -0.02 * | -0.04 – -0.00 | -0.02 * | -0.03 – -0.00 | -0.02 | -0.03 – 0.00 | -0.02 | -0.03 – 0.00 |
| pc2 | -0.03 *** | -0.05 – -0.02 | -0.04 *** | -0.05 – -0.02 | -0.03 *** | -0.05 – -0.02 | -0.04 *** | -0.05 – -0.02 | -0.03 *** | -0.05 – -0.02 |
| pc3 | -0.02 ** | -0.04 – -0.01 | -0.02 ** | -0.04 – -0.01 | -0.02 ** | -0.04 – -0.01 | -0.02 * | -0.03 – -0.00 | -0.02 ** | -0.04 – -0.01 |
| pc4 | -0.01 | -0.02 – 0.01 | -0.01 | -0.02 – 0.01 | -0.01 | -0.02 – 0.01 | -0.01 | -0.02 – 0.01 | -0.01 | -0.02 – 0.01 |
| pc5 | 0.00 | -0.01 – 0.02 | 0.00 | -0.01 – 0.02 | 0.00 | -0.01 – 0.02 | -0.00 | -0.02 – 0.02 | 0.00 | -0.01 – 0.02 |
| Age | -0.15 *** | -0.16 – -0.14 | -0.15 *** | -0.16 – -0.14 | -0.15 *** | -0.16 – -0.14 | -0.15 *** | -0.16 – -0.14 | -0.15 *** | -0.16 – -0.14 |
| Sex | -0.07 *** | -0.09 – -0.05 | -0.07 *** | -0.09 – -0.05 | -0.07 *** | -0.09 – -0.05 | -0.07 *** | -0.09 – -0.05 | -0.07 *** | -0.09 – -0.05 |
| Marginal $R^2$ / Conditional $R^2$ | 0.106 / 0.375 | | 0.102 / 0.375 | | 0.100 / 0.376 | | 0.109 / 0.374 | | 0.102 / 0.376 | |

*Abbreviations: CP = cognitive performance; SCZ = schizophrenia; NDV = neurodevelopmental disorders; IIV = intra-individual variability; PGS = polygenic score; pc = ancestry principal component; FD = framewise displacement*

*Reference category: Sex (male)*

* *p<0.05* ** *p<0.01* *** *p<0.001*

**Supplementary Table 5.** Linear Mixed Effects Models: Associations between Multi-trait PGS and PQ-BC Distress Score (European ancestry only)

| | PQ-BC Distress | | PQ-BC Distress | | PQ-BC Distress | | PQ-BC Distress | | PQ-BC Distress | |
|---|---|---|---|---|---|---|---|---|---|---|
| *Pred.* | β | *std. CI* | β | *std. CI* | β | *std. CI* | β | *std. CI* | β | *std. CI* |
| ADHD PGS | 0.08 *** | 0.07 – 0.10 | | | | | | | | |
| NDV PGS | | | 0.08 *** | 0.06 – 0.10 | | | | | | |
| SCZ PGS | | | | | 0.02 ** | 0.01 – 0.04 | | | | |
| CP PGS | | | | | | | -0.04 *** | -0.06 – -0.03 | | |
| IIV PGS | | | | | | | | | 0.01 | -0.01 – 0.02 |
| pc1 | -0.02 | -0.05 – 0.00 | -0.03 * | -0.05 – -0.00 | -0.02 | -0.04 – 0.00 | -0.02 | -0.04 – 0.01 | -0.02 | -0.04 – 0.01 |
| pc2 | -0.03 * | -0.05 – -0.00 | -0.03 * | -0.05 – -0.01 | -0.03 * | -0.05 – -0.00 | -0.03 * | -0.05 – -0.00 | -0.03 * | -0.05 – -0.00 |
| pc3 | -0.01 | -0.03 – 0.01 | -0.01 | -0.03 – 0.01 | -0.01 | -0.03 – 0.01 | -0.01 | -0.03 – 0.01 | -0.01 | -0.04 – 0.01 |
| pc4 | 0.02 | -0.01 – 0.04 | 0.02 | -0.01 – 0.04 | 0.02 | -0.01 – 0.04 | 0.02 | -0.01 – 0.04 | 0.02 | -0.01 – 0.04 |
| pc5 | 0.00 | -0.02 – 0.03 | 0.00 | -0.02 – 0.03 | 0.00 | -0.02 – 0.03 | 0.00 | -0.02 – 0.02 | 0.00 | -0.02 – 0.03 |
| Age | -0.13 *** | -0.14 – -0.12 | -0.13 *** | -0.14 – -0.12 | -0.13 *** | -0.14 – -0.12 | -0.13 *** | -0.14 – -0.12 | -0.13 *** | -0.14 – -0.12 |
| Sex | 0.05 ** | 0.01 – 0.08 | 0.05 ** | 0.02 – 0.08 | 0.05 ** | 0.02 – 0.08 | 0.05 ** | 0.02 – 0.08 | 0.05 ** | 0.02 – 0.08 |
| Marginal $R^2$ / Conditional $R^2$ | 0.035 / 0.397 | | 0.035 / 0.396 | | 0.027 / 0.397 | | 0.028 / 0.397 | | 0.026 / 0.397 | |

*Abbreviations: PQ-BC = Prodromal Questionnaire- Brief Child Version, NDV = neurodevelopmental disorders, SCZ = schizophrenia; CP = cognitive performance, IIV = intra-individual variability; PGS = polygenic score; pc = ancestry principal component; FD = framewise displacement*

*Reference category: Sex (male)*

* *p<0.05* ** *p<0.01* *** *p<0.001*

**Supplementary Table 6.** Null Associations between PGS and DMN-DAN Anticorrelation (per ABCD's inclusion recommendations), European ancestry participants

| | **DMN-DAN anticorrelation** | | **DMN-DAN anticorrelation** | | **DMN-DAN anticorrelation** | | **DMN-DAN anticorrelation** | | **DMN-DAN anticorrelation** | |
|---|---|---|---|---|---|---|---|---|---|---|
| *Pred.* | β | *std. CI* | β | *std. CI* | β | *std. CI* | β | *std. CI* | β | *std. CI* |
| ADHD PGS | <0.001 | -0.001 – 0.001 | | | | | | | | |
| NDV PGS | | | 0.001 | <-0.001 – 0.002 | | | | | | |
| SCZ PGS | | | | | -0.001 | -0.002 – 0.000 | | | | |
| CP PGS | | | | | | | -0.001 | -0.002 – 0.000 | | |
| IIV PGS | | | | | | | | | <-0.001 | -0.001 – 0.001 |
| pc1 | <-0.001 | -0.002 – 0.002 | <-0.001 | -0.002 – 0.002 | <-0.001 | -0.002 – 0.002 | -0.000 | -0.002 – 0.002 | <-0.001 | -0.002 – 0.002 |
| pc2 | -0.001 | -0.002 – 0.001 | -0.001 | -0.003 – 0.001 | -0.001 | -0.002 – 0.001 | -0.001 | -0.003 – 0.001 | -0.001 | -0.002 – 0.001 |
| pc3 | -0.002 | -0.003 – 0.000 | -0.002 | -0.003 – 0.000 | -0.002 | -0.003 – 0.000 | -0.002 | -0.003 – 0.000 | -0.002 | -0.003 – 0.000 |
| pc4 | -0.000 | -0.002 – 0.001 | -0.000 | -0.002 – 0.001 | -0.000 | -0.002 – 0.001 | -0.000 | -0.002 – 0.001 | <-0.001 | -0.002 – 0.001 |
| pc5 | 0.001 | -0.001 – 0.002 | 0.001 | -0.001 – 0.002 | 0.001 | -0.001 – 0.002 | 0.001 | -0.001 – 0.002 | 0.001 | -0.001 – 0.002 |
| Mean FD | 0.019 *** | 0.017 – 0.021 | 0.019 *** | 0.017 – 0.021 | 0.019 *** | 0.017 – 0.021 | 0.019 *** | 0.017 – 0.021 | 0.019 *** | 0.017 – 0.021 |
| Age | -0.003 *** | -0.004 – -0.002 | -0.003 *** | -0.004 – -0.002 | -0.003 *** | -0.004 – -0.002 | -0.003 *** | -0.004 – -0.002 | -0.003 *** | -0.004 – -0.002 |
| Sex | -0.011 *** | -0.014 – -0.009 | -0.011 *** | -0.014 – -0.009 | -0.011 *** | -0.014 – -0.009 | -0.011 *** | -0.014 – -0.009 | -0.011 *** | -0.014 – -0.009 |
| Marginal $R^2$ / Conditional $R^2$ | 0.060 / 0.500 | | 0.060 / 0.500 | | 0.060 / 0.500 | | 0.060 / 0.500 | | 0.060 / 0.500 | |

*Abbreviations: DMN = default mode network, DAN = dorsal attention network, NDV = neurodevelopmental disorders, SCZ = schizophrenia; CP = cognitive performance, IIV = intra-individual variability; PGS = polygenic score; pc = ancestry principal component; FD = framewise displacement; Reference groups: Sex (Male); N's by time point for participants of European descent with rsMRI scans that met ABCD's inclusion recommendation (Baseline N=4,966; 2-year N=3,439; 4-year=1,458)*

*Significance = * p<0.05 ** p<0.01 *** p<0.001*

**Supplementary Table 7.** Null Associations between PGS and DAN Functional Connectivity (per ABCD's inclusion recommendations), European ancestry participants

| | DAN FC | | DAN FC | | DAN FC | | DAN FC | | DAN FC | |
|---|---|---|---|---|---|---|---|---|---|---|
| *Pred.* | β | *std. CI* | β | *std. CI* | β | *std. CI* | β | *std. CI* | β | *std. CI* |
| ADHD PGS | -0.002 | -0.003 – 0.000 | | | | | | | | |
| NDV PGS | | | -0.002 * | -0.004 – -0.000 | | | | | | |
| SCZ PGS | | | | | 0.001 | -0.001 – 0.002 | | | | |
| CP PGS | | | | | | | -0.000 | -0.002 – 0.002 | | |
| IIV PGS | | | | | | | | | -0.000 | -0.002 – 0.002 |
| pc1 | 0.002 | -0.000 – 0.005 | 0.002 | -0.000 – 0.005 | 0.002 | -0.001 – 0.004 | 0.002 | -0.001 – 0.004 | 0.002 | -0.001 – 0.004 |
| pc2 | 0.001 | -0.002 – 0.003 | 0.001 | -0.002 – 0.003 | 0.001 | -0.002 – 0.003 | 0.001 | -0.002 – 0.003 | 0.001 | -0.002 – 0.003 |
| pc3 | 0.001 | -0.001 – 0.003 | 0.001 | -0.001 – 0.003 | 0.001 | -0.001 – 0.003 | 0.001 | -0.001 – 0.003 | 0.001 | -0.001 – 0.003 |
| pc4 | -0.001 | -0.003 – 0.002 | -0.001 | -0.003 – 0.002 | -0.001 | -0.003 – 0.002 | -0.001 | -0.003 – 0.002 | -0.001 | -0.003 – 0.002 |
| pc5 | 0.002 | -0.001 – 0.004 | 0.002 | -0.001 – 0.004 | 0.002 | -0.001 – 0.004 | 0.002 | -0.001 – 0.004 | 0.002 | -0.001 – 0.004 |
| Mean FD | -0.015 *** | -0.017 – -0.012 | -0.015 *** | -0.017 – -0.012 | -0.015 *** | -0.017 – -0.012 | -0.015 *** | -0.017 – -0.012 | -0.015 *** | -0.017 – -0.012 |
| Age | 0.004 *** | 0.003 – 0.005 | 0.004 *** | 0.003 – 0.005 | 0.004 *** | 0.003 – 0.005 | 0.004 *** | 0.003 – 0.005 | 0.004 *** | 0.003 – 0.005 |
| Sex | 0.004 * | 0.000 – 0.007 | 0.004 * | 0.000 – 0.007 | 0.004 * | 0.000 – 0.007 | 0.004 * | 0.000 – 0.007 | 0.004 * | 0.000 – 0.007 |
| Marginal $R^2$ / Conditional $R^2$ | 0.024 / 0.584 | | 0.024 / 0.584 | | 0.023 / 0.584 | | 0.023 / 0.584 | | 0.023 / 0.584 | |

*Abbreviations: DAN = dorsal attention network, FC = functional connectivity, NDV = neurodevelopmental disorders, SCZ = schizophrenia; CP = cognitive performance, IIV = intra-individual variability; PGS = polygenic score; pc = ancestry principal component; FD = framewise displacement Reference groups: Sex (Male)*

*N's by time point for participants of European descent with rsMRI scans that met ABCD's inclusion recommendation (Baseline N=4,966; 2-year N=3,439; 4-year=1,458)*

*Significance = * p<0.05 ** p<0.01 *** p<0.001*

**Supplementary Table 8.**

**ADHD PGS by Timepoint Interaction**

| | **PQ-BC Distress** | |
|---|---|---|
| *Predictors* | *Estimates* | *CI* |
| (Intercept) | -0.04 | -0.12 – 0.03 |
| ADHD PGS | 0.11 *** | 0.09 – 0.13 |
| Year 1 | -0.14 *** | -0.18 – -0.11 |
| Year 2 | -0.20 *** | -0.26 – -0.15 |
| Year 3 | -0.20 *** | -0.28 – -0.13 |
| Year 4 | -0.19 *** | -0.29 – -0.08 |
| PC1 | -1.14 | -2.32 – 0.04 |
| PC2 | -1.22 * | -2.41 – -0.04 |
| PC3 | -0.40 | -1.56 – 0.76 |
| PC4 | 0.86 | -0.28 – 2.01 |
| PC5 | 0.20 | -0.95 – 1.34 |
| Age | -0.06 *** | -0.09 – -0.02 |
| Sex | 0.05 ** | 0.02 – 0.08 |
| ADHD PGS x Year 1 | -0.03 * | -0.05 – -0.00 |
| ADHD PGS x Year 2 | -0.04 ** | -0.06 – -0.01 |
| ADHD PGS x Year 3 | -0.04 ** | -0.07 – -0.02 |
| ADHD PGS x Year 4 | -0.06 *** | -0.10 – -0.03 |
| Marginal $R^2$ / Conditional $R^2$ | 0.036 / 0.401 | |

* *p<0.05* ** *p<0.01* *** *p<0.001*

**Supplementary Table 8.** Effect sizes and confidence intervals from models examining the interaction effects of time and ADHD PGS on PQ-BC scores. Models included age, sex, and the top 5 genetic PCs as fixed effects with subject nested within family within site. Timepoint and its interaction with ADHD PGS was included as a fixed effect with baseline (Year 0) as the referent group. Reported confidence intervals reflect 95% confidence intervals.

**Supplementary Table 9.**

**CP PGS by Timepoint Interaction**

| | **PQ-BC Distress** | |
|---|---|---|
| *Predictors* | *Estimates* | *CI* |
| (Intercept) | -0.04 | -0.12 – 0.03 |
| CP PGS | -0.07 *** | -0.10 – -0.05 |
| Year 1 | -0.14 *** | -0.18 – -0.11 |
| Year 2 | -0.20 *** | -0.26 – -0.15 |
| Year 3 | -0.21 *** | -0.28 – -0.13 |
| Year 4 | -0.19 *** | -0.29 – -0.09 |
| PC1 | -0.81 | -2.00 – 0.37 |
| PC2 | -1.32 * | -2.51 – -0.13 |
| PC3 | -0.39 | -1.57 – 0.78 |
| PC4 | 0.83 | -0.33 – 1.98 |
| PC5 | 0.17 | -0.99 – 1.32 |
| Age | -0.06 ** | -0.09 – -0.02 |
| Sex | 0.05 ** | 0.02 – 0.08 |
| CP PGS x Year 1 | 0.03 * | 0.01 – 0.06 |
| CP PGS x Year 2 | 0.04 *** | 0.02 – 0.07 |
| CP PGS x Year 3 | 0.04 ** | 0.02 – 0.07 |
| CP PGS x Year 4 | 0.05 ** | 0.02 – 0.09 |
| Marginal $R^2$ / Conditional $R^2$ | 0.029 / 0.402 | |

* *p<0.05* ** *p<0.01* *** *p<0.001*

**Supplementary Table 9.** Effect sizes and confidence intervals from models examining the interaction effects of time and CP PGS on PQ-BC scores. Models included age, sex, and the top 5 genetic PCs as fixed effects with subject nested within family within site. Timepoint and its interaction with CP PGS was included as a fixed effect with baseline (Year 0) as the referent group. Reported confidence intervals reflect 95% confidence intervals.

**Supplementary Table 10.**

| **NDV PGS by Timepoint Interaction** | | |
|---|---|---|
| | **PQ-BC Distress** | |
| *Predictors* | *Estimates* | *CI* |
| (Intercept) | -0.05 | -0.13 – 0.03 |
| NDV PGS | 0.11 *** | 0.09 – 0.14 |
| Year 1 | -0.14 *** | -0.17 – -0.11 |
| Year 2 | -0.20 *** | -0.25 – -0.14 |
| Year 3 | -0.20 *** | -0.27 – -0.12 |
| Year 4 | -0.18 *** | -0.28 – -0.08 |
| PC1 | -1.45 * | -2.63 – -0.27 |
| PC2 | -1.46 * | -2.64 – -0.27 |
| PC3 | -0.38 | -1.55 – 0.78 |
| PC4 | 0.87 | -0.27 – 2.02 |
| PC5 | 0.20 | -0.95 – 1.35 |
| Age | -0.06 *** | -0.10 – -0.03 |
| Sex | 0.05 ** | 0.02 – 0.08 |
| NDV PGS x Year 1 | -0.03 ** | -0.06 – -0.01 |
| NDV PGS x Year 2 | -0.04 *** | -0.07 – -0.02 |
| NDV PGS x Year 3 | -0.05 *** | -0.08 – -0.02 |
| NDV PGS x Year 4 | -0.08 *** | -0.11 – -0.04 |
| Marginal $R^2$ / Conditional $R^2$ | 0.035 / 0.401 | |

* $p<0.05$ ** $p<0.01$ *** $p<0.001$

**Supplementary Table 10.** Effect sizes and confidence intervals from models examining the interaction effects of time and NDV PGS on PQ-BC scores. Models included age, sex, and the top 5 genetic PCs as fixed effects with subject nested within family within site. Timepoint and its interaction with NDV PGS was included as a fixed effect with baseline (Year 0) as the referent group. Reported confidence intervals reflect 95% confidence intervals.

**Supplementary Table 11.**

**SCZ PGS by Timepoint Interaction**

| | **PQ-BC Distress** | |
|---|---|---|
| *Predictors* | *Estimates* | *CI* |
| (Intercept) | -0.04 | -0.12 – 0.04 |
| SCZ PGS | 0.03 * | 0.00 – 0.05 |
| Year 1 | -0.14 *** | -0.18 – -0.11 |
| Year 2 | -0.21 *** | -0.26 – -0.15 |
| Year 3 | -0.21 *** | -0.29 – -0.14 |
| Year 4 | -0.20 *** | -0.30 – -0.09 |
| PC1 | -1.02 | -2.22 – 0.17 |
| PC2 | -1.31 * | -2.50 – -0.12 |
| PC3 | -0.53 | -1.70 – 0.64 |
| PC4 | 0.88 | -0.28 – 2.04 |
| PC5 | 0.21 | -0.95 – 1.37 |
| Age | -0.06 ** | -0.09 – -0.02 |
| Sex | 0.05 ** | 0.02 – 0.08 |
| SCZ PGS x Year 1 | 0.00 | -0.02 – 0.03 |
| SCZ PGS x Year 2 | -0.01 | -0.03 – 0.02 |
| SCZ PGS x Year 3 | 0.00 | -0.02 – 0.03 |
| SCZ PGS x Year 4 | -0.00 | -0.04 – 0.03 |
| Marginal $R^2$ / Conditional $R^2$ | 0.027 / 0.401 | |

* *p<0.05* ** *p<0.01* *** *p<0.001*

**Supplementary Table 11.** Effect sizes and confidence intervals from models examining the interaction effects of time and SCZ PGS on PQ-BC scores. Models included age, sex, and the top 5 genetic PCs as fixed effects with subject nested within family within site. Timepoint and its interaction with SCZ PGS was included as a fixed effect with baseline (Year 0) as the referent group. Reported confidence intervals reflect 95% confidence intervals.

**Supplementary Table 12.**

| One-sided permuted P-values | | | | | | | | | | | | |
|---|---|---|---|---|---|---|---|---|---|---|---|---|
| | **EUR** | | | | **AMR** | | | | **AFR** | | | |
| | ADHD | NDV | SCZ | CP | ADHD | NDV | SCZ | CP | ADHD | NDV | SCZ | CP |
| **M1** | 0.985* | 0.845* | - | 0.097* | - | - | - | - | - | - | - | - |
| **M2** | - | - | - | - | - | - | 0.007 | - | 0.042 | - | - | - |
| **M3** | - | - | - | - | - | - | - | - | - | - | - | - |
| **M4** | - | - | - | - | - | - | - | - | - | - | - | - |
| **M5** | 0.033* | 0.278* | - | - | - | - | - | - | - | - | - | - |
| **M6** | - | - | - | - | - | - | - | - | 0.048 | 0.032 | - | - |
| **M7** | 1* | 0.895* | - | - | - | - | - | - | 0.002 | - | - | - |
| **M8** | - | - | - | - | - | - | - | - | - | - | - | 0.013 |
| **M9** | - | - | 0.285 | - | - | - | - | - | - | - | - | - |
| **M10** | - | - | 0.026 | - | - | - | - | - | - | - | - | 0 |
| **M11** | 1* | 0.923* | 0.002 | 0.933* | - | - | 0.002 | - | - | - | - | - |
| **M12** | 0.739 | 0.424* | - | - | - | - | - | - | - | - | - | - |
| **M13** | - | - | - | - | - | - | - | - | - | - | - | - |
| **M14** | 0.203* | 0.463 | - | - | 0.032 | - | - | - | - | - | - | - |
| **M15** | 0.965* | 0.74* | 0.006 | 0.828* | - | - | - | - | - | - | - | - |
| **M16** | 0.237 | 0.358 | - | 0.16 | 0.003 | - | - | - | - | - | - | - |
| **M17** | 0.998* | 0.792* | - | - | - | - | - | - | - | - | - | - |
| **M18** | 0.708* | 0.772* | - | 0.502* | - | - | - | - | - | - | - | - |

* = corresponding observed associations significant after FDR correction

**Supplementary Table 12.** One-sided permuted p-values across ancestries and pPGS. Values shown represent one-sided null p-values from pPGS-PLE associations that are significant before correction for multiple comparisons ($p < 0.05$). One-sided null p-values represent the proportion of the absolute values of permuted null effect sizes that were greater than the absolute value of the observed effect. Asterisks indicate observed pPGS-PLE associations that are significant after FDR correction.

**Supplementary Table 13.**

| Competitive P-values | | | | | | | | | | | | |
|---|---|---|---|---|---|---|---|---|---|---|---|---|
| | **EUR** | | | | **AMR** | | | | **AFR** | | | |
| | ADHD | NDV | SCZ | CP | ADHD | NDV | SCZ | CP | ADHD | NDV | SCZ | CP |
| **M1** | 0.985* | 0.059* | - | 0.094* | - | - | - | - | - | - | - | - |
| **M2** | - | - | - | - | - | - | 0.023 | - | 0.056 | - | - | - |
| **M3** | - | - | - | - | - | - | - | - | - | - | - | - |
| **M4** | - | - | - | - | - | - | - | - | - | - | - | - |
| **M5** | 0.033* | 0.009* | - | - | - | - | - | - | - | - | - | - |
| **M6** | - | - | - | - | - | - | - | - | 0.061 | 0.042 | - | - |
| **M7** | 1* | 0.092* | - | - | - | - | - | - | 0.005 | - | - | - |
| **M8** | - | - | - | - | - | - | - | - | - | - | - | 0.027 |
| **M9** | - | - | 0.029 | - | - | - | - | - | - | - | - | - |
| **M10** | - | - | 0.041 | - | - | - | - | - | - | - | - | 0.002 |
| **M11** | 1 | 0.075* | 0.085 | 0.936* | - | - | 0.002 | - | - | - | - | - |
| **M12** | 0.742 | 0.027* | - | - | - | - | - | - | - | - | - | - |
| **M13** | - | - | - | - | - | - | - | - | - | - | - | - |
| **M14** | 0.205* | 0.101 | - | - | 0.033 | - | - | - | - | - | - | - |
| **M15** | 0.964* | 0.004* | 0.085 | 0.83* | - | - | - | - | - | - | - | - |
| **M16** | 0.237 | 0.044 | - | 0.159 | 0.023 | - | - | - | - | - | - | - |
| **M17** | 0.998* | 0.134* | - | - | - | - | - | - | - | - | - | - |
| **M18** | 0.704* | 0.135 | - | 0.505* | - | - | - | - | - | - | - | - |

* = corresponding observed associations significant after FDR correction

**Supplementary Table 13.** Competitive p-values across ancestries and pPGS. Values shown represent competitive p-values, which are two-sided and represent the proportion of permuted null p-values that were less than (i.e., more significant) the observed p-value. Asterisks indicate observed associations that were significant after FDR correction. Dashes indicate instances in which permutation was not performed because the observed associations between raw (i.e., original, non-permuted) pPGS and PQ-BC were not significant before correction for multiple comparisons ($p < 0.05$).

**Supplementary Table 14.**

Hurdle Models: Associations between IIV Composite and PQ-BC

| | PQ-BC | |
|---|---|---|
| *Predictors* | *std. Beta* | *standardized CI* |
| **Count Model** | | |
| (Intercept) | 7.17 *** | 6.64 – 7.76 |
| IIV Composite z-score | 1.08 *** | 1.05 – 1.10 |
| Age | 0.86 *** | 0.84 – 0.88 |
| Sex | 1.03 * | 1.01 – 1.06 |
| 50-99,999k | 0.89 ** | 0.84 – 0.96 |
| 100k+ | 0.77 *** | 0.72 – 0.83 |
| Non-White | 1.13 *** | 1.06 – 1.20 |
| **Zero-Inflated Model** | | |
| (Intercept) | 0.72 ** | 0.58 – 0.88 |
| IIV Composite z-score | 0.85 *** | 0.81 – 0.88 |
| Age | 1.63 *** | 1.56 – 1.70 |
| Sex | 1.03 | 0.99 – 1.07 |
| 50-99,999k | 1.37 *** | 1.22 – 1.54 |
| 100k+ | 1.95 *** | 1.73 – 2.19 |
| Non-White | 0.73 *** | 0.66 – 0.81 |
| Marginal $R^2$ | 0.057 | |

$^{*} p<0.05 \quad ^{**} p<0.01 \quad ^{***} p<0.001$

IIV = intra-individual variability in reaction time. Reference groups = income (<50,000k); race/ethnicity (White), sex (Male).

**Supplementary Table 15.**

Hurdle Models: Associations between attention-related functional connectivity and PQ-BC

| | **PQ-BC** | | **PQ-BC** | |
|---|---|---|---|---|
| *Predictors* | *std. Beta* | *std. CI* | *std. Beta* | *std. CI* |
| (Intercept) | 5.77 *** | 5.31 – 6.25 | 5.81 *** | 5.35 – 6.30 |
| DMN-DAN anticorrelation | 1.09 ** | 1.03 – 1.16 | | |
| DAN Functional Connectivity | | | 0.96 | 0.91 – 1.02 |
| Mean FD | 1.03 | 0.98 – 1.08 | 1.04 | 0.99 – 1.09 |
| Age | 0.83 *** | 0.79 – 0.88 | 0.83 *** | 0.79 – 0.88 |
| Sex | 1.09 ** | 1.02 – 1.15 | 1.08 ** | 1.02 – 1.14 |
| **Zero-Inflated Model** | | | | |
| (Intercept) | 1.30 * | 1.06 – 1.59 | 1.29 * | 1.05 – 1.58 |
| DMN-DAN anticorrelation | 0.80 *** | 0.73 – 0.87 | | |
| DAN anticorrelation | | | 1.15 ** | 1.05 – 1.26 |
| Mean FD | 0.95 | 0.87 – 1.03 | 0.92 | 0.85 – 1.00 |
| Age | 1.62 *** | 1.48 – 1.77 | 1.62 *** | 1.48 – 1.76 |
| Sex | 0.98 | 0.90 – 1.07 | 0.99 | 0.91 – 1.09 |

| | | |
|---|---|---|
| Marginal $R^2$ / Conditional $R^2$ | 0.047 / 0.454 | 0.070 / NA |

** p<0.05 ** p<0.01 *** p<0.001*

Abbreviations: DMN = default mode network; DAN = dorsal attention network; FD = framewise displacement

**Supplementary Table 16.**
Hurdle Models: Associations between ADHD PGS and PQ-BC (European participants only)

| | PQ-BC | |
|---|---|---|
| *Predictors* | *std. Beta* | *standardized CI* |
| **Count Model** | | |
| (Intercept) | 5.24 *** | 5.05 – 5.44 |
| ADHD EUR PGS | 1.11 *** | 1.08 – 1.14 |
| pc1 | 0.97 * | 0.94 – 1.00 |
| pc2 | 0.96 ** | 0.93 – 0.99 |
| pc3 | 0.98 | 0.96 – 1.01 |
| pc4 | 1.01 | 0.98 – 1.04 |
| pc5 | 1.00 | 0.97 – 1.02 |
| Age | 0.84 *** | 0.82 – 0.86 |
| Sex | 1.08 *** | 1.05 – 1.11 |
| **Zero-Inflated Model** | | |
| (Intercept) | 1.66 *** | 1.43 – 1.93 |
| ADHD EUR PGS | 0.76 *** | 0.72 – 0.80 |
| pc1 | 1.02 | 0.97 – 1.08 |
| pc2 | 1.02 | 0.96 – 1.07 |
| pc3 | 1.01 | 0.96 – 1.07 |
| pc4 | 0.99 | 0.94 – 1.05 |

| | | |
|---|---|---|
| pc5 | 0.98 | 0.93 – 1.03 |
| Age | 1.78 *** | 1.71 – 1.84 |
| Sex | 0.99 | 0.94 – 1.04 |
| Marginal $R^2$ | 0.047 | |

* *p<0.05* ** *p<0.01* *** *p<0.001*

Abbreviations: ADHD = attention deficit/hyperactivity disorder; EUR = European; PGS = polygenic score; pc = ancestry principal component; reference group (sex): Male.

**Supplementary Table 17.**
Hurdle Models: Associations between CP PGS and PQ-BC (European participants only)

| | PQ-BC | |
|---|---|---|
| *Predictors* | *std. Beta* | *standardized CI* |
| **Count Model** | | |
| (Intercept) | 5.28 *** | 5.08 – 5.49 |
| CP EUR PGS | 0.93 *** | 0.91 – 0.96 |
| pc1 | 0.97 | 0.95 – 1.00 |
| pc2 | 0.96 ** | 0.93 – 0.99 |
| pc3 | 0.99 | 0.96 – 1.02 |
| pc4 | 1.01 | 0.98 – 1.04 |
| pc5 | 0.99 | 0.97 – 1.02 |
| Age | 0.84 *** | 0.82 – 0.86 |
| Sex | 1.09 *** | 1.06 – 1.12 |
| **Zero-Inflated Model** | | |
| (Intercept) | 1.65 *** | 1.41 – 1.92 |
| CP EUR PGS | 1.12 *** | 1.06 – 1.18 |
| pc1 | 1.01 | 0.95 – 1.06 |
| pc2 | 1.02 | 0.96 – 1.08 |

| | | |
|---|---|---|
| pc3 | 1.01 | 0.96 – 1.07 |
| pc4 | 0.99 | 0.94 – 1.05 |
| pc5 | 0.98 | 0.93 – 1.03 |
| Age | 1.78 *** | 1.71 – 1.84 |
| Sex | 0.98 | 0.93 – 1.04 |
| Marginal $R^2$ / Conditional $R^2$ | 0.041 / 0.438 | |

*Abbreviations: CP = cognitive performance; EUR = European ancestry; pc = ancestry principal components. Reference group (sex): Male. * p<0.05 ** p<0.01 *** p<0.001*

**Supplementary Table 18.**
Hurdle Models: Associations between NDV PGS and PQ-BC (European participants only)

| | PQ-BC | |
|---|---|---|
| *Predictors* | *std. Beta* | *standardized CI* |
| **Count Model** | | |
| (Intercept) | 5.24 *** | 5.05 – 5.43 |
| NDV EUR PGS | 1.11 *** | 1.08 – 1.15 |
| pc1 | 0.96 ** | 0.93 – 0.99 |
| pc2 | 0.96 ** | 0.93 – 0.99 |
| pc3 | 0.99 | 0.96 – 1.01 |
| pc4 | 1.01 | 0.98 – 1.04 |
| pc5 | 1.00 | 0.97 – 1.03 |
| Age | 0.84 *** | 0.82 – 0.86 |
| Sex | 1.09 *** | 1.06 – 1.12 |
| **Zero-Inflated Model** | | |
| (Intercept) | 1.66 *** | 1.43 – 1.93 |
| NDV EUR PGS | 0.76 *** | 0.72 – 0.80 |
| pc1 | 1.04 | 0.98 – 1.10 |
| pc2 | 1.03 | 0.97 – 1.09 |
| pc3 | 1.01 | 0.96 – 1.07 |

| | | |
|---|---|---|
| pc4 | 0.99 | 0.94 – 1.05 |
| pc5 | 0.97 | 0.92 – 1.03 |
| Age | 1.78 *** | 1.71 – 1.85 |
| Sex | 0.99 | 0.93 – 1.04 |
| Marginal $R^2$ / Conditional $R^2$ | 0.048 / 0.438 | |

*Abbreviations: NDV = neurodevelopmental disorders; EUR = European ancestry; PGS = polygenic scores; pc = ancestry principal components. Reference group (sex): Male. * p<0.05 ** p<0.01 *** p<0.001*

**Supplementary Table 19.**

Hurdle Models: Associations between SCZ PGS and PQ-BC (European participants only)

| | PQ-BC | |
|---|---|---|
| *Predictors* | *std. Beta* | *standardized CI* |
| **Count Model** | | |
| (Intercept) | 5.28 *** | 5.08 – 5.49 |
| SCZ EUR PGS | 1.05 *** | 1.02 – 1.08 |
| pc1 | 0.97 * | 0.94 – 1.00 |
| pc2 | 0.96 ** | 0.93 – 0.99 |
| pc3 | 0.98 | 0.96 – 1.01 |
| pc4 | 1.01 | 0.98 – 1.04 |
| pc5 | 1.00 | 0.97 – 1.02 |
| Age | 0.84 *** | 0.82 – 0.86 |
| Sex | 1.09 *** | 1.06 – 1.12 |
| **Zero-Inflated Model** | | |
| (Intercept) | 1.65 *** | 1.41 – 1.93 |
| SCZ EUR PGS | 0.95 | 0.90 – 1.01 |
| pc1 | 1.01 | 0.96 – 1.07 |
| pc2 | 1.02 | 0.96 – 1.08 |

| | | |
|---|---|---|
| pc3 | 1.02 | 0.97 – 1.08 |
| pc4 | 0.99 | 0.94 – 1.05 |
| pc5 | 0.97 | 0.92 – 1.03 |
| Age | 1.78 *** | 1.71 – 1.84 |
| Sex | 0.98 | 0.93 – 1.04 |
| Marginal $R^2$ | 0.039 | |

*SCZ = schizophrenia; EUR = European ancestry; PGS = polygenic scores; pc = ancestry principal component. Reference group (sex): Male. * p<0.05   ** p<0.01   *** p<0.001*

**Supplementary Table 20.**

| | *PRSCS* | *PRSCS0* | *PRSET* |
|---|---|---|---|
| **M5** | 0.029** | 0.019** | 0.011 |
| **M16** | 0.019** | 0.025** | 0.005 |
| **M9** | 0.013 | 0.005 | 0.005 |
| **M14** | 0.024** | 0.022** | 0.016 |
| **M8** | 0 | 0.011 | 0.005 |
| **M12** | 0.018* | 0.024** | 0.012 |
| **M6** | 0.004 | 0.012 | 0.01 |
| **M2** | 0.012 | 0.013 | 0.015 |
| **M13** | 0.004 | 0.004 | 0.001 |
| **M3** | 0.015 | 0.021** | 0.01 |
| **M4** | 0.011 | 0.008 | 0.016 |
| **M10** | 0.003 | 0.002 | -0.004 |
| **M18** | 0.036** | 0.042** | 0.017* |
| **M17** | 0.023** | 0.031** | 0.022** |
| **M15** | 0.035** | 0.038** | 0.031** |
| **M1** | 0.035** | 0.039** | 0.029** |
| **M7** | 0.023** | 0.029** | 0.017* |
| **M11** | 0.021** | 0.029** | 0.025** |

PRSCS: effects derived by partitioning posterior effect sizes
PRSCS0: effects derived by partitioning summary statistics
PRSET: effects derived from PRSet
Dependent variable: PQ-BC
** = FDR significant
* = $p < 0.05$

**Supplementary Table 20.** Effect size estimates of ADHD pPGS on PQ-BC stratified by method of partitioning. Left-most column: pPGS derived by partitioning posterior effect sizes; middle column: pPGS derived by partitioning summary statistics; right-most column: pPGS derived by PRset default parameters. Double asterisks indicate FDR significance; single asterisks, uncorrected significance.

**Supplementary Table 21.**

| | PQ-BC Distress Score | |
|---|---|---|
| *Predictors* | *Estimates* | *CI* |
| (Intercept) | -0.18 *** | -0.25 – -0.12 |
| SCZ PrimAll | 0.02 ** | 0.01 – 0.04 |
| PC1 | -0.91 | -2.10 – 0.29 |
| PC2 | -1.33 * | -2.53 – -0.13 |
| PC3 | -0.57 | -1.75 – 0.60 |
| PC4 | 0.84 | -0.32 – 2.00 |
| PC5 | 0.16 | -1.00 – 1.32 |
| Age | -0.13 *** | -0.14 – -0.12 |
| Sex | 0.05 ** | 0.02 – 0.08 |
| Marginal $R^2$ / Conditional $R^2$ | 0.026 / 0.397 | |

* *p<0.05* ** *p<0.01* *** *p<0.001*

**Supplementary Table 21.** Estimates from SCZ PGS - PQ-BC model using multi-ancestry GWAS summary statistics. Effect of schizophrenia PGS on PQ-BC Distress score in European participants. This SCZ PGS was calculated from a GWAS of a multi–ancestry discovery sample (Trubetskoy et al. 2022). Results in EUR samples reported in the main text leverage GWAS summary statistics from the same study, but were conducted in an ancestrally homogenous set of Europeans.

**Supplementary Table 22.**

| Average baseline scores by # of complete timepoints | | | | | | | |
|---|---|---|---|---|---|---|---|
| | | **1** | **2** | **3** | **4** | **5** | **p** |
| Baseline PQ-BC | Mean (SD) | 0.4 (1.5) | 0.4 (1.3) | 0.3 (1.2) | 0.2 (1.2) | 0.2 (1.2) | 0.001 |
| Baseline IIV Composite | Mean (SD) | 0.3 (0.8) | 0.3 (0.7) | 0.2 (0.7) | 0.1 (0.6) | 0.0 (0.6) | <0.001 |

Scores are standardized (z-scored) across all timepoints

1 = 1 complete timepoint of PQBC data

2 = 2 complete timepoints of PQBC data

...

5 = 5 complete timepoints of PQBC data

p = omnibus P-value

**Supplementary Table 22.** Comparing mean baseline scores to number of complete timepoints. Cells show the mean and standard deviation of baseline PQ-BC and IIV (both z-scored) stratified by the number of complete timepoints. Group 1 comprises individuals with complete PQ-BC data for only 1 timepoint; group 5 comprises individuals with complete PQ-BC data for 5 timepoints. Values in the right-most, “p” column represent omnibus P values testing whether the mean baseline PQ-BC or IIV are equal across groups.

**Supplementary Table 23.**

| | ß | 95% CI | P |
|---|---|---|---|
| **ADHD** | | | |
| PGS*Year 1 | -0.026 | [-0.051, -0.001] | 3.93e-02* |
| PGS*Year 2 | -0.035 | [-0.061, -0.01] | 5.99e-03* |
| PGS*Year 3 | -0.042 | [-0.068, -0.017] | 1.22e-03* |
| PGS*Year 4 | -0.064 | [-0.097, -0.032] | 9.97e-05* |
| **NDV** | | | |
| PGS*Year 1 | -0.034 | [-0.059, -0.009] | 7.72e-03* |
| PGS*Year 2 | -0.044 | [-0.069, -0.018] | 7.01e-04* |
| PGS*Year 3 | -0.051 | [-0.076, -0.025] | 9.69e-05* |
| PGS*Year 4 | -0.075 | [-0.107, -0.043] | 6.02e-06* |
| **CP** | | | |
| PGS*Year 1 | 0.030 | [0.005, 0.055] | 1.80e-02* |
| PGS*Year 2 | 0.045 | [0.02, 0.07] | 4.87e-04* |
| PGS*Year 3 | 0.041 | [0.015, 0.066] | 1.88e-03* |
| PGS*Year 4 | 0.053 | [0.02, 0.086] | 1.72e-03* |
| **SCZ** | | | |
| PGS*Year 1 | 0.001 | [-0.024, 0.026] | 9.53e-01 |
| PGS*Year 2 | -0.008 | [-0.033, 0.017] | 5.44e-01 |
| PGS*Year 3 | 0.002 | [-0.024, 0.028] | 8.78e-01 |
| PGS*Year 4 | -0.005 | [-0.038, 0.028] | 7.66e-01 |

**Supplementary Table 23.** Sensitivity analyses replicating PGS by time interactions on PLEs while accounting for missingness. These models closely mirror those represented in Supplementary Tables 8-11, but include the 5-level term for data completeness (described above in Supplementary Table 22) modeled as a fixed effect covariate. Only statistics for interaction terms are represented here. Not shown are the estimates and statistics for the other fixed effects covariates (as shown in Supplementary Tables 8-11).

### Supplementary Figures

**Supplementary Figure 1. Schematic representation of data used across visits, by domain**

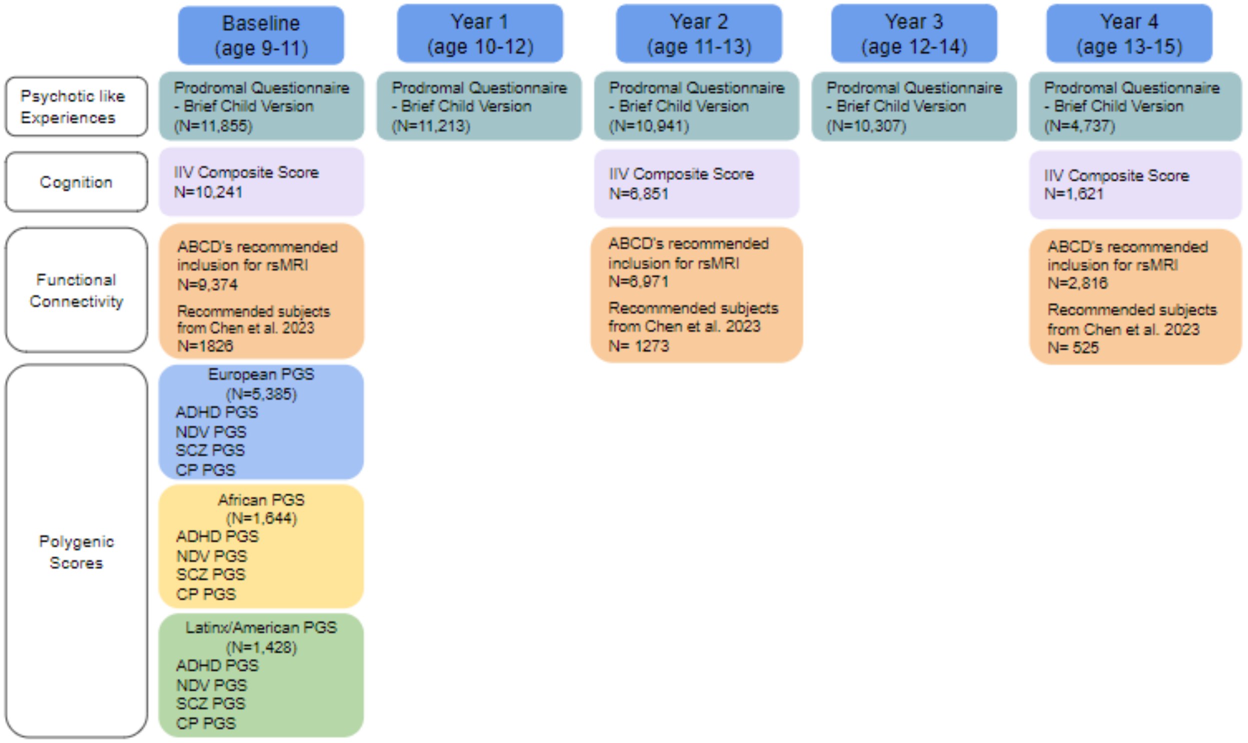


**Supplementary Figure 1.** Abbreviations: Intra-individual variability (IIV); polygenic score (PGS). PGS were derived from the Psychiatric Genomics Consortium (PGC) (Demontis et al. 2023: ADHD N=225,534, Trubetskoy et al. 2022: SCZ N=130,644, cross-disorder neurodevelopmental disorders N=113,826) and Social Science Genetic Association Consortium (Lee et al. 2018: cognitive performance (CP) N = 1,131,881; UK Biobank (Wootton et al. 2023, IIV = 404,302 individuals). Note: Data from Year 4 has only been partially released at this time.

**Supplementary Figure 2**.

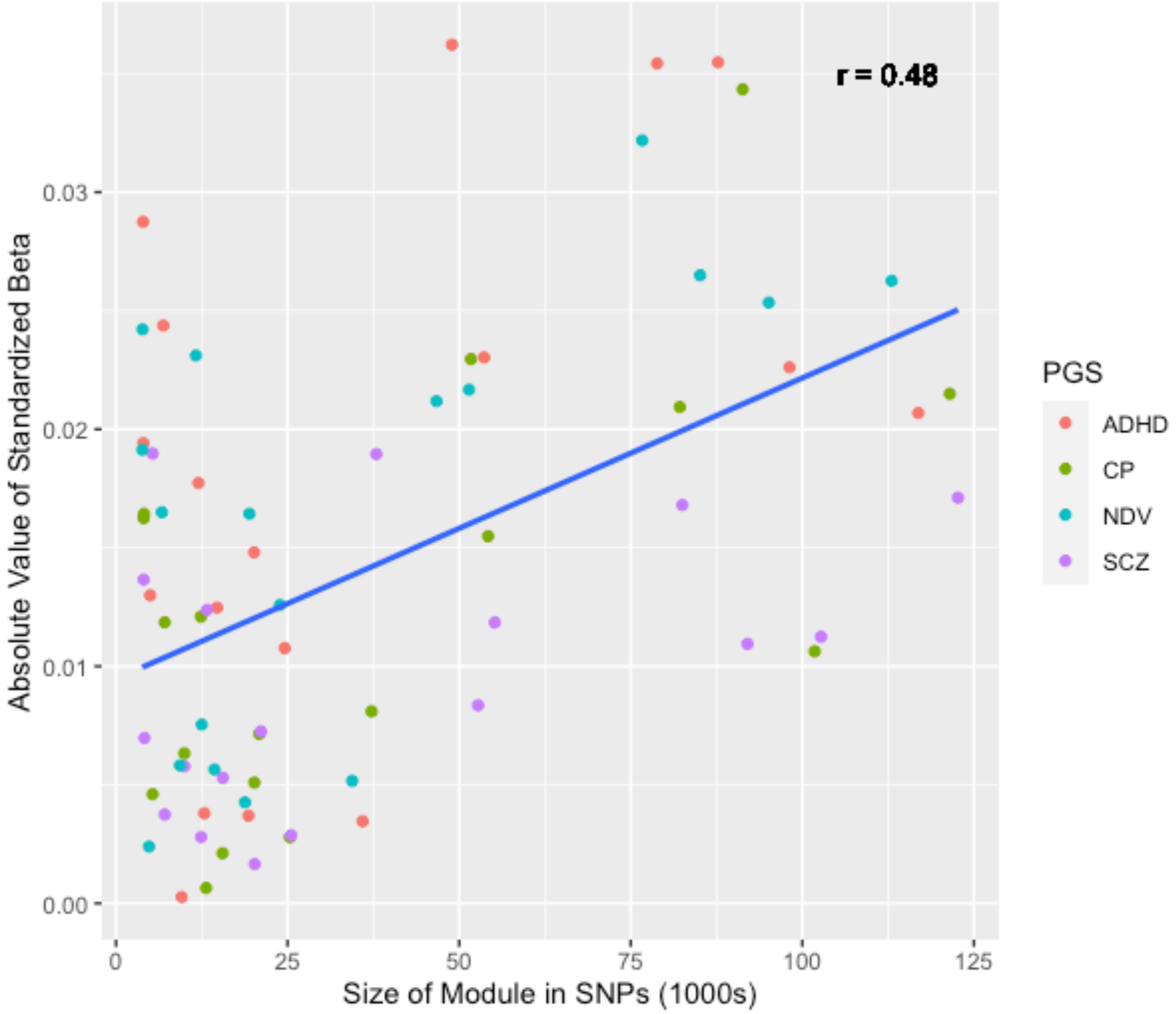


**Supplementary Figure 2.** Relationship between module size and effect size for pPGS. Plotted on the y-axis are the standardized betas from models with modular pPGS predicting PQ-BC in the EUR sample. Represented on the x-axis are the sizes of the corresponding modules in 1000s of SNPs. Pearson's r correlation coefficient printed in the top right corner of the figure.

**Supplementary Figure 3.**

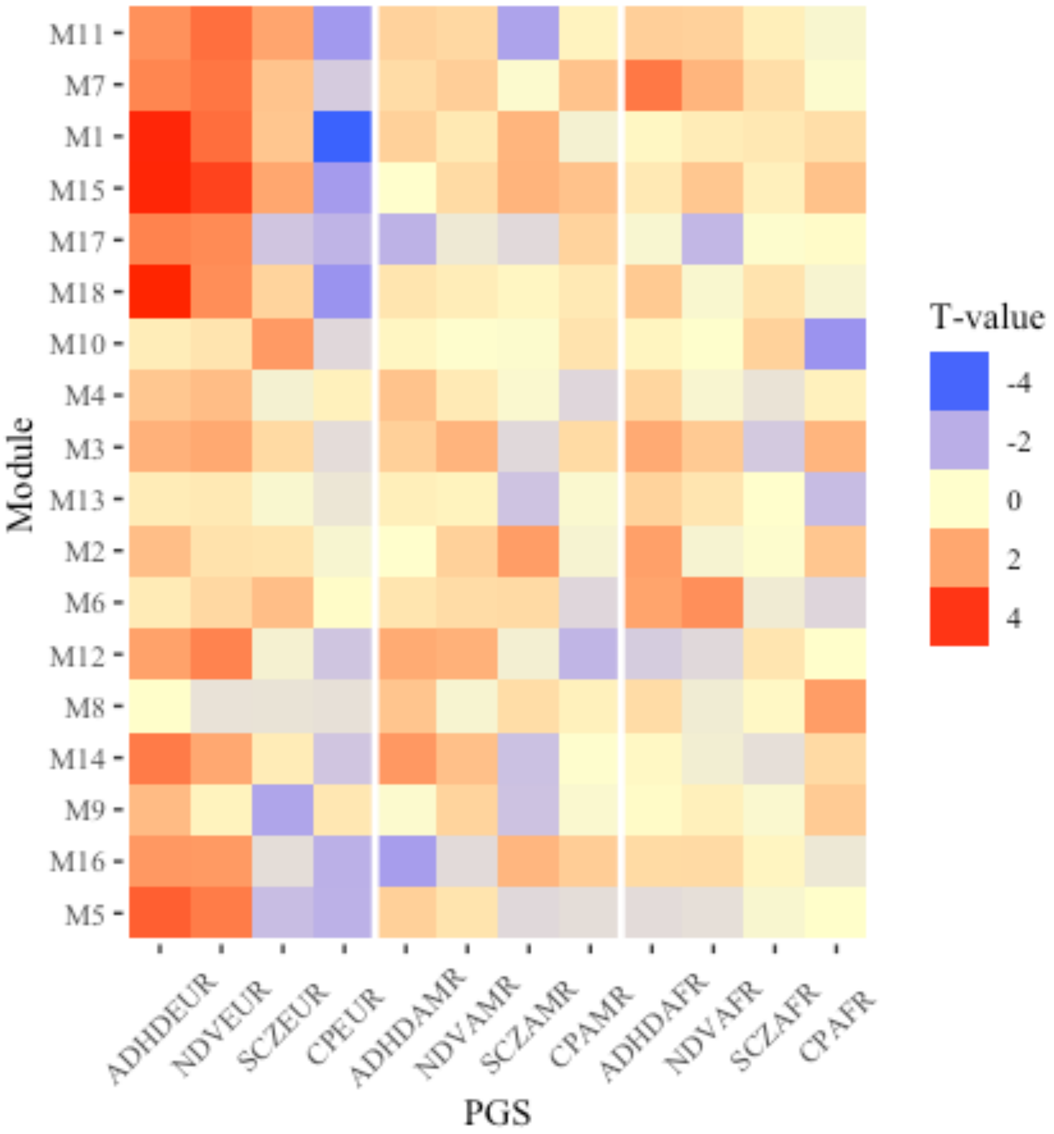


**Supplementary Figure 3.** Heatmap of pPGS effects on PQ-BC in European, American/Latinx, and African samples. Modules are ordered from bottom to top from smallest to largest module. Each column represents a different pPGS with ancestry appended to PGS name (e.g., ADHDAMR = ADHD pPGS in American/Latinx population). Besides associations in EUR (see **Figure 4**), no effects represented here were significant after FDR correction (number of tests = 216). The left-most group of columns (EUR results) is identical to Figure 4. ADHD = attention deficit/hyperactivity disorder; NDV = neurodevelopmental disorders; SCZ = schizophrenia; CP = cognitive performance; EUR = European ancestry; AMR = American/Latinx; AFR = African.

**Supplementary Figure 4.**

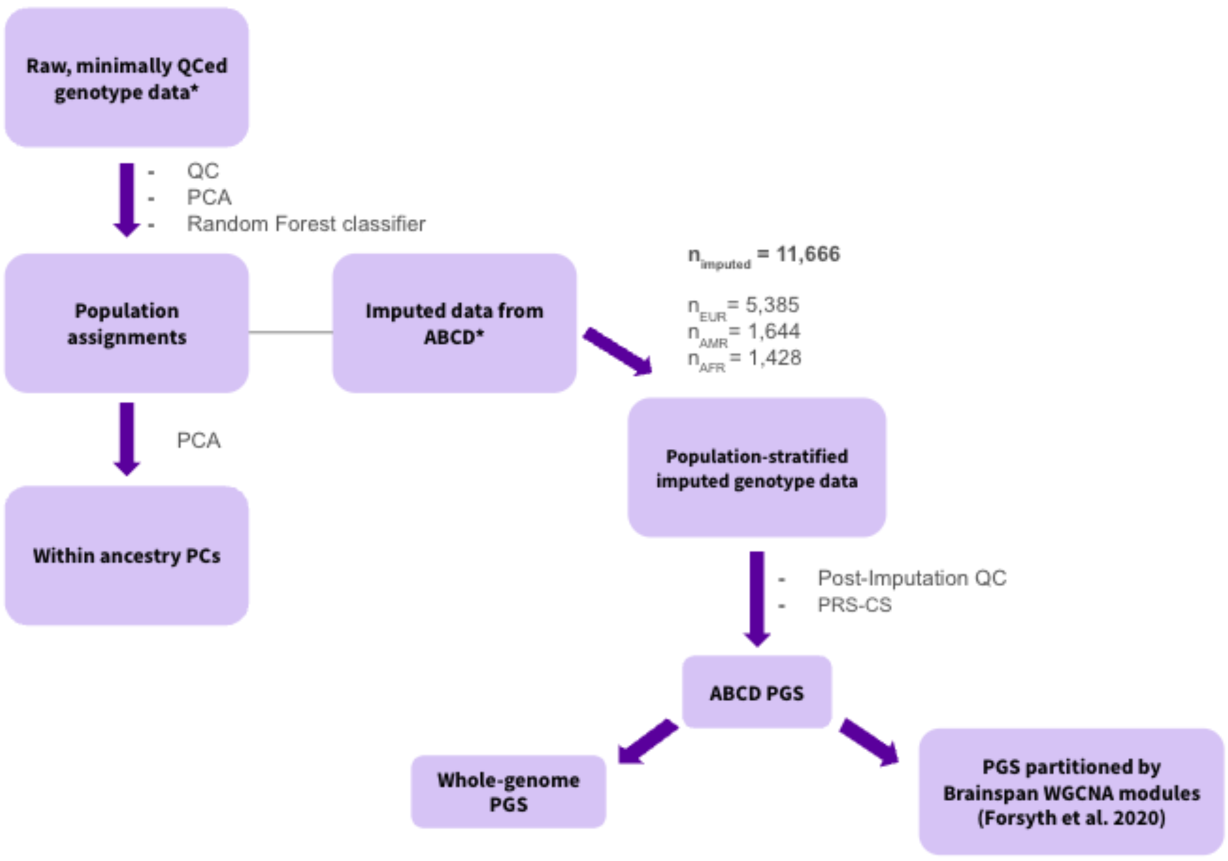


**Supplementary Figure 4.** Overall polygenic scores calculation workflow. Diagram representing workflow of generating polygenic scores. Asterisks indicate data which were provided by and downloaded directly from ABCD.

**Supplementary Figure 5.**

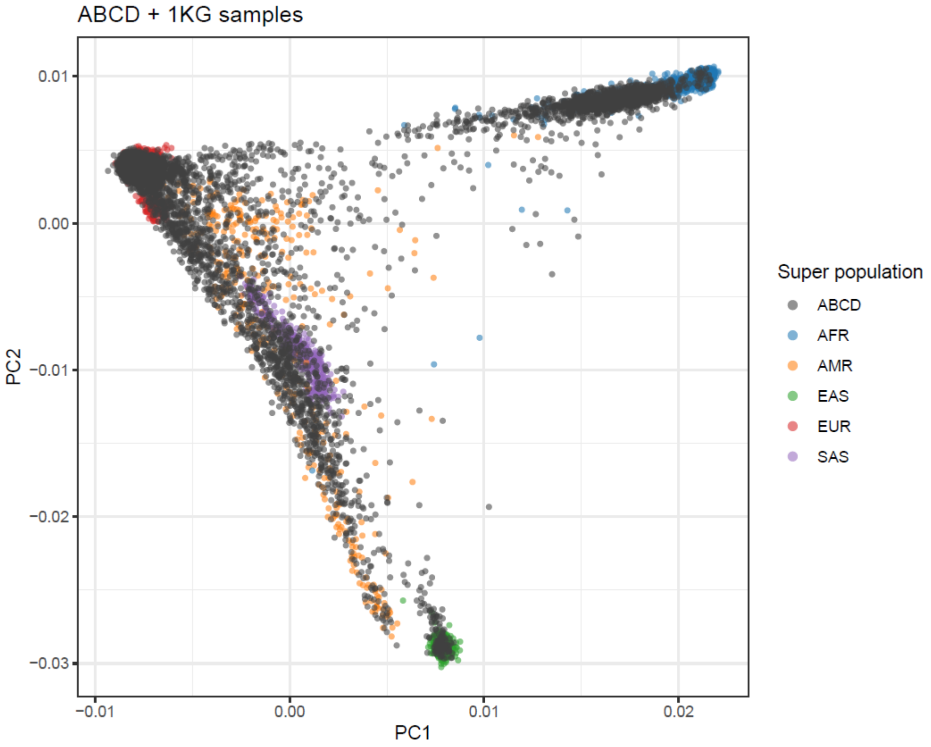


**Supplementary Figure 5.** Visualization of principal components for ancestral assignment. Scatter plot of top two principal components with ABCD data points superimposed on 1KG data points. Data points in gray represent ABCD participants; blue, yellow, green, red, and purple represent data points from the 1KG sample. All 5 superpopulations are shown here (African (AFR), American/Latinx (AMR), East Asian (EAS), European (EUR), and South Asian (SAS)); however, only AFR, AMR, and EUR participants were included for main analyses, given the small N's for EAS and SAS .

Included Ancestries:
N (European) = 5,385
N (African) = 1,644
N (Latinx/AMR) = 1,428

**Supplementary Figure 6.**

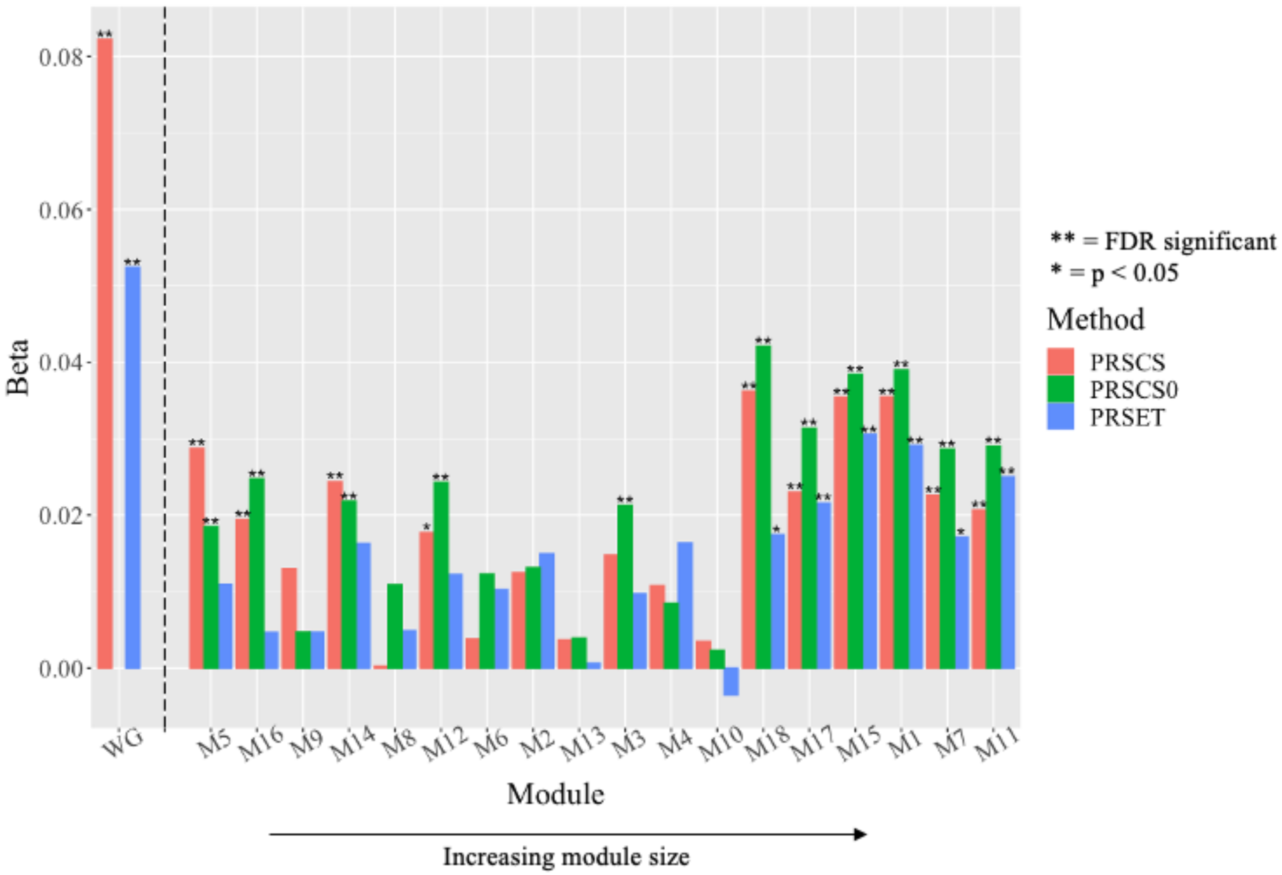


**Supplementary Figure 6.** Comparison of ADHD PGS by method of partitioning. Y-axis plots the effect size of each pPGS on unstandardized PQ-BC Modules are ordered from left to right from smallest to largest module. Red (PRSCS) represents effects of PRS-CS-derived pPGS where posterior effect sizes were partitioned by module membership; green (PRSCS0) represents those from PRS-CS-derived pPGS where summary statistics were partitioned by module membership; blue represents effects from PRSet-derived pPGS. The dependent variable is PQ-BC. Single star represents uncorrected significance and a double star indicates significance after FDR correction. Effects represented to the left of the dotted vertical line, labeled WG on the x-axis, are of whole genome PGS, where red represents effects of PGS derived by PRSCS and blue of those derived by PRSice, which PRSet leverages for its PGS calculation.

**Supplementary Figure 7**

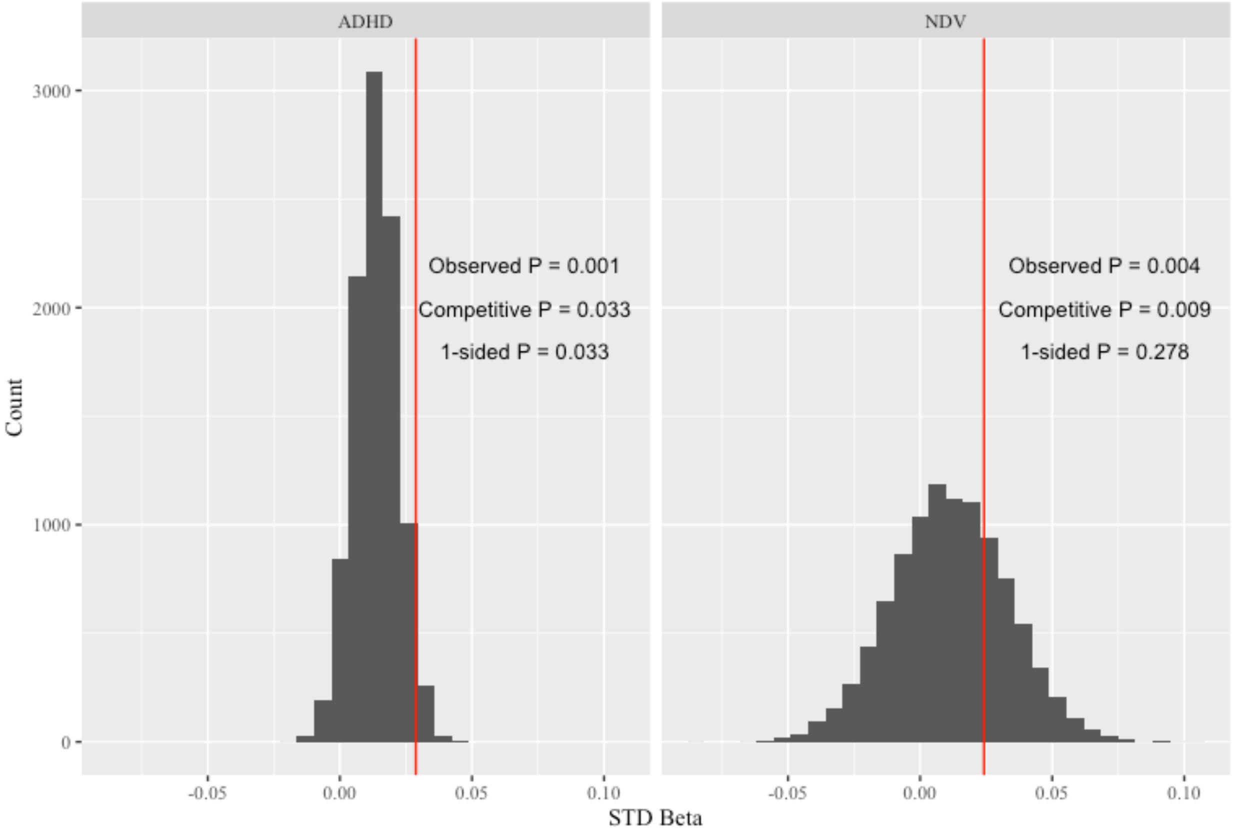


**Supplementary Figure 7.** Graphical depiction of criteria for a module pPGS to be considered as significantly associated with PQ-BC due to the intrinsic biological properties of that module. On the left is the permuted, null distribution of ADHD M5 pPGS with the red vertical line representing the observed effect (standardized ß). On the right is the analagous distribution for NDV M5 pPGS. The observed, competitive, and one-sided permuted P are labeled to the right of each distribution. For NDV M5, although the observed P-value passes FDR correction and the competitive P is less than 0.05, the one-sided permuted P is greater than 0.05 and thus did not pass our stringent criteria.

**Supplementary References**


Carlozzi, N. E., Tulsky, D. S., Kail, R. V., & Beaumont, J. L. (2013). VI. NIH Toolbox Cognition Battery (CB): Measuring processing speed. *Monogr. Soc. Res. Child Dev.*, *78*(4), 88–102.

Chen, J., Tam, A., Kebets, V., Orban, C., Ooi, L. Q. R., Asplund, C. L., Marek, S., Dosenbach, N. U. F., Eickhoff, S. B., Bzdok, D., Holmes, A. J., & Yeo, B. T. T. (2022). Shared and unique brain network features predict cognitive, personality, and mental health scores in the ABCD study. *Nat. Commun.*, *13*(1), 2217.

Choi, S. W., García-González, J., Ruan, Y., Wu, H. M., Porras, C., Johnson, J., Bipolar Disorder Working group of the Psychiatric Genomics Consortium, Hoggart, C. J., & O'Reilly, P. F. (2023). PRSet: Pathway-based polygenic risk score analyses and software. *PLoS Genet.*, *19*(2), e1010624.

Cicero, D. C., Krieg, A., & Martin, E. A. (2019). Measurement Invariance of the Prodromal Questionnaire–Brief Among White, Asian, Hispanic, and Multiracial Populations. *Assessment*, *26*(2), 294–304.

Demontis, D., Walters, G. B., Athanasiadis, G., Walters, R., Therrien, K., Nielsen, T. T., Farajzadeh, L., Voloudakis, G., Bendl, J., Zeng, B., Zhang, W., Grove, J., Als, T. D., Duan, J., Satterstrom, F. K., Bybjerg-Grauholm, J., Bækved-Hansen, M., Gudmundsson, O. O., Magnusson, S. H., … Børglum, A. D. (2023). Genome-wide analyses of ADHD identify 27 risk loci, refine the genetic architecture and implicate several cognitive domains. *Nat. Genet.*, *55*(2), 198–208.

Forsyth, J. K., Nachun, D., Gandal, M. J., Geschwind, D. H., Anderson, A. E., Coppola, G., & Bearden, C. E. (2020). Synaptic and Gene Regulatory Mechanisms in Schizophrenia, Autism, and 22q11.2 Copy Number Variant–Mediated Risk for Neuropsychiatric Disorders. *Biol. Psychiatry*, *87*(2), 150–163.

Ge, T., Chen, C.-Y., Ni, Y., Feng, Y.-C. A., & Smoller, J. W. (2019). Polygenic prediction via Bayesian regression and continuous shrinkage priors. *Nat. Commun.*, *10*(1), 1776.

Hagler, D. J., Jr, Hatton, S., Cornejo, M. D., Makowski, C., Fair, D. A., Dick, A. S., Sutherland, M. T., Casey, B. J., Barch, D. M., Harms, M. P., Watts, R., Bjork, J. M., Garavan, H. P., Hilmer, L., Pung, C. J., Sicat, C. S., Kuperman, J., Bartsch, H., Xue, F., … Dale, A. M. (2019). Image processing and analysis methods for the Adolescent Brain Cognitive Development Study. *Neuroimage*, *202*, 116091.

Hughes, D. E., Kunitoki, K., Elyounssi, S., Luo, M., Bazer, O. M., Hopkinson, C. E., Dowling, K. F., Doyle, A. E., Dunn, E. C., Eryilmaz, H., Gilman, J. M., Holt, D. J., Valera, E. M., Smoller, J. W., Cecil, C. A. M., Tiemeier, H., Lee, P. H., & Roffman, J. L. (2023). Genetic patterning for child psychopathology is distinct from that for adults and implicates fetal cerebellar development. *Nat. Neurosci.*, *26*(6), 959–969.

Karcher, N. R., Barch, D. M., Avenevoli, S., Savill, M., Huber, R. S., Simon, T. J., Leckliter, I. N., Sher, K. J., & Loewy, R. L. (2018). Assessment of the Prodromal Questionnaire–Brief Child Version for Measurement of Self-reported Psychoticlike Experiences in Childhood. *JAMA Psychiatry*, *75*(8), 853–861.

Karcher, N. R., Paul, S. E., Johnson, E. C., Hatoum, A. S., Baranger, D. A. A., Agrawal, A., Thompson, W. K., Barch, D. M., & Bogdan, R. (2022). Psychotic-like Experiences and Polygenic Liability in the Adolescent Brain Cognitive Development Study. *Biol Psychiatry Cogn Neurosci Neuroimaging*, *7*(1), 45–55.

Lee, J. J., Wedow, R., Okbay, A., Kong, E., Maghzian, O., Zacher, M., Nguyen-Viet, T. A., Bowers, P., Sidorenko, J., Karlsson Linnér, R., Fontana, M. A., Kundu, T., Lee, C., Li, H., Li, R., Royer, R., Timshel, P. N., Walters, R. K., Willoughby, E. A., … Cesarini, D. (2018). Gene discovery and polygenic prediction from a genome-wide association study of educational attainment in 1.1 million individuals. *Nat. Genet.*, *50*(8), 1112–1121.

Loewy, R. L., Pearson, R., Vinogradov, S., Bearden, C. E., & Cannon, T. D. (2011). Psychosis risk screening with the Prodromal Questionnaire–brief version (PQ-B). *Schizophr. Res.*, *129*(1), 42–46.

Trubetskoy, V., Pardiñas, A. F., Qi, T., Panagiotaropoulou, G., Awasthi, S., Bigdeli, T. B., Bryois, J., Chen, C.-Y., Dennison, C. A., Hall, L. S., Lam, M., Watanabe, K., Frei, O., Ge, T., Harwood, J. C., Koopmans, F., Magnusson, S., Richards, A. L., Sidorenko, J., … Schizophrenia Working Group of the Psychiatric Genomics Consortium. (2022). Mapping genomic loci implicates genes and synaptic biology in schizophrenia. *Nature*, *604*(7906), 502–508.

Wootton, O., Shadrin, A. A., Mohn, C., Susser, E., Ramesar, R., Gur, R. C., Andreassen, O. A., Stein, D. J., & Dalvie, S. (2023). Genome-wide association study in 404,302 individuals identifies 7 significant loci for reaction time variability. *Mol. Psychiatry*, *28*(9), 4011–4019.

Zelazo, P. D., Anderson, J. E., Richler, J., Wallner-Allen, K., Beaumont, J. L., & Weintraub, S. (2013). II. NIH Toolbox Cognition Battery (CB): Measuring executive function and attention. *Monogr. Soc. Res. Child*

*Dev.*, *78*(4), 16–33.
